# Decoding personal and population variability in the human multiome

**DOI:** 10.64898/2026.10.28.26366658

**Authors:** Daniel J. Panyard, Martín Acosta Parra, Aubrey K. Roberts, Hirotaka Ieki, Tejaswini Mishra, Nasim Bararpour, Anisha Jaiswal, Sophia M. Schüssler-Fiorenza Rose, Kévin Contrepois, Daniel Hornburg, Sara Ahadi, Xiaotao Shen, Themistocles L. Assimes, Michael P. Snyder

## Abstract

**Background:** With recent advances in technology, multiomic data have proliferated rapidly. Biobanks are now developing genomic, proteomic, lipidomic, and metabolomic data sets on the order of hundreds of thousands of people. However, bridging the gap between population-level data and the individual—a major challenge in precision medicine—remains a novel and challenging task.

**Methods:** We analyzed the landscape of contemporary multiomics data and the drivers of population and personal molecular variability using a longitudinal cohort uniquely suited to precision health applications. Using healthy routine visit data (94 participants, median five visits per participant), we profiled the human multiome across eight molecular omic types and 1.2 million measurements, with replication in a sister multiomic cohort (322 participants).

**Results:** Molecular data tended to cluster by omes, with the metabolome and proteome showing less intraomic correlation than other data types. While most blood-based molecular measurements followed log-normal distributions, between 2.0% and 8.1% of molecules showed evidence of bimodality, with cases driven by genetic polymorphisms or dietary differences (e.g., SERPINA5 [P = 1.4e-6], 2-piperidinone [P = 5.7e-5]). Molecular variation across the population was generally higher than that within an individual over time (e.g., xenobiotic metabolites, progestin steroids). Analyzing personal molecular variation identified several participants with notable temporal shifts (intraindividual CV > 4 SDs higher than population CV) in free fatty acids (FFA 20:1 and FFA 20:2) or cytokines (IL-17F and IFN-beta), which were unlikely to be detected cross-sectionally. Body mass index (BMI) had a much larger multiomic footprint (23.6% of molecules associated) than demographic factors like sex or age (5.7% and 1.9%, respectively), although machine-learning predictive models of sex (top ROC AUC of 0.98) were more successful than those for BMI (top R^2^ of 0.59 by cytokines) or age (top R^2^ of 0.34 by metabolites). Purely multiomic predictive models explained modest to high proportions of variation (R^2^ = 0.22 to 0.94) in 20 common clinical tests, highlighting the potential clinical utility of these data even within a modestly sized cohort.

**Conclusion:** Analysis of the molecular distributions, variation, and covariation of longitudinal multiomics data revealed clinically relevant differences at both the population and personal levels. Based on these findings, we conclude with six key considerations for the next generation of population omics as the field advances toward the goals of precision medicine.

Multiomic data have proliferated rapidly in biomedical research, but many aspects of molecular variability remain poorly understood. We investigated in detail the drivers of personal and population variability in a longitudinal, multiomic cohort uniquely suited to precision health applications (94 participants, eight omic types, 1.2 million molecular measurements), with replication in a sister multiomic cohort (322 participants). While most molecules followed log-normal distributions, between 2.0% and 8.1% of molecules were bimodal, with cases driven by genetic polymorphisms or dietary differences (e.g., SERPINA5 [P = 1.4e-6], 2-piperidinone [P = 5.7e-5]). The metabolome and proteome showing less intraomic correlation than the lipidome or cytokines, and molecular variation across the population was generally higher than that within an individual over time with. Analyzing personal molecular variation identified several participants with notable temporal shifts (intraindividual CV > 4 SDs higher than population CV) in free fatty acids (FFA 20:1 and FFA 20:2) or cytokines (IL-17F and IFN-beta), which were unlikely to be detected cross-sectionally. Body mass index (BMI) was more broadly associated with molecular levels (23.6% of molecules associated) than were demographic factors like sex or age (5.7% and 1.9%, respectively), although machine-learning predictive models of sex (top ROC AUC of 0.98) were more successful than those for BMI (top R^2^ of 0.59 by cytokines) or age (top R^2^ of 0.34 by metabolites). Purely multiomic predictive models were modest to highly predictive (R^2^ = 0.22 to 0.94) of 20 common clinical tests, highlighting the potential clinical utility of these data even within a modestly sized cohort. Our findings reveal six key considerations (missing variability, temporality, feasible clinicomics, complex modeling, aging complexity, and molecular matching) for precision medicine.

## Introduction

Precision medicine, the “prevention and treatment strategies that take individual variability into account”, remains one of the major goals of 21st century science^1^. This idea is by no means novel nor theoretical: clinical care has long sought more individualized treatments, from the matching of blood transfusions to a patient’s blood type^1^ to the effort to develop a “New Taxonomy of Disease”^2^. Much of the progress in precision medicine has been driven by technology. Improvements in genotyping and sequencing have powered precision medicine successes in cystic fibrosis care, oncology, and pharmacogenomics^3^. More recently, high-throughput and high-plex measurement of molecules (e.g., proteins, metabolites, and lipids) at the population scale has become possible. There has been a rapid increase in biobank multiomics at the scale of tens to hundreds of thousands of individuals, such as in the UK Biobank^4^, deCODE Genetics^5^, FinnGen^6^, BioBank Japan^7^, Precision Health Research Singapore’s SG100K^8^, the Million Veteran Program^9^, and TOPMed^10^. These massive data sets hold enormous potential for revolutionizing our understanding of diseases at the molecular level.

However, to understand how the molecular landscape changes in disease, we must first understand how it looks in health. Characterizing typical molecular distributions requires an understanding of how these molecules vary across populations, environments, and lifestyles. Researchers have begun to explore how omics data vary by characteristics like age, sex, body size, and common clinical metrics, but these studies are often constrained by limitations in longitudinality, number of omes included, and assumptions about molecular distributions that may obscure clinically relevant patterns^11–15^.

The goals of this study were to 1) assess patterns of molecular variation at the population and personal levels; 2) identify and contrast the drivers of molecular covariation; and 3) develop multiomic machine-learning models of demographic, anthropomorphic, and clinical measures (Figure 1). To achieve these goals, we analyzed a prototypical precision medicine cohort, the Integrated Personal Omics Profiling (iPOP) study, that is observational, longitudinal, and multiomic. We focused on healthy visits of 94 people, each with an average of 5 visits of data with 8 different omes quantified. Collectively, these analyses reveal how precision-health multiomics requires attention to distribution, temporality, and molecular context. We conclude with six key insights from our results that provide insight into how researchers can best take advantage of rapidly growing multiomic data to move closer to the goal of precision health.

**Figure 1.**
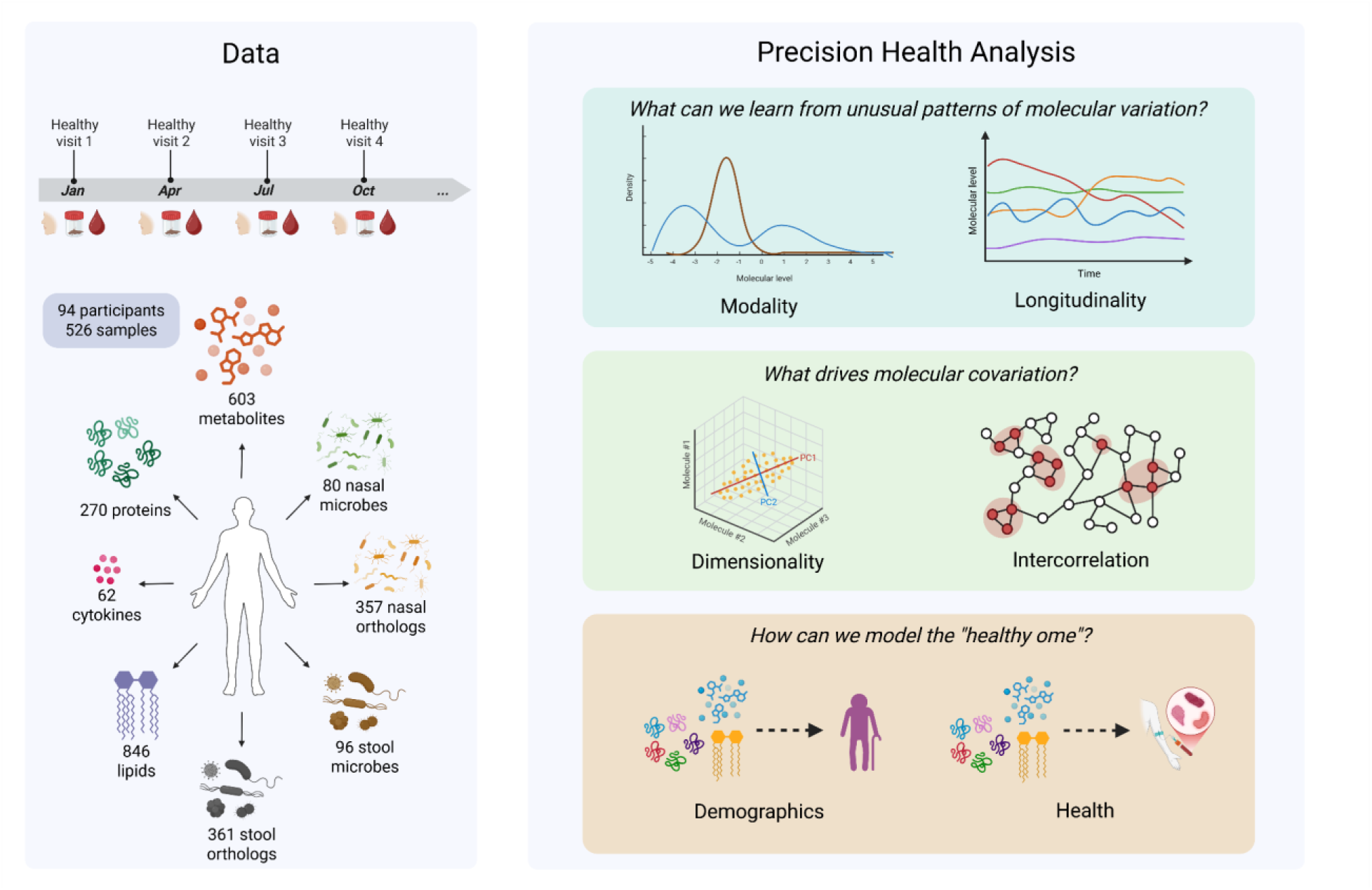
Study overview. The sampling strategy and data generation from the iPOP cohort are summarized on the left side. In total, 94 participants provided blood, stool, and nasal swab samples at quarterly “healthy” visits, with each participant attending an average of 5 visits (526 total visits). Up to 8 different omes of data were generated from each visit. Our analyses focused on understanding the healthy human multiome from a variety of perspectives. Using these data, we explored the following questions: 1) how patterns of molecular variation connect to health and biology; 2) how intermolecular covariation reflects existing technology and biology; and 3) how we can train useful machine-learning models on complex, low-structure multiomics data to capture high-level demographic and clinical measures.

## Results

### Cohort

A total of 526 routine “healthy” visits from 94 participants were analyzed, with a median of 5 visits per participant (Figure 1; Supplementary Figure 1; Supplementary Table 1), with visits occurring roughly every 3.4 (SD = 0.2) months and covering a mean span of 1.7 (SD = 1.2) years (Supplementary Figure *2*; Supplementary Figure *3*). The cohort was primarily older middle-aged adults (mean age = 55.4 years; SD = 9.9), though ages ranged from the 30s through 70s (Supplementary Figure 4). The participants were roughly evenly split by sex. Participant BMIs were most often overweight (44.7%), with a BMI distribution mostly in line with the national average except for fewer severely obese (1.1%) and more healthy BMIs (24.5%)^16^ (Supplementary Figure 5). The race/ethnicity distribution was mostly self-described as white (69.1%) and Asian (20.2%).

The number of samples with data from each ome varied, ranging from 386 visits with stool microbiome data at the lowest to 488 visits with metabolomic data at the highest (Supplementary Figure *6*). Most visits (n = 452; 85.9%) had all of the blood molecular data (proteins, cytokines, lipids, and metabolites). A total of 295 visits (56.1%) had each type of omic data. The number of features by omic type varied substantially, with the fewest number of features in the cytokine panel (62 cytokines) and the most in the lipids (846 lipid species) (Supplementary Figure *7*; Supplementary Table *2*). A total of 49 clinical tests were available for analysis (Supplementary Table *3*).

### Dimensionality analysis

To understand the density of information captured by each ome, we conducted a PCA of each omic data set. The percentage of omic variation captured by the first PC varied substantially by ome, with lipids (44.7%) and cytokines (37.9%) at the higher end and proteins (7.9%) and metabolites (8.8%) at the lower end (Figure *2*; Supplementary Figure *8*; Supplementary Table *4*). By the fourth PC, the amount of new variance explained was around 5-8% per PC for each ome. From the perspective of the cumulative variance explained in an “avalanche plot”, the proteomic and metabolomic data sets were more information-rich. For example, to account for 50% of the total variance of the ome, 12 PCs were needed for both the proteomic and metabolomic data sets compared to just 3 or fewer PCs needed for cytokines, lipids, and both ortholog data sets.

**Figure 2.**
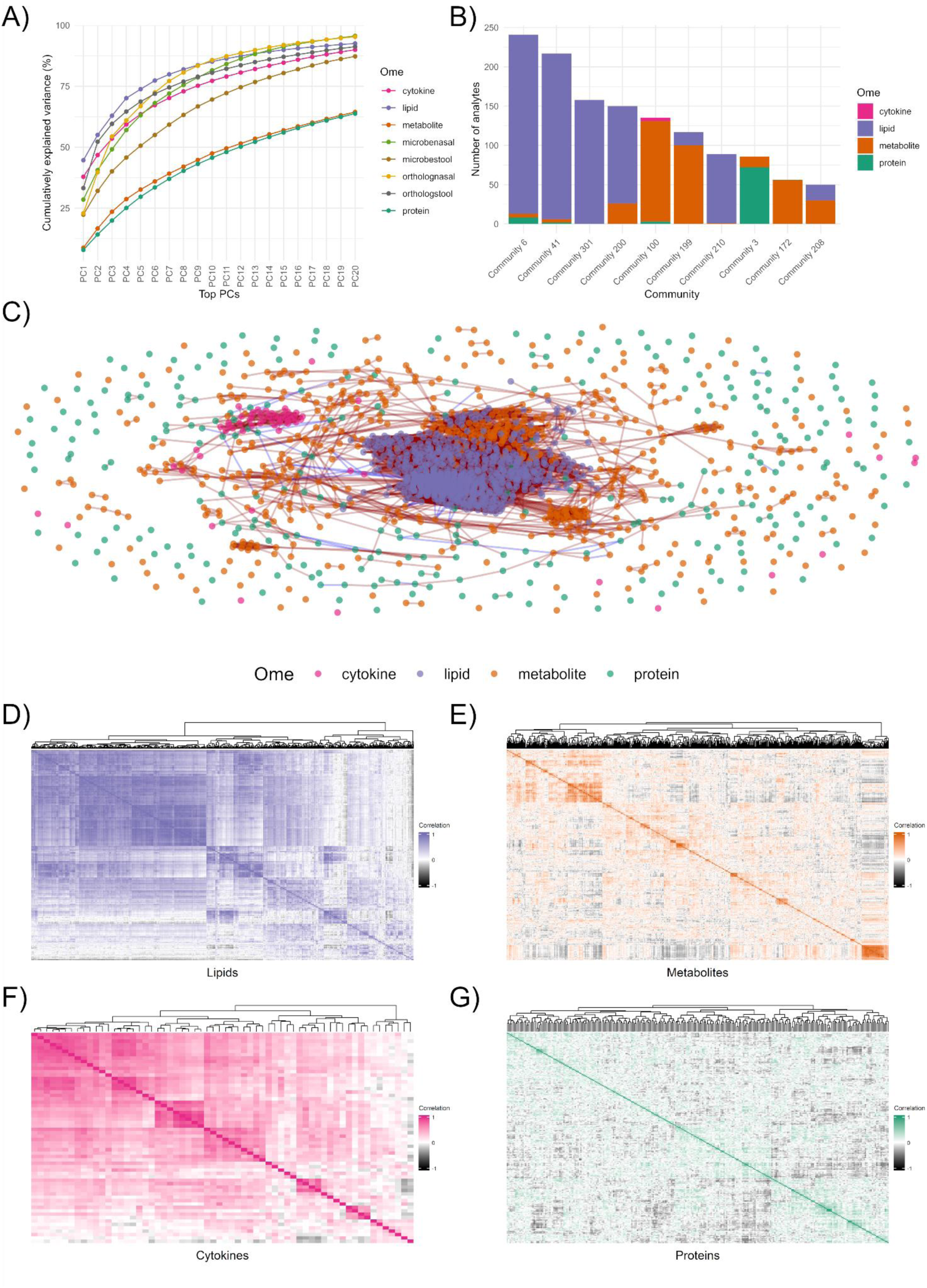
Intraomic and interomic covariation. **A)** An avalanche plot from the multiomic PCA is shown, highlighting the cumulative variation captured by successive principal components for each ome. Differences in the information captured by each data set appear as different slopes to the curves, reflecting intraomic correlation patterns and differences in quantification. **B)** Membership of the different communities identified in a Gaussian graphical model (GGM) derived from partial correlations among the blood omic data is shown. The top 9 largest communities are broken down by ome. Most communities were dominated by one primary ome. **C)** The network from the GGM analysis is shown. Edges represent partial correlations, with red indicating positive correlations and blue negative correlations. Each point represents a different molecule measured in the data set, colored by ome. **D-G)** Intraomic correlation heat maps are shown for **D)** lipids, **E)** metabolites, **F)** cytokines, and **G)** proteins. The ome’s theme color indicates positive correlation, black indicates negative correlation, and white indicates no correlation. Different correlation patterns are seen across different omes, with lipids and cytokines showing much larger blocks of correlated variables than metabolites and proteins.

We tested these results for replication in the Human Personalized Omics Profiling (hPOP) cohort^17^, a multiomic cohort with 322 participants from around the world that includes proteomic, metabolomic, lipidomic, and other data. Despite differences in the study population, technologies used, and the number of analytes measured per ome, similar trends in dimensionality between omes were observed in hPOP. As in the iPOP cohort, the metabolomic and proteomic data sets were more dimensionally rich than lipidomics. About proteomic 9 and 11 untargeted metabolomic PCs were needed to capture 50% of the variation in their respective data sets, compared to just 1 PC needed for the lipidomics data (Supplementary Figure *9*). The hPOP cohort targeted plasma metabolomics data was more similar to the iPOP lipidomics data in terms of needing only 2 PCs to capture 50% of the variance, whereas the untargeted urine metabolomics was much more similar to the iPOP untargeted plasma metabolomics (18 PCs needed).

Intraomic correlations were calculated for each ome and represented by correlation heat maps (Figure *2*; Supplementary Figure *10*; Supplementary Figure *11*; Supplementary Figure *12*; Supplementary Figure 13; Supplementary Table *5*). These correlation structures generally reflected the findings from the PCA avalanche plots: the omes that needed more PCs to capture some amount of the variance were also the omes with weaker correlation structure. Fewer blocks of tightly correlated molecules were observed for the proteins and metabolites than for the lipids or cytokines.

A Gaussian Graphical Model (GGM) network analysis of the blood omes (proteins, cytokines, lipids, and metabolites) indicated that the molecules tended to cluster by ome according to their partial correlations (Figure *2*). Of the largest network communities identified, one ome generally dominated the membership of each community, though the molecular subtypes varied by community (Supplementary Table 6). The largest community (Community 6) had 241 members and was dominated by lipids (esp. CEs, CERs, PCs, PEs, PIs, and SMs) along with betaine and several lipids from the metabolomic data set (e.g., lysoPC (14:0) and lysoPE (16:1)) and several apolipoproteins from the proteomic data set (e.g., APOM, APOC3, APOB, and APOD). This community also included proteins involved in iron homeostasis, coagulation, amino acid metabolism, and immune regulation, such as ceruloplasmin, prothrombin, alanine-glyoxylate aminotransferase (AGXT), and pregnancy zone protein (PZP). The next largest community (Community 41) was again dominated by lipids, but primarily by DAGs and TAGs, while also including proteins APOC2 and APOC4 and metabolites glycine and L-serine.

The network itself reinforces the dominance of intraomic relationships, with clear clustering of the lipids and cytokines and more nuanced (but still distinct) clusters of metabolites and proteins (Figure *2*). A total of 251 molecules were not included in a community with any other molecule, including 11 cytokines, 90 metabolites, and 150 proteins. The top molecules by degree and hub score were lipids (specifically, TAGs and DAGs), likely reflecting the large number and intercorrelation of these molecules in our lipidomic data set.

### Modality analysis

In multiomic analysis, it is common to assume that molecular variables are normally distributed, often after a log or other transformation. To investigate the extent to which these assumptions hold true, we analyzed the modality of each molecular variable. After QC and transformations, the majority of each ome’s variables were roughly normal and unskewed (skew between −2 and 2), although the proportion of unskewed variables varied, from 58% (metabolites) to 99% (lipids) (Supplementary Figure *14*; Supplementary Table *7*). However, when a stacked density plot of all variables for a given ome was created, it became evident that some individual variables had peaks of data far from the mean, indicating potential multimodality (Figure *3*).

**Figure 3.**
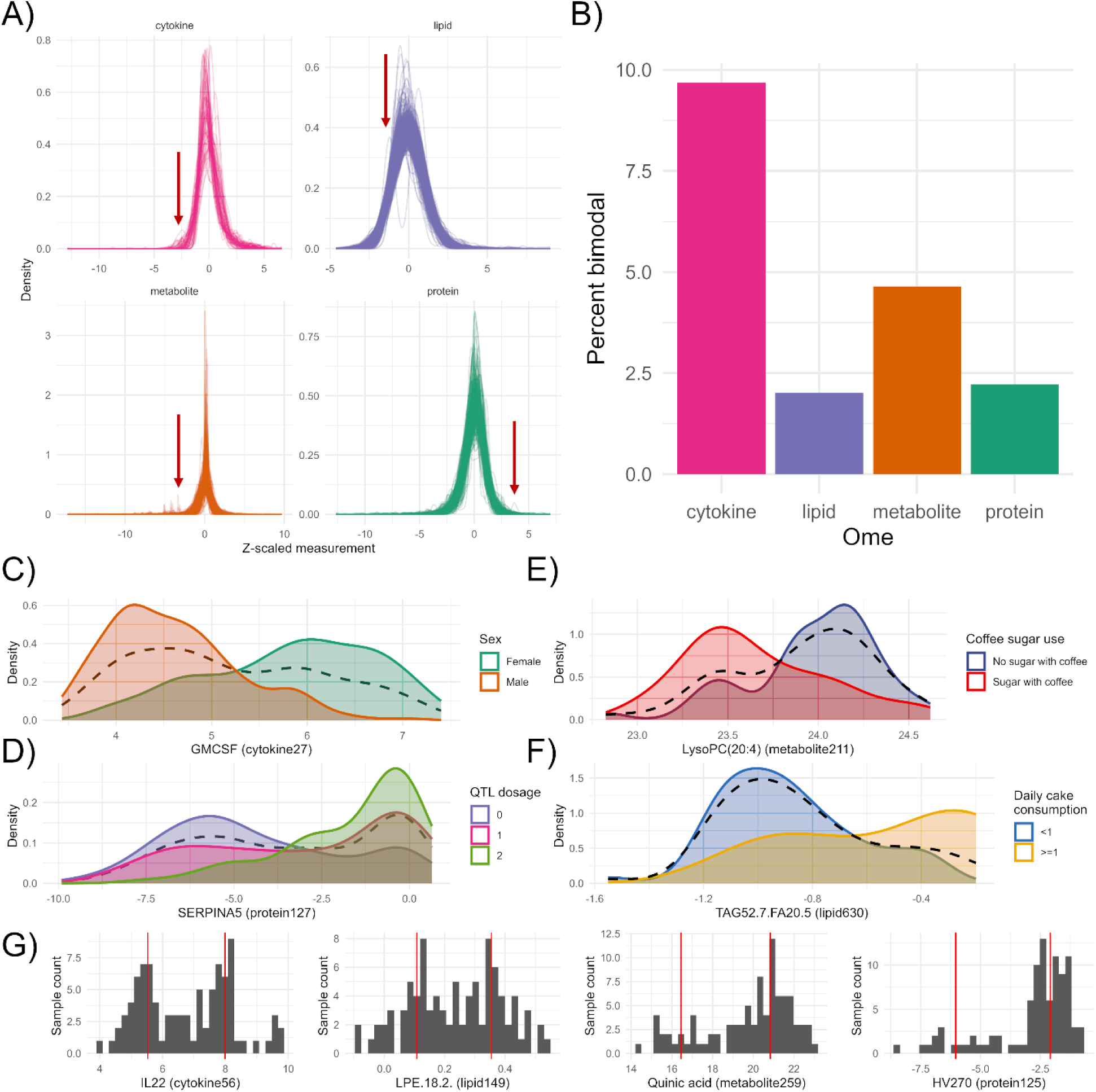
Distributions and modality in the plasma multiome. Evidence of non-normally distributed molecular measurements in the blood is shown. **A)** Stacked distribution density plots for all measured plasma cytokines, lipids, metabolites, and proteins are shown (transformed median values per molecule per participant). While the majority of molecules appear normally distributed (indicating that normal distributional assumptions may be appropriate in general), secondary peaks can be seen in all four omes (red arrows). **B)** The proportion of molecules in each ome with potential evidence of bimodality (bimodal index, BI > 1.3) is shown. **C)** The distribution of granulocyte-macrophage colony-stimulating factor (GM-CSF; cytokine27) is shown. The dashed black line shows the overall distribution of GM-CSF, which showed evidence of bimodality (BI = 1.52). This bimodality is largely explained by sex and BMI, with a clear separation of females and males when stratified (green and orange distributions, respectively). **D)** The distribution of plasma SERPINA5 levels is shown. The dashed black line shows the overall distribution of SERPINA5, which showed evidence of bimodality (BI = 1.84). When SERPINA5 levels were stratified by the allele dosage for a known pQTL (rs2731674), two peaks corresponding to the two forms of the alleles were observed. Homozygous dominant individuals (purple) tended to have lower levels of SERPINA5 than homozygous recessive individuals (green), with heterozygous individuals (pink) having an intermediate distribution. **E)** The distribution of plasma lysophosphatidylcholine 20:4 (metabolite211) is shown. The dashed black line shows the overall distribution of lysoPC 20:4, which showed evidence for bimodality (BI = 1.37). This bimodality is partly explained by presumed coffee use, with lower levels among participants who reported using sugar with coffee. **F)** The distribution of plasma triacylglyceride 52:7 and fatty acid 20:5 (lipid630) is shown. The dashed black line shows the overall distribution of the compound, which showed evidence for bimodality (BI = 1.39). This bimodality is partly explained by reported cake consumption, with lower levels among participants reporting less cake consumption. **G)** The distributions of four bimodal molecules that were not associated with any observed demographic or clinical factor are shown. The vertical red lines indicate the estimated distribution modes. Such unexplained bimodal examples (“missing variability”) were seen in all four omes.

Using a conservative bimodal index threshold of > 1.3 and after removing any potential technical artifacts, 55 molecules were considered potentially bimodal, including 2.0% of lipids, 2.2% of proteins, 4.5% of metabolites, and 8.1% of cytokines (Supplementary Table *8*). The molecules with the largest bimodal indices were manually plotted, revealing a number of distinctly multimodal distributions, with examples from each ome (Figure *3*). To rule out demographic or clinical explanations for these distributions, we checked each bimodal molecule for associations with age, sex, BMI, race/ethnicity, diabetic status, and any of the clinical tests. In some rare cases, these associations appeared sufficient to explain the bimodality of the molecule. For example, in the case of granulocyte-macrophage colony-stimulating factor (GM-CSF; cytokine27; BI = 1.52), sex and BMI were both strongly associated with GM-CSF levels, with demographic factors accounting for 75.4% of the variance in the molecule. Visually, stratifying by sex alone replicated the bimodality of the underlying distribution. Of the 55 putatively bimodal molecules, 25 had one or more demographic or clinical associations and were thus considered to have their distributions at least partly explained by known factors, even if the variance explained was relatively minor.

Some bimodal patterns also appeared to be partially explained by known QTLs. Among the 11 analytes with previously reported QTLs, three QTLs showed significant associations in the iPOP cohort: SERPINA5 (protein127), N-methyllysine (metabolite52), and complement factor B (CFB; protein21). For example, the SERPINA5 protein has a known pQTL (rs2731674), and this variant risk allele dosage was significantly associated with SERPINA5 protein levels (P = 1.4 x 10^-6^) (Figure *3*). After adjustment for this genotype dosage, the bimodal index decreased from 1.93 to 1.36, suggesting that the observed bimodality was partly (but not completely) driven by genetic variation (Supplementary Figure *15*, Supplementary Table 9, Supplementary Table 10).

Other bimodal molecules appeared to be related to dietary patterns. The metabolite 2-piperidinone (metabolite365; P = 5.72e-5), for example, was associated with the use of sugar in tea in iPOP, indicating a potential relationship of 2-piperidinone with tea, which has been noted before^18^. Similarly, the metabolite lysophosphatidylcholine 20:4 (metabolite211; P = 0.0030) was associated with sugar use in coffee, indicating a potential relationship with coffee consumption. The lipid phosphatidylethanolamine (P-16:0/22:6) was associated with the consumption of oil-rich fish and white fish (lipid296; P = 6.89e-6), with greater fish consumption associated with higher levels of the lipid. A group of triacylglyceride lipids (lipid630, lipid722, lipid796, lipid538) were associated with cake consumption (P = 0.0013-0.0022).

The remaining unexplained bimodal molecules included strikingly bimodal distributions across all four of the blood omic types (metabolite259, quinic acid; protein125, HV270; lipid149, lysophoshpatidylethanolamine 18:2; cytokine56, IL22) (Figure *3*). Both pathway enrichment and GSEA analyses identified only one enriched annotation among the full set of bimodal analytes, which was xenobiotic metabolites (GSEA q value = 0.037).

We then assessed whether the bimodality of these molecules replicated in the hPOP cohort. Of the 30 unexplained bimodal molecules, 10 were also measured in the hPOP cohort. Despite the differences in platforms and populations, we found strong evidence that two analytes were bimodal: plasma levels of the protein SERPINA5 (BI = 1.87) and plasma levels of the metabolite quinic acid (BI = 1.33 and 1.56 for the untargeted platform).

### Cross-sectional variation analysis

For each of the blood omes, the interindividual (i.e., “cross-sectional”) CVs were calculated for each molecule, providing a measure of variability across the population (Figure *4*). Lipids had a much lower mean interindividual CV (15.5%) than proteins (56.8%), metabolites (65.2%), and cytokines (56.0%) (Supplementary Figure *16*). Among the lipids, TAGs and DAGs had substantially higher CVs (19.4% and 13.9%, respectively) than the other lipid classes (Supplementary Table *11*). Among metabolites, two super pathways had much higher CVs compared to the others: xenobiotics (100.7%) and cofactors and vitamins (81.3%). Individual CV outliers were also common, particularly among the lipid super pathway (Supplementary Table *12*). Among the metabolite subpathways, progestin steroids were the most variable (198.1%), followed by primary bile acid metabolism (146.9%) and food component/plant metabolites (131.0%) (Supplementary Table *13*).

**Figure 4.**
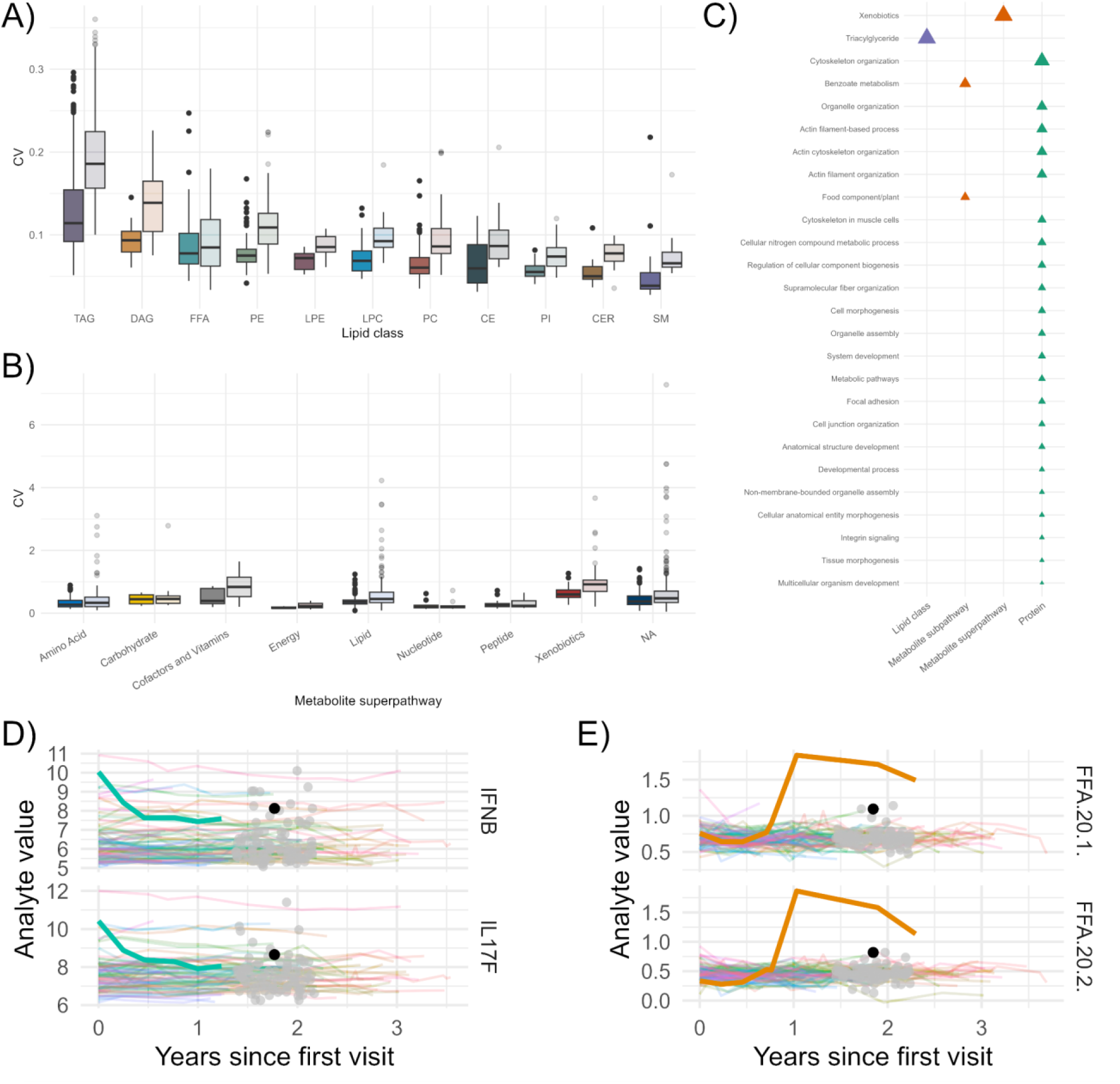
Cross-sectional and longitudinal molecular variation. **A)** The coefficients of variation (CV) distributions for lipid species are shown, stratified by lipid class. Both the mean intraindividual CVs (calculated across all visits per participant, then the mean across participants was taken per molecule) and the interindividual CVs (cross-sectional, calculated across median per-participant lipid measurements) are shown. Intraindividual CVs are in the darker color while the interindividual CVs are in the lighter color. For nearly all classes, the interindividual CV was substantially larger than the intraindividual CV, except in the case of free fatty acids (FFA). **B)** The interindividual (lighter color) and intraindividual (darker color) CVs for metabolites are shown, stratified by metabolite superpathway. **C)** Enriched pathways among molecules with high (up arrow) or low (down arrow) intraindividual CVs from a gene-set enrichment analysis (GSEA) are shown. Greater longitudinal variability was seen for xenobiotics, triacylglycerides, cytoskeleton structure-related proteins, and benzoate metabolites. **D)** An example of a longitudinal trajectory that could be easily missed in a cross-sectional analysis is shown. The levels of two different cytokines are plotted over time (colored lines, by participant). The median values per-participant are shown in the center as gray points. One participant (darker lines and point) started with relatively high levels of two cytokines (interferon beta and interleukin 17F), but gradually returned to more population-average values over 6 months. **E)** Another example of a longitudinal trajectory that was poorly captured by cross-sectional data, this time from lipid levels. Here, one individual’s levels of two free fatty acids (20:1 and 20:2) underwent an upward shift after about 9 months and remained elevated over the next year. The elevated state was >8 standard deviations above the rest of the population, a difference that was muted when viewed cross-sectionally.

### Longitudinal variation analysis

The intraindividual CVs (i.e., the CV in one molecule’s values for one person over time, also called the “longitudinal CV”) differed from the interindividual CVs in some ways (Figure *4*, Supplementary Figure *17*). As with the interindividual CVs, the intraindividual CVs were generally lowest among the lipids (mean intraindividual CV = 10.5%) and higher among the metabolites (40.7%) and proteins (50.3%). However, there was much less intraindividual variability among cytokines (18.2%) than interindividual variability (56.0%).

The GSEA analysis identified several enriched annotations among the molecules with the greatest intraindividual variation (Figure *4*) (Supplementary Table *14*). Similar to the interindividual CV analysis, xenobiotic metabolites and TAG lipids tended to vary the most longitudinally among participants (GSEA q values of 7.0 x 10^-10^ and 1.0 x 10^-9^, respectively). Unlike the interindividual CV analysis, however, metabolites related to benzoate metabolism, metabolites related to food component/plant pathways, and proteins related to the cytoskeleton, muscle, and other pathways tended to vary the most, potentially reflecting how diet and exercise tend to be dynamic for participants over time.

The inter- and intraindividual CVs were then compared by molecular subtype. Among lipid classes, the interindividual CVs were generally higher than the intraindividual CVs, with one major exception: FFAs tended to be just as variable over time within individuals as across individuals (Figure *4*; Supplementary Table *15*). Among metabolite superpathways, intraindividual CVs were generally lower than interindividual CVs for xenobiotic, lipid, energy, and cofactor and vitamin metabolites. For the remaining superpathways, the inter- and intraindividual CVs were more similar (Supplementary Table *16*). Among metabolite subpathways, there was a large difference observed for progestin steroids (namely, pregnanediol-3-glucuronide and pregnanolone sulfate): the mean interindividual CV for these molecules was 198.1%, while the mean intraindividual CV was 34.8%, indicating far more variability across participants than within a participant over time and perhaps reflecting strong sex-specific effects for this molecule (Supplementary Table *17*).

A systematic analysis of molecule-specific inter- and intraindividual CVs was then performed. First, the interindividual CV of each molecule was plotted against its mean intraindividual CV (Supplementary Figure *18*). As expected, the interindividual CV was higher than the mean intraindividual CV for 78.2% of all metabolites, lipids, proteins, and cytokines, reflecting the fact that molecular differences between individuals are typically greater than individual molecular changes over time. Several exemplars of this pattern were identified, including interferon-beta (cytokine17), prenanediol-3-glucuronide (metabolite331), and IGHD (protein75) (Supplementary Figure *19*; Supplementary Figure *20*; Supplementary Figure *21*).

### Individual molecular hypervariation

However, these analyses focused on the mean intraindividual CVs, which average over all individuals in our data set and thus may obscure relevant personalized molecular trajectories. To identify cases in which someone’s personal molecular trajectory was exceedingly different from the rest of the population, we analyzed each person’s intraindividual CV for each molecule against the mean intraindividual CV for that molecule. Because CV-based summaries do not capture timing, shape of trajectory, or the stability of estimates in participants with few visits, we interpreted these results as an exploratory prioritization strategy for identifying notable longitudinal patterns rather than as a definitive biological classification. Here, we focused on the most extreme hypervariant individuals: examples where an individual’s level of variation for a molecule was greater than 4 SDs higher than the population mean. There were 364 such individual-level molecular outliers in our cohort.

One notable example involved two cytokines (IL17F and IFNB; cytokine1 and cytokine17) for an individual whose mean levels of these molecules were only 1.08 and 1.65 SDs above the rest of the cohort, a difference that would likely go unnoticed in a cross-sectional analysis (Figure *4*). However, their intraindividual CVs for these molecules were 7.33 and 7.47 SDs higher than those of their peers. When the full trajectories were plotted for this individual against the population, a distinctive pattern was seen where this individual’s cytokine levels dropped markedly over the first 6 months of the study before appearing to level out at a slightly-higher-than-average level. In comparison, most other members of the cohort had more stable levels over time, even if those levels were on the higher end of the distribution. In this case, the personal trajectory revealed a unique feature beyond the cross-sectional mean.

Another notable example involved two free fatty acids (FFA 20.1 and FFA 20.2; lipid115 and lipid116) for an individual (Figure *4*). Here, their levels for these lipids during the first 9 months of their participation were unremarkable compared to the rest of the cohort. However, at around the 9-month mark, their lipid levels shifted upward and appeared to remain elevated for over a year of follow-up. While the mean values of these lipids were high compared to the rest of the cohort (3.16 and 2.74 SDs higher), the intraindividual CVs were markedly higher (8.21 and 8.67 SDs higher than the cohort).

### Molecular associations with demographic and anthropometric characteristics

We then analyzed the associations of each molecule with key demographic variables: visit age, sex, and baseline BMI. Our primary model used all available visit data in a generalized linear mixed model, with one molecule as the outcome and demographic variables as predictors, including a random intercept for the participant to account for repeated measures. Of the 2,675 omic variables tested, 630 (23.6%) had one or more FDR-significant associations (Supplementary Table *18*). Proportionally, BMI had the greatest number of omic associations (16.7% of variables associated), followed by sex (5.7%) and visit age (1.9%) (Figure *5*).

**Figure 5.**
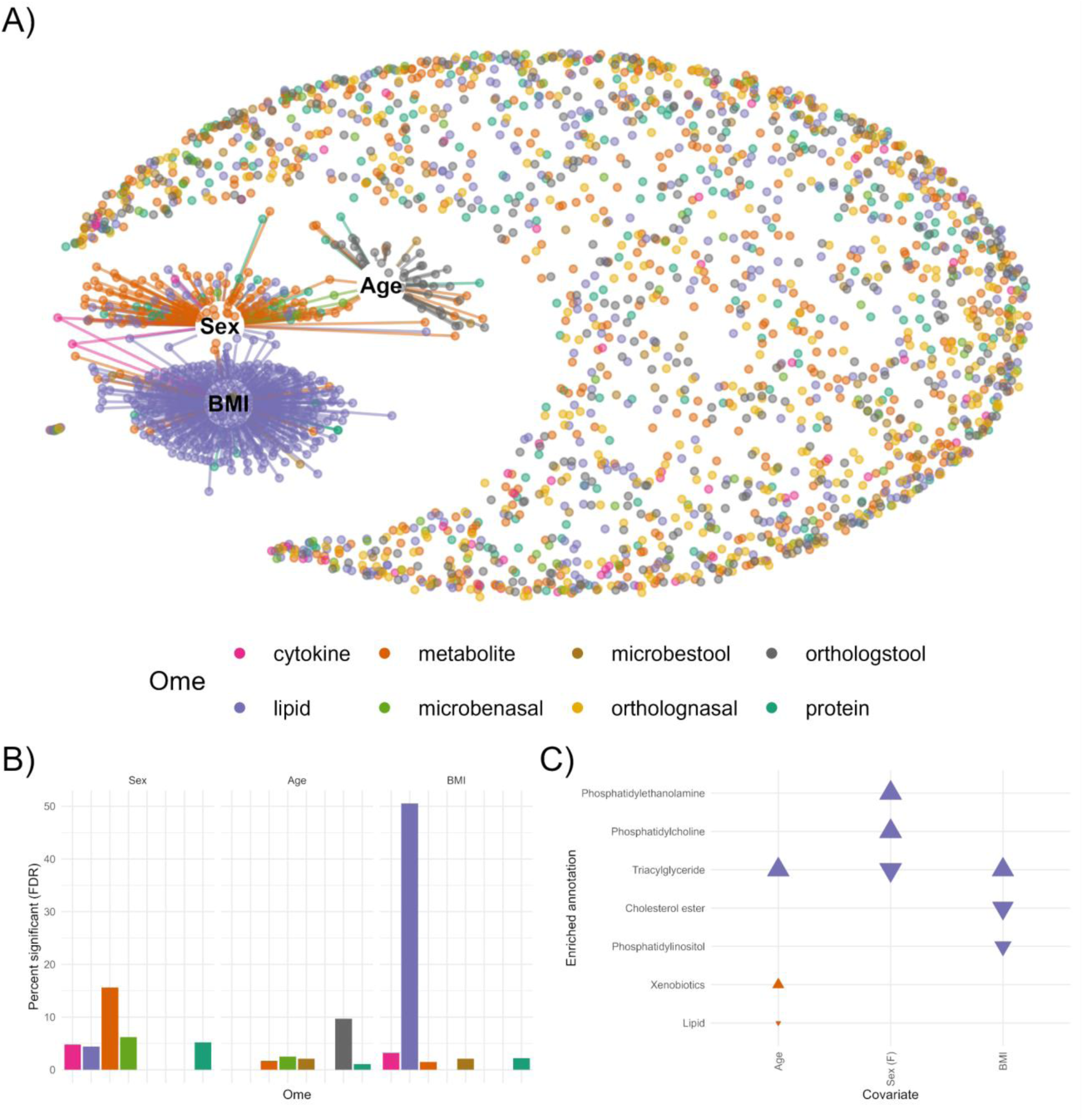
Demographic associations with the longitudinal multiome. **A)** A network visualization of omic measurements associated with sex, age, and BMI is shown. Statistically significant associations with a false discovery rate < 0.05 after multiple testing correction are shown with an edge, colored by the type of molecule associated. **B)** The percentage of molecules associated with each demographic factor (sex, age, and BMI) is shown, stratified by ome. BMI associations were dominated by lipids, whereas sex and age had more varied molecular associations. **C)** Enriched (GSEA) functional annotations among the molecules associated with each demographic are shown. An up arrow represents enrichment among positively associated molecules, while a down arrow represents enrichment among negatively associated molecules.

To examine the extent to which each molecule was explained by demographic or anthropometric measures (fixed effects in our models), we quantified the marginal R^2^ for each model. Generally speaking, the marginal R^2^s were modest, ranging from a mean marginal R^2^ of 0.055 for proteins to 0.111 for cytokines and lipids. However, for some individual molecules, the variance explained by demographics was much higher. The variables best described by demographics were the cytokines GM-CSF (marginal R^2^ = 0.689) and leptin (marginal R^2^ = 0.626), both of which were significantly associated (FDR q < 0.05) with BMI and sex and nominally associated with age (P < 0.05). The next three variables were metabolites (two androgenic steroid metabolites and acetylcarnosine) with marginal R^2^s near 0.40 that were all strongly associated with sex.

The most striking set of associations was for BMI (Figure *5*). A total of 446 different omic variables were associated with BMI, and the vast majority of these (427) were lipids. Relative to all lipids tested, just over half (50.5%) of all lipids were associated with BMI. These lipid-BMI associations were enriched the most for TAGs (simple enrichment q = 3.0 x 10^-26^) (Supplementary Table *19*; Supplementary Table *20*). A variety of other molecules were associated, including 9 metabolites, 6 proteins (HP, SERPIND1, ENO1, CFH, LBP, and APOF), 2 cytokines (leptin and GM-CSF), and 2 stool microbes (Negativicutes and Selenomonadales).

Sex had the next-highest number of associated omic variables (153), with most of these being metabolites (94), followed by lipids (37), proteins (14), nasal microbes (5), and cytokines (3) (Figure *5*). The metabolites were enriched for long-chain fatty acids (q = 5.5 x 10^-6^) while the lipids were enriched for phosphatidylcholines (q = 6.4 x 10^-19^), free fatty acids (q = 0.0043), and sphingomyelins (q = 0.0091). The top-associated variables by q value were the metabolite acetylcarnosine and the cytokine GM-CSF. Several molecules previously known to be associated with sex were replicated here, including the protein PZP (higher in females^19^), the cytokine leptin (higher in females^20^), and several metabolite derivatives of sex hormones (e.g., 5 alpha-Androstan-3 alpha, 17 beta-diol, 17-glucuronide, higher in males^21^).

The 52 age-associated variables were mostly stool orthologs (35), followed by metabolites (10), proteins (3), stool microbes (2), and nasal microbes (2) (Figure *5*). The age-associated metabolites were enriched for androgenic steroids (q = 0.002). The top-associated variables by q value were two nasal microbes (both for actinomycetales) followed by two insulin growth factor proteins, IFG2R and IGFALS.

### Omic prediction models for demographics

Recognizing the potential for complex molecular associations with high-level demographics, we built a series of machine-learning models using different omic subsets to predict participant demographics (sex, BMI, chronological age, and PhenoAge, a measure of biological age) (Supplementary Figure *22*). Our goal here was not to find an alternative way to measure demographics, but rather the following: 1) assess how well each demographic variable is captured by purely omic data sets; 2) compare different machine-learning models in terms of their effectiveness for modeling omic data; and 3) identify the top molecular features or pathways related to each demographic in an exploratory analysis. Thus, these models provide much more than predictions of demographics; they provide insight into the molecular biology connected to them across the multiome.

The baseline characteristics of the participants included in the “blood omic” and “microbial” data sets used in the machine-learning models were comparable (Supplementary Table *21*; Supplementary Table *22*). Using each ome individually as well as all omes jointly, we trained and tested a variety of machine-learning models (LASSO, ridge regression, elastic net, random forest, and XGBoost) to predict demographic characteristics (Figure *6*).

**Figure 6.**
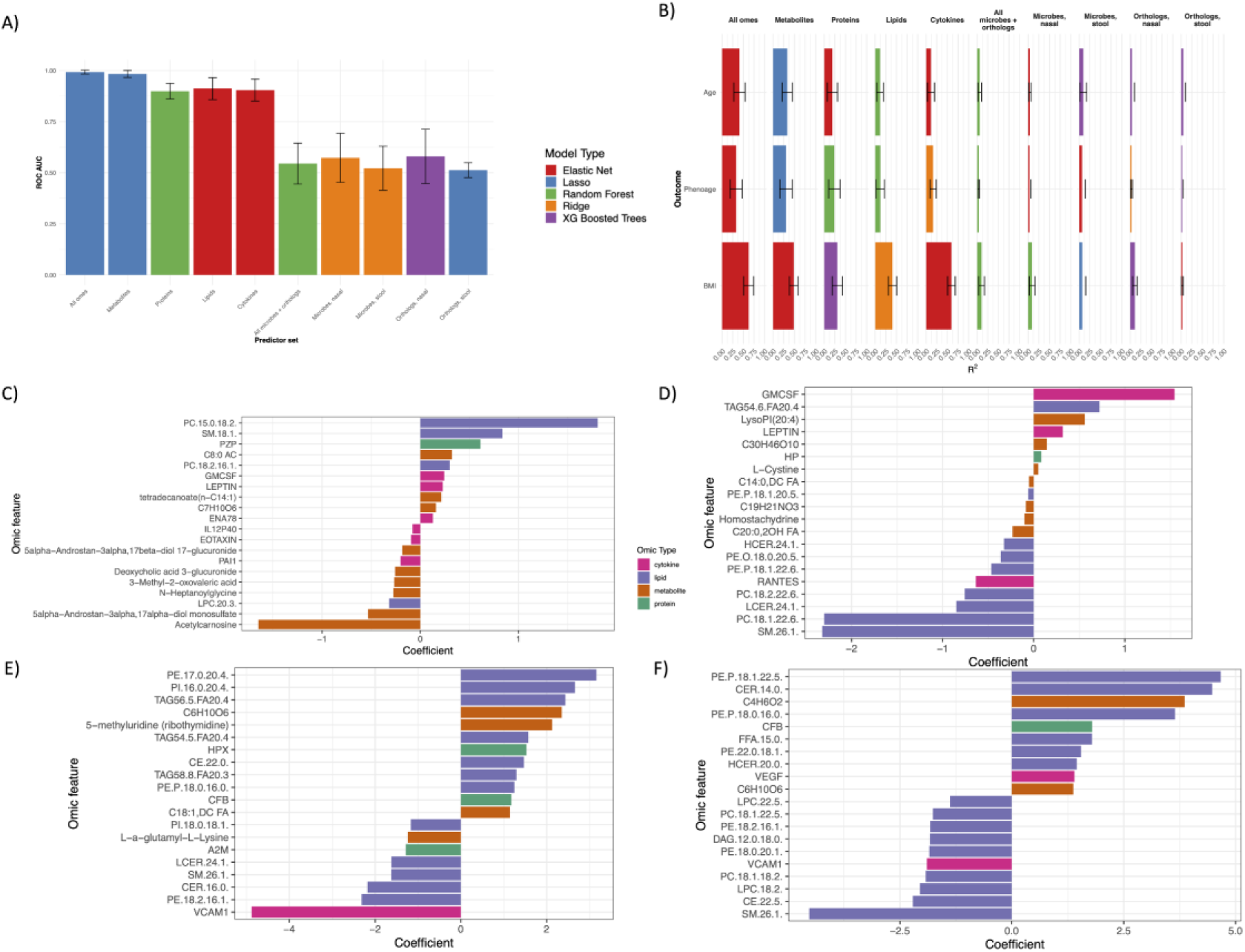
Machine-learning models of demographic variables using omic predictors. **A-B)** Bar plots displaying the performance (R^2^ or ROC AUC) of the highest performing model type (elastic net regularized regression “Elastic Net”, LASSO regularized regression “Lasso”, ridge regularized regression “Ridge”, “Random Forest”, and Gradient Boosted Trees “XGBoost”) across the 10 predictor sets (all blood omes, all microbes, or one ome in isolation) for sex **(A)** and continuous demographic outcomes **(B)** (age, PhenoAge, BMI). **C-F)** Extracted top features from the final models fit on the “all blood omes” data set to predict sex **(C)**, BMI **(D)**, age **(E)**, and PhenoAge **(F)**.

Among the demographic and anthropometric variables explored here, sex was by far the best-predicted from multiomic data (Figure *6*; Supplementary Table *23*). The average ROC AUC was up to 0.98 when using metabolite data (LASSO) and around 0.90 for at least one of the lipid-, cytokine-, or protein-based models. BMI was the next best predicted demographic. Cytokines performed the best for predicting BMI, achieving an R^2^ of 0.59 with the best model (elastic net). Metabolite models for BMI at best achieved an R^2^ of 0.48 (elastic net), followed by lipid models (R^2^ of 0.41; ridge regression) and finally protein models (R^2^ of 0.31; XGBoost). Age was the most difficult demographic to capture. For chronological age, metabolites again achieved the best prediction (best R^2^ = 0.34; LASSO), followed by proteins (R^2^ = 0.19; elastic net), lipids (R^2^ = 0.11; random forest), and cytokines (R_2_ = 0.11; elastic net).

Across all predictor sets, the microbiome consistently underperformed compared to blood ome predictors in predicting demographic or anthropomorphic characteristics (Figure *6*). To verify that this difference was not due to the greater number of observations in the “all blood omes” data set, this analysis was repeated with the “complete” data set wherein all samples had all omic data available (n = 295), thus ensuring all omic models were using the same sample size. The performance ranges for all models across the blood omic and microbial predictors are summarized in Table *1*. As expected, nearly all models had decreased performance when the “complete” data set was used, likely because of the smaller sample size. However, even after controlling for the number of observations, models trained on microbial variables severely underperformed compared to those trained on blood omic variables.

**Table 1.**
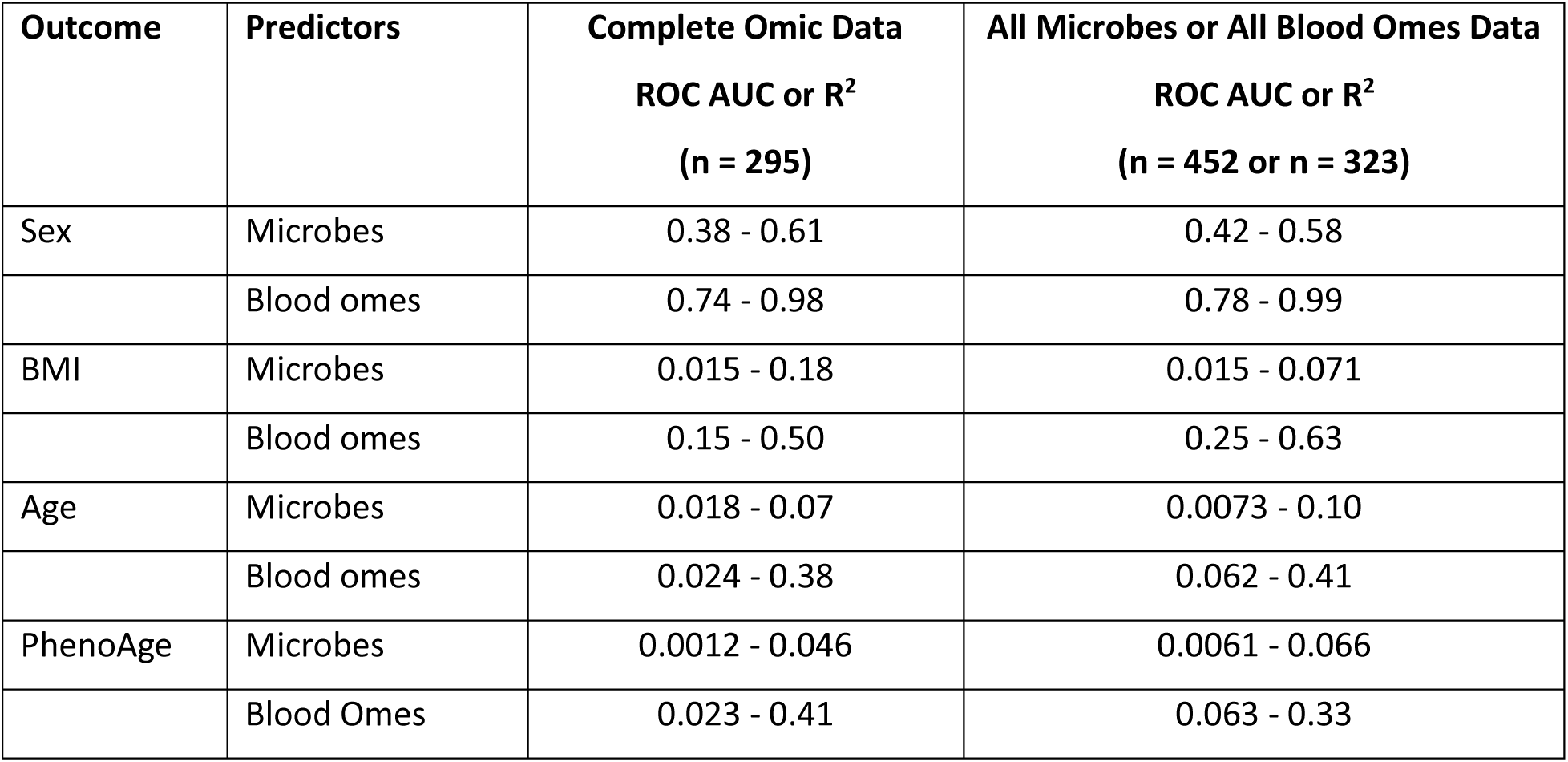
Predictive performance of models for demographic outcomes. The ranges are inclusive of every predictor set (individual blood ome or all blood omes) for the microbial or blood omic variables in the “complete” data set (n = 295) compared to the “all microbes” (n = 323) or “all blood omes” (n = 452).

### Top predictive features of demographic models

Next, we sought to fit our best-performing model type for each outcome (as determined by the initial test on the full training set using all blood omic predictors) to extract the top features for these models in an exploratory analysis (Figure *6*; Supplementary Table *24*; Supplementary Table *25*). In general, the top features of the demographic outcomes tended to be metabolomic and lipidomic features. In the case of BMI, the top features were almost exclusively cytokines and lipids, which is to be expected as BMI is known to be correlated with inflammatory markers and blood lipid content. For sex, metabolites of androgenic hormones, which are known biomarkers of sex, were among the top features. The top features for age harbored the highest level of diversity in omic type (Figure 6).

### PhenoAge prediction

As an alternative to modeling chronological age directly, we calculated PhenoAge and modeled it using the same procedure as for the other demographics (Figure *6*). We found that our PhenoAge model had nearly the same predictive power as our aging model and shared many of the same lipidomic and metabolomic top features, albeit with higher coefficients in our PhenoAge model for select metabolomic predictors.

### Sensitivity analyses for demographic prediction models

As previously mentioned, the performance metrics improved for nearly all models as a result of the larger training and test sets when the “all blood omes” data set (n = 452) was used compared to using the data set limited to samples with all blood and all microbe data available (n = 295). However, the exception to this trend was that the performance decreased when metabolite data were used to predict age or PhenoAge (Supplementary Figure 23, Supplementary Figure 24, Supplementary Figure 25).

To investigate this counterintuitive loss of performance, both the “complete” data set and the “all blood ome” data set were compared in terms of the distribution of the number of visits per subject. This analysis revealed one outlier participant who was overrepresented in the “all blood omes” data set (29 visits vs. a mean of 5 visits). Partitions with this subject in the held-out test data set incurred the largest drops in model performance. To further investigate the impact of this particular participant, the final model was re-fit on a subset of the “all blood omes” data set that excluded those visits from this participant that were missing in the “complete” data set, which left 430 visits in the data set. However, even after downsampling this outlier participant, the same drop in performance for the metabolite-age or metabolite-PhenoAge models was seen in the “all blood omes” data set compared to the “complete” data set (Supplementary Figure 26).

### Replication in hPOP

We then reran this analysis in the hPOP cohort. In general, similar platforms in hPOP tended to perform at similar predictive levels as in iPOP, with the relative performance of different omes being similar (Supplementary Table 26). The major exception was that the Olink affinity-based proteomics platform, which was only available in hPOP, was the best predictive ome for sex, age, and BMI.

As in iPOP, most omes could predict sex accurately in the hPOP cohort, achieving a mean ROC AUC upwards of 0.99 in the multiomic model. The best predictive ome for sex in hPOP was the Olink proteomics data set with a mean ROC AUC of 0.97. However, the relative performance of the omes measured by technologies similar to those used in iPOP was similar between hPOP and iPOP: the metabolomic data performed second-best in hPOP (ROC AUC = 0.94) followed by lipidomics (ROC AUC = 0.89) and mass spectrometry-based SRM proteomics (ROC AUC = 0.86). Similarly, for age, the Olink proteomics platform was the best predictor in hPOP (mean R^2^ = 0.50), but the next best omes followed the same order as in iPOP: metabolomics (mean R^2^ = 0.40), SRM proteomics (mean R^2^ = 0.38), and then lipidomics (mean R^2^ = 0.12). For BMI, the best model in hPOP was again the Olink proteomics platform (mean R^2^ = 0.39), followed by SRM proteomics, metabolomics, and lipidomics, which had similar performances (mean R^2^ = 0.20, 0.19, and 0.17, respectively).

### Omic prediction models for clinical tests

In a similar fashion to how we trained models to predict demographics, we trained and tested a variety of machine-learning models to predict 49 common clinical tests (Figure *7*; Supplementary Table *27*; Supplementary Table *28*). As before, our broad goal in building predictive models for existing clinical tests was more about what these models could teach us about multiomic data: 1) to what extent can validated health metrics (as represented by standard clinical tests) be captured by rising multiomic technology; 2) which model types work best for translating multiomic data to clinical outcomes; and 3) which specific molecules or pathways are most relevant to different clinical tests.

**Figure 7.**
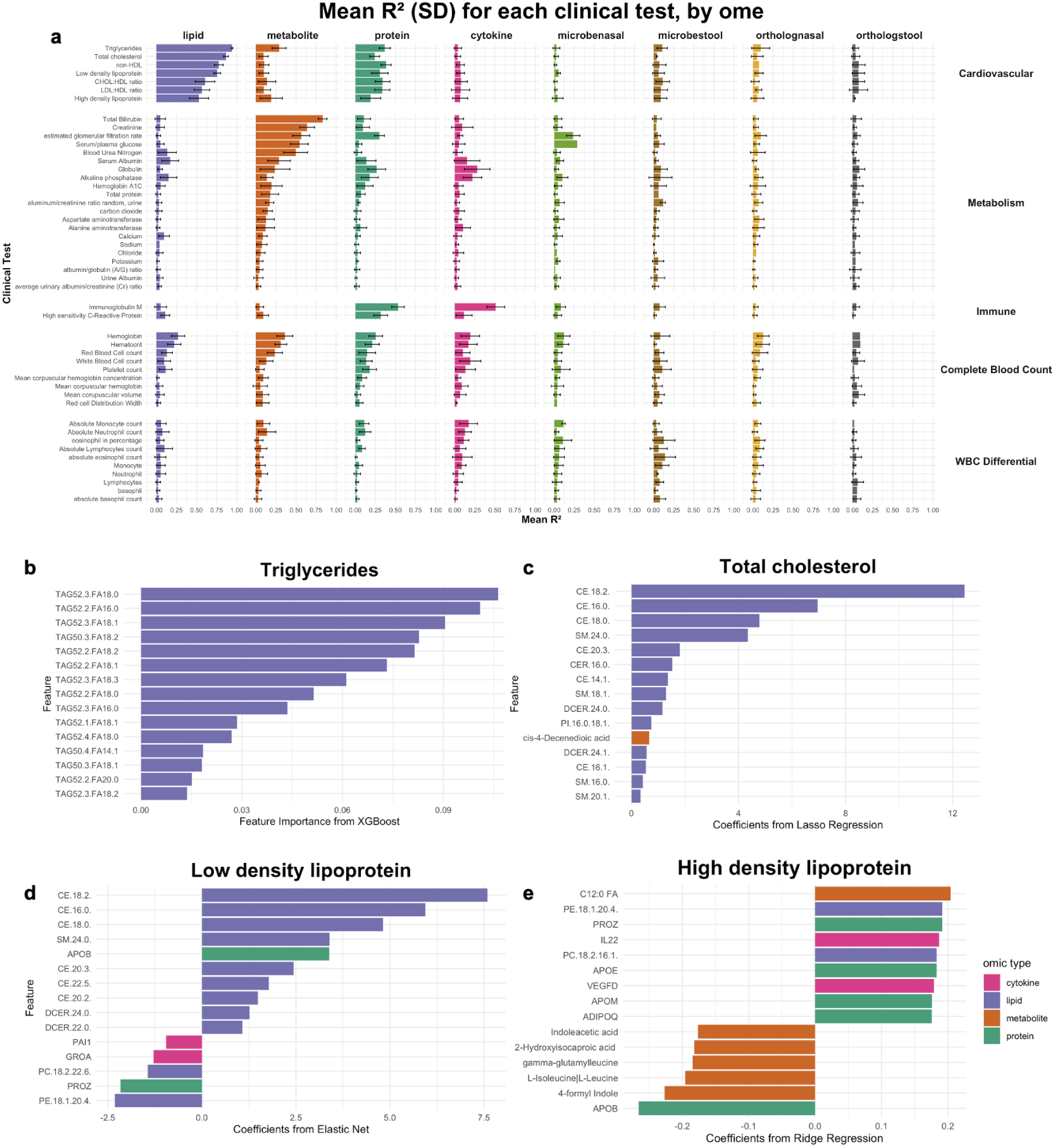
Machine-learning models for clinical laboratory tests and their top blood omic predictors. **A)** The predictive performance (mean R^2^ and standard deviation [SD]) of each omic predictor set is shown for each clinical test. In general, the lipidome, metabolome, and proteome predicted a variety of clinical tests well, whereas the microbiome (microbenasal, microbestool, ortholognasal, orthologstool) had poor predictive performance. Cardiovascular health markers, such as triglycerides (R^2^ = 0.94 [0.01]) and total cholesterol (R^2^ = 0.87 [0.03]), were among the best-predicted clinical tests. **B)** The top blood omic features predicting triglycerides are shown, which were predominantly lipid species (triacylglycerols). **C)** The top blood omic features predicting total cholesterol included cholesterol esters (CE 18.2, CE 16.0) and ceramides (CER 16.0, DCER 24.1), which are emerging cardiovascular risk factors. **D)** LDL cholesterol was predicted by APOB and several lipid species. **E)** HDL cholesterol was predicted by a greater variety of omic types, including lipids, cytokines (namely, interleukin-22), and proteins, including adiponectin (ADIPOQ), an anti-inflammatory hormone secreted by adipose tissue.

Cardiovascular health markers—triglycerides (R^2^ = 0.94 [0.01] with XGBoost) and total cholesterol (R^2^ = 0.87 [0.03] with LASSO)—were the best-predicted clinical tests. These cardiovascular markers, along with others, including non-HDL cholesterol, LDL cholesterol, the CHOL:HDL ratio, the LDL:HDL ratio, and HDL cholesterol, were well-predicted by the lipidome (R^2^ range = 0.53-0.94) (Figure *7*). Liver markers (total bilirubin R^2^ = 0.83 [0.05] with XGBoost) and metabolic markers including creatinine (R^2^ = 0.64 [0.09] with LASSO) were the next best-predicted tests, and they were best predicted by the metabolome (Figure *7*). Serum/plasma glucose was also well-predicted by the metabolome (R^2^ = 0.54 [0.11] with random forest) (Figure 7). Given these models had predictors that were analogous to the clinical analyte of interest (e.g., molecules annotated to triglycerides or creatinine), we consider these to be examples of “platform validation,” where omics platforms reproduced similar information to clinical assays (Supplementary Figure 27).

Several clinical test models generated additional biological insight: they were modestly predicted by one or more omes (mean R^2^ > 0.2) without relying on predictors that were analogous to the clinical analyte. For instance, estimated glomerular filtration rate was strongly predicted by metabolites (R^2^ = 0.56 [0.11] with XGBoost) (Figure 7), where its top features did not include creatinine (Supplementary Figure 28). Immune-related clinical tests were also well-predicted and generated additional insights. For example, immunoglobulin M was well-predicted by cytokines (R^2^ = 0.51 [0.11] with ridge regression) (Figure 7); its top features included IL2 and IL17F (Supplementary Figure 28). HsCRP was predicted by the proteome (R^2^ = 0.32 [0.09] with XGBoost), with other distinct immune and acute-phase proteins (complement component 9 [C9], lipopolysaccharide-binding protein [LBP], and serum amyloid A1 [SAA1]) as top predictors (Supplementary Figure 28). Other tests in this category can be found in Supplementary Figure 28. In these cases, omics data captured additional biological information rather than measuring the same analyte as the clinical test. Lastly, 29 clinical tests were poorly predicted across all omes (mean R^2^ <= 0.2), including some CBC tests and all white blood cell (WBC) differential tests (Figure 7).

In general, the microbiome did not predict clinical tests as well as the blood omics data (Figure *7*). To evaluate whether the smaller size of the microbiome data set explained these findings (blood omics data were more routinely collected [n = 452 visits] than microbial data [n = 323 visits]), we reran all blood omic and microbiome models on a smaller, shared data set of 295 visits where all data were available. This sensitivity analysis revealed similar findings, with the microbiome underperforming relative to the blood omics data (Supplementary Figure *29*).

Lastly, in terms of the relative performance of different machine learning models, while many machine learning models performed similarly, XGBoost models had consistent performance across omes and were often the top models, especially for the well-predicted clinical tests (Supplementary Table *27*).

### Top predictive features of clinical tests

Triglycerides and total cholesterol were primarily predicted by lipid species (particularly triglyceride species and cholesterol esters) (Figure *7*). Notably, ceramides (CER.16.0, DCER.24.0, and DCER.24.1) and sphingomyelins (e.g., SM.24.0) were also top predictors of total cholesterol, echoing studies that have increasingly recognized them as independent risk factors for cardiovascular health^22^.

In contrast to triglycerides, low density lipoprotein cholesterol (LDL-C) and high-density lipoprotein (HDL) were predicted by a greater variety of omes, including metabolites, proteins, and cytokines (Figure *7*). In addition to cholesterol esters, LDL-C was also predicted by apolipoprotein B (APOB), a component of LDL and a known predictor of cardiovascular disease^23,24^. Conversely, HDL was predicted by phosphatidylethanolamines (PE.18.1.20.4) and phosphatidylcholines (PC.18.2.16.1) and was inversely associated with APOB (Figure *7*). HDL was also associated with Interleukin-22 (IL22) and adiponectin (ADIPOQ), an anti-inflammatory hormone secreted by adipose tissue. Lastly, leptin, a metabolic hormone not routinely assessed in the clinic, was one of the top features that predicted several clinical tests (e.g., serum/plasma glucose, hsCRP, hemoglobin, hematocrit, and alkaline phosphatase). All of these tests were robustly predicted (R^2^ > 0.2 for any ome) in our original models (Supplementary Figure 27, Supplementary Figure 28, Supplementary Table 29). The coefficients and feature importance for the models predicting other clinical tests can be found in the accompanying supplementary tables (Supplementary Table *29*, Supplementary Table *30*).

### Replication in hPOP

We then sought to replicate our findings in the hPOP cohort, where 5 clinical tests were available to predict, all of which had also been analyzed in iPOP (namely, fasting glucose, fasting creatinine, high-sensitivity CRP, albumin, and total alkaline phosphatase) (Supplementary Table 31). Despite the differences in omic technologies between iPOP and hPOP, the omic trends and overall predictive performance of these models were generally consistent in hPOP. For instance, fasting glucose and fasting creatinine were both best predicted by metabolomic data sets, and the mean R^2^ was generally similar to that of iPOP (fasting glucose: mean [SD] R^2^ = 0.59 [0.16] in hPOP and R^2^ = 0.54 [0.11] in iPOP; fasting creatinine: mean [SD] R^2^ = 0.48 [0.09] in hPOP and R^2^ = 0.64 [0.09] in iPOP). The remaining three clinical tests were less well predicted (again, similar to iPOP), but the models in hPOP were less effective than those in iPOP (mean R^2^ ranged from 0.07 to 0.09 for alkaline phosphatase, albumin, and hsCRP, respectively), likely reflecting the lower sample size in hPOP.

## Discussion

In this study, we used longitudinal multiomics from a deeply profiled precision-health cohort to explore different aspects of the human multiome. We showed how distributional structure, temporal stability, and individual-specific trajectories can be analyzed to derive health-relevant insights, and we showed how multiomic data can model demographic and clinical metrics. Altogether, our results highlight several key considerations for precision health researchers (Figure *8*).

**Figure 8.**
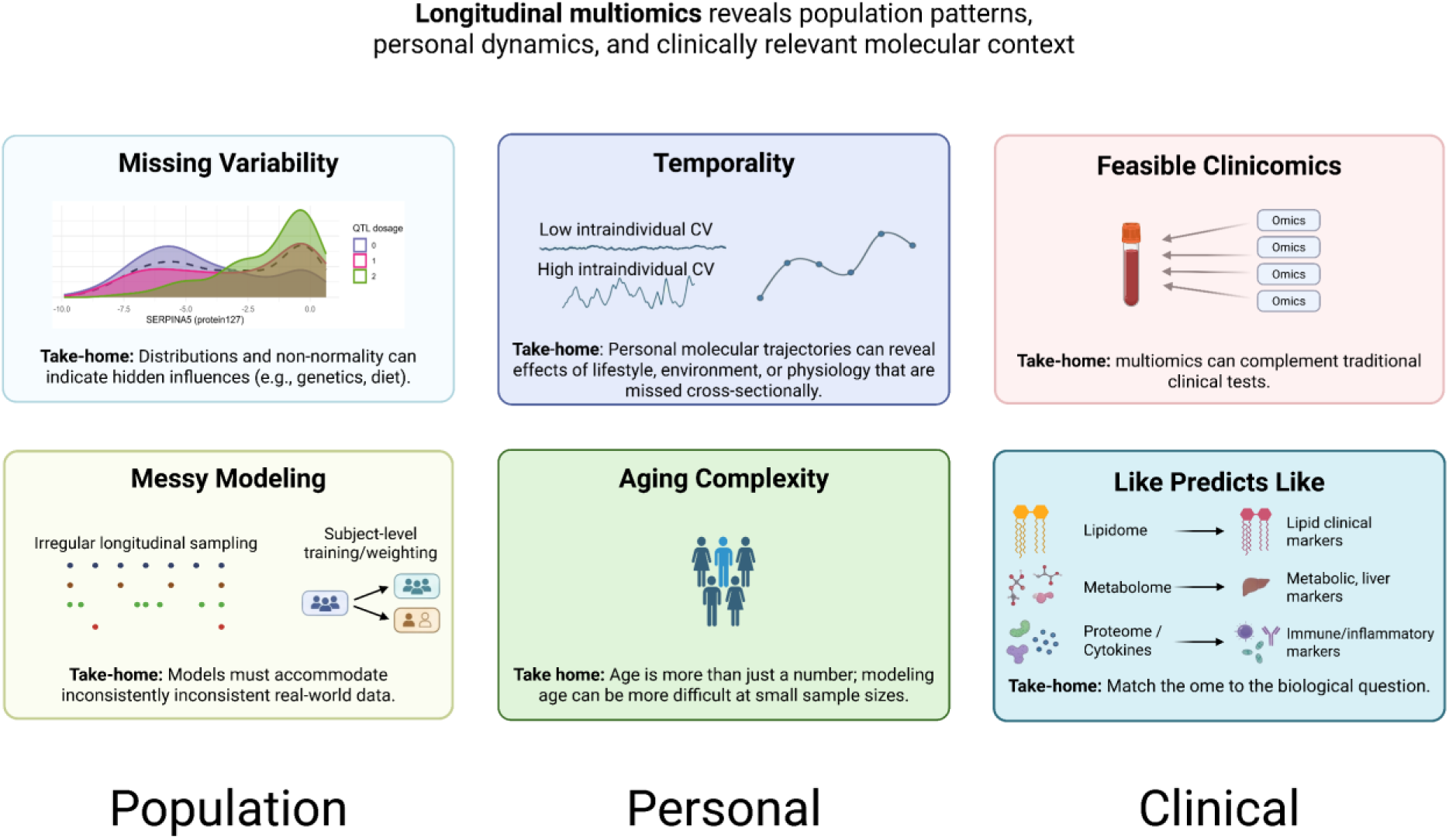
Key lessons for using longitudinal multiomics in precision health. Six practical considerations for implementing multiomic data in precision health are shared: distributions can reveal missing variability; modeling must account for longitudinality, irregularity, and outliers; temporal trends can reveal unique features; aging is complex; multiomic data can contextualize clinical tests; and omic data often best predict similar clinical domains.

### Missing variability

One of the greatest challenges in observational data analysis is accounting for different sources of variability in exposures of interest, particularly when those exposures are not directly observed. With a few exceptions (e.g., GM-CSF), most molecules here and in similar studies have had a sizable portion of “missing variability” unexplained by demographic factors^11,25–27^. We found that a simple analysis of molecular distributions can help flag molecules that are more likely to have strong genetic or environmental influences. Across the omes analyzed here, a sizeable fraction of molecules (between 2% and 8%) appeared to be non-normally distributed. Through the integration of demographic, genetic, and dietary data, we demonstrated how some of these bimodal distributions can be at least partly explained. For example, the levels of the protein SERPINA5 were shown to aggregate across our population by pQTL genotype, and the levels of four triglyceride molecules were associated with cake consumption. However, approximately half of the bimodal molecules we identified were not associated with any measured technical, demographic, clinical, or genetic factors. Such bimodality has been noted in numerous studies and contexts (Supplementary Note)^28–37^, but systematic evaluation for bimodality in omics data is relatively uncommon. Despite their potential relevance to precision health analysis, these findings stand at odds with how population omics data are typically handled: a one-size-fits-all transformation (usually logarithmic or rank-based inverse-normal transformations) is applied for convenience and because it works for most variables in a high-dimensional omic data set.

Thus, we make two recommendations for future multiomics research. First, population omics researchers should consider explicitly testing for non-normality and, where possible, exploring potential drivers of non-normally distributed molecules. While the majority of molecules do appear normally distributed, the non-normal exceptions may provide clues that technical, genetic, or lifestyle factors may be confounding the data, even in the absence of direct measurement of those factors. Testing for bimodality may be an easy way to assess for potential confounding and identify results that merit extra caution in interpretation. Second, there is a need for methodologies that bridge the cellular level (where bimodalities have been found in response to cellular states) to the organismal level (where bimodalities are often not tested). Methods or pipelines to systematically identify these molecules and test for potential explanations may be invaluable to observational multiomic studies where environmental confounding of molecular data is likely to be commonplace.

### Temporality

With this longitudinal multiomic data set, we found important differences in interindividual (across the population) and intraindividual (over time for an individual) variation. As expected, interindividual variation was generally greater than intraindividual variation, but this difference was starkest for cytokines, xenobiotics, lipid, energy, and cofactor and vitamin metabolites. Other studies have reported similar trends, for example, in longitudinal proteomics from the SCAPIS study^26^. In contrast, some molecules, like TAGs, tend to vary as much over time for a given individual as they do across a population. The extent to which a given molecule tends to be temporally stable (i.e., has a low intraindividual CV) affects how informative a cross-sectional study of that molecule can be. As we wait for more longitudinal omics from population biobanks, information about molecules with low vs. high intraindividual variation should be considered when interpreting molecular associations from cross-sectional analyses. Moreover, on the individual level, a shift in molecules that otherwise tend to be stable (e.g., here, a participant had IFN-beta and IL-17F cytokine levels that gradually shifted over 6 months) can indicate clinically relevant changes. Such temporal shifts may easily be missed in cross-sectional cuts or aggregations of data, but they represent a useful feature for precision health.

### Feasible clinicomics

In recent years, there has been a push to use omics data where current clinical tests are invasive, expensive, or otherwise infeasible for routine use. For example, research has shown that omics and wearable data can be useful for predicting measures of insulin resistance^38^. A prior study in iPOP likewise showed that multiomic data, in addition to demographics, was able to capture a substantial portion of the variance for the steady-state plasma glucose test (SSPG; R^2^ = 0.87) and the oral glucose tolerance test (OGGT; R^2^ = 0.71)^39^, which are highly informative but laborious tests to perform that take multiple hours to complete. Our findings scale up this multiomic prediction effort to 49 common clinical tests. The lipidome had the highest predictive power for many cardiovascular markers, including triglycerides (R^2^ = 0.94) and total cholesterol (R^2^ = 0.87). The metabolome and proteome were the next most predictive omes, with the metabolome predicting liver and metabolic markers and the proteome predicting immune and inflammatory markers. In contrast, the nasal and stool microbiomes and orthologs had poor predictive performance (R^2^ < 0.2) for nearly all clinical laboratory tests.

We classified our clinical test predictive models into three categories: 1) platform validation, 2) potential clinical utility, or 3) limited clinical utility. We considered our results to show platform validation when an omic platform included the target of the clinical test. Here, 10 clinical tests were well-captured by omics technology (R^2^ > 0.5) while including omic measurements that matched the clinical test (e.g., predicting creatinine from a clinical test with creatinine measured in the metabolomics data set). Secondly, 10 clinical test models (20%) showed potential clinical utility: they were modestly predicted by one or more omes (mean R^2^ > 0.2) without omic predictors that matched the clinical analyte. Lastly, 29 clinical tests were not well-predicted by any ome (R^2^ <= 0.2), particularly several CBC tests and WBC differential tests, thus indicating limited clinical utility. This poor performance was not unexpected given our omics platforms measured bulk samples rather than cellular composition.

The top features of our best predictive models, while considered exploratory, highlighted several emerging biomarkers that have been associated with traditional clinical laboratory tests. Ceramides and sphingomyelins, emerging risk factors for CVD, were the top predictors of total cholesterol. Ceramides, especially CER.16.0 and CER.24.1, have been recognized as independent drivers of CVD^22,40^ and can predict CVD mortality in patients with coronary artery disease better than LDL-C alone^41^. Our findings shed light on the biology associated with clinical tests, reinforce the utility of “non-traditional” biomarkers, and suggest new avenues for exploration. Overall, these findings suggest that high-dimensional omics data may help contextualize, complement, or even one day supplant some clinical laboratory measurements in the future, particularly as technological development helps improve the standardization and quantification of omics data.

### Like predicts like

A common question facing many investigators is which omic data sets to invest in for a given cohort. While no simple answer exists to this question, our findings provide high-level insight into differential omic utility. Generally speaking, we found that intraomic relationships were stronger than interomic, and our predictive models reinforced the idea that “like predicts like”. The greatest number of associations with BMI was found among the lipidome. Similarly, lipids were the best predictors for lipid-related outcomes like LDL-C and cholesterol. To some extent, we expect this trend to be due to platform effects: it makes sense that molecules measured by the same platform at the same time would correlate more than molecules measured in a different manner. In prior studies, we have seen similar trends when multiple omics were compared as predictors of some outcome of interest. For example, in studying Alzheimer’s disease, where two primary biomarkers (amyloid and tau) are both proteins, the cerebrospinal fluid proteome was better than the metabolome or genome in predicting these biomarkers^42^. While important and different biological signals may be found in different omic data, researchers interested in prediction may want to consider matching the ome to the outcome to the extent possible.

### Age is more than just a number

One of the greatest and most common risk factors across all diseases is aging^43^. With the advent of multiomics data, researchers have identified many associations between age and molecular data, like proteomics, metabolomics, transcriptomics, and more^12,44–46^. However, despite the number of these known associations, we found that predicting age was much more challenging than predicting other demographic features. When all four blood omes (lipids, cytokines, metabolites, and proteins) were used simultaneously as predictors, the best model achieved an average R^2^ of 0.41, lagging behind the predictions of BMI (best R^2^ of 0.63) and sex (ROC AUC of 0.99). While much better age prediction is possible when using larger data sets^47^, we note that the relative challenge of predicting age compared to other demographics at a given sample size underscores the complexity of human aging. Furthermore, we observed that models of age using metabolites were inconsistent in their performance depending on the data set we used, which perhaps reflected a small sample size relative to the complexity of aging metabolic processes^12^. Some of the top predictors for age here overlapped with known associations (e.g., branched-chain amino acids and VCAM1), although the direction of effect did not always align with prior studies (Supplementary Note).

### Messy modeling

A major area of growth in biomedical research has recently come from the use of machine learning and AI methodologies, which can capture complex interdependencies among high-dimensional data that may be harder to model with more traditional statistical methods^48,49^. Most models focus on cross-sectional data, where each participant is measured once. However, as omic data sets expand longitudinally, we will need a stronger modeling infrastructure that can handle high-dimensional data measured over time, where samples are inconsistently inconsistent in structure with varying numbers of visits, time between visits, and total duration of observation. Here, we used a participant-based training scheme that weighted observations by the inverse of the number of visits for each participant. For most outcomes and omic data sets, this framework performed well. However, we did see an issue in model performance due to the presence of one highly sampled individual whose aging metabolic profile appeared to be unusual enough as to skew the model’s training and performance. In contrast to efforts to understand population-level trends in health and disease, precision medicine must accommodate such outliers, because exceptional individual differences may very well represent important health information. Much progress has already been made in adapting machine-learning and AI methodologies to longitudinal data (e.g., random forest^50^ and neural networks^51^), which will be key to precision health applications in the future.

This study had several limitations. First, our sample was limited to the geographical region of the San Francisco Bay Area, which limits the generalizability of our results to populations not well-represented there. A related potential issue is that iPOP was originally designed to focus on diabetes and prediabetes and thus was enriched for these phenotypes. However, prediabetes is common in the United States (43.5% of U.S. adults are estimated to be prediabetic^52^), and the prediabetes prevalence in iPOP (56.4%) was not far off from this national average. Another related question is how to control for confounders in observational data analysis. We controlled for diabetes status in our longitudinal regression analyses to help remove associations or bias driven by diabetes status, but for some molecules, controlling for diabetes may obscure real associations or create false positives, depending on the underlying causal relationships. As is the case for observational multiomic analysis in general, our novel associations for molecules that may be related to diabetes would need appropriate replication to determine their causality. Second, most of our omics data were generated in-house with mass spectrometry-based methods. Increasingly, population biobanks are turning to commercial services to generate large-scale proteomics and metabolomics data^4–10^. Different platforms often correlate poorly with each other, particularly for proteomics^53–55^, which may limit the generalizability of some of our findings when different platforms are used. Still, many of our findings were replicated in the hPOP cohort, where different omic technologies and platforms were used (e.g., Olink vs. mass-spectrometry proteomics and targeted vs. untargeted metabolomics), which supports the generality of some of the omic trends shown here. Also, different data quality control steps, imputation, and transformation choices could impact some of the analyses shown here (e.g., the identification of bimodal distributions). Thus, the use of these methods should be considered an exploratory first step to identify putative bimodal distributions, with targeted replication in independent data sets and with different omics platforms an important follow-up step (as we did here with replication in hPOP and with genetic and dietary data). Finally, while our cohort is one of the largest longitudinal, observational, and multiomic cohorts available, the sample size of 526 visits from 94 participants is still relatively small compared to the scope of cross-sectional cohorts, which are now in the hundreds of thousands. While we were able to replicate many of our cross-sectional findings in the hPOP cohort, we were unable to test some of our other insights due to a lack of similar longitudinal multiomic data. Thus, our findings should be viewed as generating early insights into the analytical questions that will likely be at the forefront of precision health as population biobanks increasingly expand longitudinally^56^.

The goal of precision medicine is lofty, but new molecular technologies are bringing us closer to achieving it. As our molecular understanding of diseases and how to treat them grows, so too must our understanding of health across the breadth of populations and the depth of biology. Real-world observational, multiomic data sets are growing rapidly, opening up tremendous opportunities to understand how an individual compares—on a molecular level—to the population. Our results show the immense potential of high-dimensional molecular data for precision health, from population-level omic patterns, to personal molecular trajectories, to demographic and clinical modeling. As our technologies and data continue to evolve, we look forward to ever deeper insights into the rich molecular landscape of humanity.

## Methods

### iPOP cohort

This study was conducted using data from the iPOP cohort, which has been previously described^39,57–60^. Briefly, the cohort includes 109 individuals recruited primarily from the San Francisco Bay Area. The cohort is unique in its combination of breadth, depth, and sample size of molecular profiling. The cohort is longitudinal and ongoing (data used here were collected between 2010 and 2017), with participants returning approximately every 3 months for sample collection (blood, saliva, and stool) at quarterly visits. All blood samples were collected after an overnight fast. Additional visits were also conducted for various reasons, including when participants developed an illness, notable stress, recent travel, etc. Here, however, we focused only on the visits that were considered “healthy”; i.e., there were no other unusual circumstances documented for the visit. At each visit, a full suite of multiomic data was generated: proteomics, cytokines, metabolomics, lipidomics, stool microbiome, nasal microbiome, a comprehensive battery of routine clinical tests, and more. Baseline survey data (e.g., lifestyle, diet) and genome-wide genotyping were also collected.

After enrollment in the study, each participant also underwent a number of glucose and insulin tests^39^. Each participant was classified as normoglycemic, prediabetic, or diabetic based on various measures of metabolic health (e.g., patient report, hemoglobin A1c [HbA1C] fasting glucose, glucose tolerance test, etc.). While the cohort was enriched for individuals with prediabetes or diabetes, the cohort otherwise represented a relatively healthy sample of the Bay Area population. The diabetic status of participants was controlled for, where possible, in subsequent analyses.

### Proteomics

Untargeted proteomics data were generated from blood plasma samples through sequential window acquisition of all theoretical (SWATH) mass spectrometry, as described in detail in previous iPOP work^39,58^. In brief, the iPOP samples were randomized and then analyzed on a TripleTOF 6600 System with a DuoSpray Source and 25 μm I.D. electrode (SCIEX). The sum of the top three most abundant peptides were summed to estimate protein abundances. Batch effects were removed with Perseus (v. 1.4.2.40) by subtracting the principal components showing a major batch bias. In cases where multiple variables were annotated to the same protein, all such variables were removed. Proteins were annotated with the corresponding gene symbol, Entrez ID (Gene ID), and Uniprot ID using the biomaRt, mygene, and org.Hs.eg.db R packages, with additional manual annotation as needed.

### Lipidomics

The generation of lipidomics data for the iPOP cohort is reported in detail in a previous manuscript^59^. Briefly, lipids were characterized in iPOP blood plasma samples using mass spectrometry (MS). Lipidyzer, a high-throughput quantitative lipidomics instrument comprising a triple-quadrupole MS (Sciex QTRAP 5500) and a differential mobility separation (DMS) device, was used to identify and quantify plasma lipid levels. Known spike-in standards were used to estimate the absolute quantities (units reported: nmol/mL) of each lipid.

After initial quality control, 846 lipid species were quantified in our healthy subcohort across various lipid subclasses: free fatty acids (FFA), triacylglycerols (TAG), diacylglycerols (DAG), cholesterol esters (CE), phosphatidylcholines (PC), lysophosphatidylcholines (LPC), phosphatidylethanolamines (PE), alkyl ether substituent containing PEs (PE-O), alkenyl ether (plasmalogen) substituent containing Pes (PE-P), lysophosphatidylethanolamines (LPE), sphingomyelins (SM), phosphatidylinositols (PI), ceramides (CER), hexosylceramides (HCER), lactosylceramides (LCER), and dihydroceramides (DCER). Lipid levels were log_2_-transformed before analysis. Missing values were imputed with a K-nearest-neighbor algorithm, accounting for a truncated distribution^59^.

### Metabolomics

Untargeted metabolomics data were generated from plasma samples using liquid chromatography mass spectrometry (LC-MS), as previously described in detail^61,62^. To maximize coverage of the human metabolome, four different analyses were used to quantify metabolites: HILIC ESI (+), HILIC ESI (-), RPLC ESI (+), and RPLC ESI (-). These analyses were run on a Thermo Ultimate 3000 RSLC system with a Thermo Q Exactive plus mass spectrometer. Samples were prepared and analyzed randomly, with quality control samples used to equilibrate the LC-MS system. Metabolite features were identified and quantified by mass/charge ratio and retention time from the raw MS files using the ProteoWizard MS convert tool and XCMS (v. 1.39.4) software package. Grouping and annotation were performed with the CAMERA package (v. 1.16.0). LOESS normalization was used to account for signal drift with time. Additional batch normalization was performed for each metabolite mode and polarity combination using the ComBat method (sva package, v. 3.52.0)^63^. Features were identified against a local library and multiple public databases (e.g., HMDB, FoodB, and DrugBank). Metabolite annotations were manually reviewed for accuracy. In some cases, multiple peaks were annotated as the same metabolite. When these peaks were spread out less than or equal to 1 minute in retention time, the peak values were summed to provide a single measurement for that metabolite. When peaks were spread out greater than 1 minute of retention time, the peaks were assumed to be poorly identified, and their annotations were changed to be unknown. Phospholipid measurements in HILIC mode were removed due to potential issues with detection.

### Cytokines

Cytokine profiling of participant blood samples was performed by the Stanford Human Immune Monitoring Center, as detailed in their published protocol^64^. A 62-plex Luminex antibody-conjugated bead capture assay (Affymetrix) was used. The cytokines were additionally annotated with the corresponding gene symbol, Entrez ID (Gene ID), and Uniprot ID using the biomaRt, mygene, and org.Hs.eg.db R packages, with additional manual annotation as needed.

One sample’s cytokine data included 10 cytokines with an implausible value of 0, and many of the sample’s other cytokines were on the low end of the distributions for those cytokines in our sample, likely indicating an issue with the sample or its processing. This sample was removed from further analysis.

### Microbiome

Microbiome data were collected on various iPOP samples using the Human Microbiome Project – Core Microbiome Sampling Protocol A (https://www.hmpdacc.org/). Full details of the microbiome data collection for iPOP are available^57^, but here, we only used data from the stool and nasal samples. Metagenomic DNA was isolated from samples following the HMP Protocol # 07-001, v12.0, and the MOBIO PowerSoil DNA Extraction kit. Two main kinds of microbiome data were generated: microbial taxonomy proportions and KEGG ortholog quantities.

The taxonomic data (here called “microbe” variables) came from targeted rRNA gene (16S) amplification and sequencing. The hyper-variable regions V1-V3 of 16S were amplified from the metagenomic DNA using primers that also had an adaptor sequence for sequencing. Sample-unique barcodes were used to create amplicons, which were pooled and sequenced on an Illumina MiSeq sequencing platform. The sequencing data were processed with Illumina software, converted to operational taxonomic units (OUT) via Usearch against the GreenGenes database (May 2013 version) and assigned to taxonomies using the RDP-classifier. The result for each sample was the proportional breakdown of reads across different levels of taxonomy (phylum, class, order, family, and genus).

KEGG ortholog measurements (here called “ortholog” variables) were generated through whole-genome metagenomic shotgun sequencing (mWGS). Each ortholog was annotated with its KEGG orthology ID (“KO ID”).

### Genetics

Whole-genome sequencing (WGS) of part of the iPOP cohort (n = 73) was performed as part of a previously reported study^65^. Genomic DNA was isolated from whole blood, and WGS was performed using the Novogene PCR-free WGS platform, aiming for 90 gigabases (30x coverage) per sample. The read alignment and variant calling were performed using Sentieon’s DNAseq pipeline, which adheres to GATK best practices while providing significant computational efficiency and accuracy. The pipeline processes data from FASTQ to VCF and supports extensive QC measures, such as metrics for base quality score distribution, read alignment quality, and duplicate marking, which were integrated into the workflow. Sample- and variant-level QC for WGS was conducted using PLINK2 software, where samples with high missingness or excess heterozygosity and variants with high missingness or Hardy–Weinberg equilibrium (HWE) P values < 1 x 10^-6^ were filtered out. Genotype principal components (PCs) were calculated using PLINK2.

### Clinical tests

Blood samples were also analyzed for 51 well-established clinical tests, including a basic lipid panel, complete metabolic panel, complete blood count (CBC) with differential, high-sensitivity C-reactive protein (CRP), hemoglobin A1c (HbA1c), immunoglobulin M, estimated glomerular filtration rate (eGFR), and others. Across clinical tests, it was common for some values to be reported with a character string indicating that the lab value was above or below a particular value (usually the instrument’s limit of detection). In these cases, the lab values were imputed to be the threshold value (e.g., “<2” was recoded as 2) for use in subsequent analyses.

### PhenoAge

As a measure of biological aging, we implemented the PhenoAge model in iPOP using the formula provided by the authors^66,67^. PhenoAge uses a combination of chronological age and a variety of common clinical test metrics to estimate an individual’s biological age. PhenoAge was used as an alternative outcome of interest for the machine-learning prediction models.

### Data harmonization

The various clinical and molecular data sets were combined at the sample level, ensuring that different data types for a sample originated from the same time point. To organize the various omics data, analytes were given an identifier comprising the type of omics data and a unique number (e.g., “protein23”, “cytokine7”, “metabolite204”).

Study visits from the iPOP data repository from 2010 to 2017 were considered for subsequent analyses. Any study visits missing key covariate data (i.e., sex, age at visit, self-reported ethnicity, baseline BMI, or diabetes status) were excluded from analysis. Participants from a fiber diet study were also excluded. Furthermore, since our aim was to study patterns of molecular variation in the typical human multiome, we limited our analysis to routine study visits where participants self-reported as healthy (i.e., no reported illness, travel, recent medical care, acute stress, acute weight loss, etc.).

### Data transformations

The protein and lipid data were log_2_-transformed. The cytokine and metabolite data were examined for normality and were both found to be consistently right-skewed. A log_2_ transformation (with an offset of 1.1 added) was applied to each cytokine and metabolite variable.

Because the microbial taxa data (“microbe” variables) were proportions, the data were non-normally distributed. To facilitate analysis downstream, a log_2_ transformation (with an offset of 0.001 added) was applied to each variable.

### Dimensionality analysis

Each omic data set was analyzed separately with a principal component analysis (PCA). Each individual variable was first Z-score normalized before the PCA, which was performed using the prcomp function of the stats R package (v.4.4.3). The variance explained per PC was extracted and compared across omic data sets using scree (variance-per-PC) and avalanche (cumulative-variance-per-PC) plots.

Intraomic relationships between molecules of a given ome were assessed in several ways. First, all pairwise correlations between molecules were calculated for each omic data set, using all available samples for each ome and calculating the median value per participant across samples. Significant intraomic correlations were those with an FDR q value < 0.05. Heatmaps with hierarchical clustering (Euclidean distance) were generated for each ome (R package ComplexHeatmap, v.2.20.0).

A Gaussian graphical model (GGM) and network analysis were also performed for each individual ome along with a multiomic network combining all blood omes (i.e., proteins, lipids, cytokines, and metabolites)^68^. First, each molecular variable was Z-score transformed (i.e., “scaled”). A GGM was estimated using graphical lasso (glasso) and 100 values of lambda using the high-dimensional undirected graph estimation (huge) R package (v.1.3.5). The best model and optimal lambda were chosen according to the rotation information criterion (RIC). The resulting GGM was then visualized as a partial correlation graph network (R package ggraph v.2.2.1, igraph v.2.1.1, tidygraph v.1.3.1). Edge weights were defined as the partial correlation. Node centrality metrics (including degree, hub score, and authority score) were calculated for each molecule. Nodes were ranked according to the mean of their ranks for each individual centrality metric. Community membership was estimated with the cluster_louvain function^69^, and the modularity score for the graph was calculated. Networks were visualized with the graphopt algorithm.

### Modality analysis

For each protein, cytokine, lipid, and metabolite variable, we analyzed the modality of its distribution (i.e., whether each variable was unimodal, bimodal, trimodal, etc.). To facilitate the analysis, the median value across visits per participant per analyte was analyzed. Several different modality analysis methods were assessed, including the bimodal index^70^ (R package BimodalIndex, v.1.1.9), Hartigan’s dip statistic^71^ (R package diptest, v.0.77-1), Bayesian unimodal and multimodal estimation functions (R package LaplacesDemon v.16.1.6), and the bimodality coefficient^72^ (R package mousetrap, v.3.2.3). To identify a given variable as bimodal (or non-unimodal) for the Bimodal Index, a threshold of >1.3 was used to consider a variable bimodal (more conservative than the threshold of 1.1 suggested by the authors). For the bimodality coefficient, a threshold of > 0.555, as proposed by the method’s authors, was used to identify bimodal variables. The different modality metrics were compared by examining the distributions of the putatively bimodal analytes and the distribution of the metric values across the variables analyzed. The bimodal index (at a threshold of 1.3) appeared to best identify non-unimodal molecules and was the focus of our investigations moving forward.

The list of putatively non-unimodal analytes identified by the bimodal index was then further analyzed to understand potential drivers of the differences across the cohort. Non-unimodal analytes were tested for associations with sample storage duration, age, sex, baseline BMI, diabetic status, and any of the clinical tests. Analytes that had at least one significant (FDR q < 0.05) association were considered to be at least partly explained by known factors.

Unexplained analytes were then further examined for biological trends through enrichment analyses. Pathway enrichment was assessed using Gene Ontology (Biological Process) and KEGG (for proteins and cytokines), lipid class (lipids), and metabolite superpathway and subpathway (metabolites) annotations, using both standard enrichment and Gene Set Enrichment Analysis (GSEA) algorithms with the R package clusterProfiler (v.4.12.6). In every case, enrichment was calculated against the total set of molecules available in the iPOP study (e.g., protein enrichment was tested against the background of all proteins measured in iPOP). Significantly enriched annotations (P < 0.05 after FDR correction) were reported.

Unexplained bimodal analytes were further evaluated for potential genetic and dietary associations. We searched the GWAS catalog^73^ (accessed on 4/22/2026) to identify analytes with previously reported genome-wide significant quantitative trait loci (QTL) associations and extracted the corresponding variant information from the catalog. For each analyte, a representative variant was selected for the reported QTL association. We compared the analyte distributions across genotypes using linear regression. We then examined the distribution of residuals after fitting the linear regression and recalculated the BI to assess whether the QTL explained the observed bimodal pattern. To account for potential ancestry-related confounding, we also performed the same analyses with adjustment for the first five genotype PCs. For dietary associations, we tested the association of each bimodal molecule with dietary intake items from the surveys given to iPOP participants, controlling for age, sex, and BMI. Only 68 iPOP participants had sufficient dietary information to be included in this analysis.

### Personalized omic variation analysis

To compare the extent of population variation for each molecule, a coefficient of variation (CV) analysis was performed. The untransformed median value across visits per person per molecule was used to calculate the “interindividual CV” for each molecule, given the issues with interpreting the CV on log-transformed data, which can lead to inflated CVs. The distributions of these interindividual CVs were compared by omic type and molecular subtypes (for the lipids and metabolites). Differences in interindividual CV distributions were calculated with a Kruskal-Wallis test.

A second type of CV was also estimated, which we called the “intraindividual CV”. This intraindividual CV is an individual-and-molecule-level metric based on the CV of one participant’s levels of one molecule over all of their available data points, thus reflecting how much that molecule varies over time for that specific participant. The intraindividual CVs were only calculated when at least three measurements for a molecule were available for an individual. Pathway enrichment analyses (both hypergeometric and GSEA) were used to identify types of molecules with greater variation than others, using GO, KEGG, lipid class, and metabolite super- and subpathways as the annotations of interest.

The interindividual and intraindividual CVs were compared in several ways. First, at the molecular level, the interindividual CV for each molecule was compared to the mean intraindividual CV across participants for that molecule and visualized. The distributions of interindividual and intraindividual CVs for molecules were also compared within omic types and subclasses (namely, by lipid class, metabolite superpathway, and metabolite subpathway). Classes of molecules with a large difference between the mean interindividual CV and mean intraindividual CV were identified. Molecules with a mean intraindividual CV that was substantially different from the interindividual CV (defined as the ratio between the two) were identified and plotted. Molecules that differed in intraindividual CV from similar molecules were identified as those with a mean intraindividual CV that was more than 4 standard deviations greater than the mean intraindividual CV of that omic type, lipid class, metabolite superpathway, or metabolite subpathway.

Hypervariant individuals were also identified at the level of a single molecule, lipid class, metabolite superpathway, metabolite subpathway, and omic type. Given differences in sample count, follow-up time, and other factors in iPOP that might affect a single individual’s CV for a molecule, we considered this analysis more exploratory. A hypervariant individual was defined as someone whose intraindividual CV for a given molecule was more than 4 SDs greater than the population mean CV for that molecule. In other words, a hypervariant individual is someone whose variation for a molecule over time is much greater than that seen in the rest of the study population for that molecule. However, given the exploratory nature of this analysis, we considered this label more of a way to prioritize outlier samples than as a strict classification. In a similar fashion, hypervariant individuals were those whose mean intraindividual CV for a subset of molecules (i.e., by omic type, lipid class, metabolite superpathway, or metabolite subpathway) was >= 4 SDs greater than that of the rest of the population. The distributions of these hypervariant individuals’ intraindividual CVs compared to the rest of the population were plotted.

### Molecular associations with demographics

The association of each molecule with demographic and anthropometric variables (age, sex, and BMI) was estimated in a series of regression analyses using all available sample-level data. For each molecule, a linear mixed model was run with the level of the molecule regressed on visit age, sex, baseline BMI, population (White, Asian, Black, and Hispanic), and diabetic status (diabetic, prediabetic, or normoglycemic), along with a random intercept for the participant. The marginal and conditional R^2^ were estimated for each model, and the effect estimates, P values, and partial R^2^ values were estimated for each covariate of interest. Statistically significant molecular associations were identified as those with an FDR q value < 0.05 to account for multiple testing. Regressions were run using the lme4 (v.4.4), qvalue (v.2.36.0), broom.mixed (v.0.2.9.5), MuMIn (v.1.48.4), and partR2 (v.0.9.2) packages.

Key biological pathways enriched among the associated molecules were identified through a series of enrichment and GSEA analyses, as described above. Enrichment analyses were run against GO BP, KEGG, lipid subclass, metabolite superpathway, and metabolite subpathway annotations.

### Omic prediction models for demographics and clinical tests

Using an array of machine learning models, we sought to determine the omes, analytes, and model types that best predicted the four demographic features (age, PhenoAge^66^, sex, BMI) and 49 routine clinical laboratory tests (the insulin and fasting insulin measures were excluded due to missingness being > 50%). For each outcome, we tested different sets of predictors. Generally, each predictor set was the set of all variables for one ome (e.g., all metabolite variables). We also tested combined predictor sets using either all blood omic variables or all microbial variables together. Thus, for i number of omes with j number of omic features, the models were fitted using the following equations:

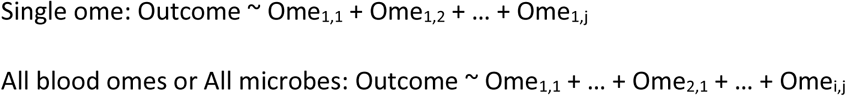

To maximize the number of available observations for model training, we created three data sets. A complete blood omics data set (i.e., samples with lipidomics, proteomics, metabolomics, and cytokines) was generated by restricting to visits where all blood omics data were collected (n = 89 subjects, 452 visits; titled the “all blood omes” data set). A complete microbiome data set (i.e., samples with all nasal/stool microbe and ortholog data) was similarly created (n = 79 subjects, 323 visits; titled the “all microbes” data set). A third, smaller “complete” data set with all omics data was generated (i.e., samples with all blood omic and all microbiome data available; n = 75 subjects, 295 visits) and used in secondary analyses to compare the blood ome vs. microbe model performances when trained on data sets of the same size. Prior to modeling, predictors with zero variance were removed, and the remaining features were transformed using the Z score. Model performance was determined by omics data alone without adding any demographic features (e.g., BMI, age) that may have explained a large portion of variance in the outcomes.

Models were implemented using the tidymodels package in R^74^. We used five model types, including regularized linear regression (ridge regression, Least Absolute Shrinkage and Selection Operator [LASSO], and elastic net using the glmnet package^75^) and tree-based models (random forest and eXtreme Gradient Boosting [XGBoost] with the ranger^76^ and xgboost^77^ packages). Each data set was divided into a 75% train and 25% held-out test set. To prevent data leakage, train-test splits were constructed such that all visits from a unique subject were placed within the same set (*group_initial_split*; tidymodels). Stratified sampling was also performed based on the number of visits (< 3, 3-6, and > 6 visits) each participant had, to ensure train-test splits had similar proportions of visit frequencies. These procedures were also applied to all cross-validation folds (*group_vfold_cv*; tidymodels).

Hyperparameters were tuned using 5-fold cross-validation in the training data set. The best-performing hyperparameters were selected, and a final model was fitted on the training set. Observations were weighted by 1/n—where *n* is the number of visits per subject—during model fitting and prediction to reduce the influence of participants who were overrepresented in the data set. Model performance (R^2^ for continuous outcomes and ROC AUC for binary outcomes) was evaluated on the 25% held-out test set. For each outcome (n = 52), predictor set (n = 10), and model type (n = 5) combination, we reported the average test set performance (e.g., mean R^2^ [SD]) across 10 random seeds.

For feature interpretation, we refit a final model for each outcome using the full blood omic feature sets (“all blood omes”) or microbial feature set (“all microbes”). Feature rankings were considered exploratory and were not used to estimate predictive performance, which was instead based only on the aforementioned held-out test sets. For each outcome, we identified the model type with the highest mean R^2^ from the “all blood omes” or “all microbes” models. Using this model type, we refitted a final model on the full data set (n = 452 or n = 323), omitting the train/test split, for that outcome. The coefficients for the linear models were extracted, and the feature importance scores for the random forest and gradient-boosted models were generated using the VIP package^78^.

### Replication analyses

Replication of the cross-omic PCA findings was performed in a sister multiomics cohort called the Human Personalized Omics Profiling (hPOP) study. This cohort has been recently described in detail^17^. In short, it is a primarily cross-sectional, multiomic cohort comprising 322 participants from around the world. Relevant to our purposes here, hPOP includes plasma proteomic (Olink and Selected Reaction Monitoring (SRM), n = 200 proteins), plasma metabolomic (targeted [Biocrates], n = 435 metabolites; untargeted [LC-MS/MS], n = 401 metabolites), urine metabolomic (untargeted [LC-MS/MS]), and plasma lipidomic (Lipidyzer, n = 711 lipids) data.

Replication of the PCA analysis was performed in the hPOP samples, with interomic comparisons made as described above with an avalanche plot.

Replication of the modality analysis was performed in hPOP. For each analyte that was bimodal in iPOP and also measured in hPOP, we calculated the bimodal index in hPOP using the BimodalIndex package. Replication of a bimodal signal was assessed using the bimodal index and a threshold of >= 1.3 (same as that used in the iPOP analysis).

Replication of the demographic and clinical test prediction modeling was also performed in hPOP. Using the same machine-learning training and testing approach described above, we compared the predictive performance of different omes for different demographic and clinical outcomes. The predictor variable sets analyzed in hPOP included the targeted plasma metabolomic data, the plasma proteomic data (Olink and SRM), lipids, and hormones (n = 9 hormones), The demographic outcomes included age, sex, and BMI. The clinical tests were much fewer than in iPOP, including only 5 different tests (fasting glucose, fasting creatinine, high-sensitivity C-reactive protein, albumin, and total alkaline phosphatase). Replication was focused on the relative performance of different omes for different outcomes as well as the overall performance of the best models for each outcome. Given the general similarity in performance across different model architectures from the main analysis in iPOP, the modeling process for replication in hPOP was simplified to use only LASSO models.

## Acknowledgments

We would like to thank the participants, research coordinators, and staff of the iPOP cohort, without whose support this research would not have been possible. We also thank Dr. John Tamaresis and Jeandele Elliot from the Stanford Department of Biomedical Data Science Data Studio for their advice. We thank Lori Buesking for her advice on visualization. The research reported in this publication was supported by the National Center for Advancing Translational Sciences of the National Institutes of Health under Award Number UM1TR004921. This work was supported by National Institutes of Health (NIH) grants U54DK102556 (M.P.S.), R01 DK110186-03 (M.P.S.), R01HG008164 (M.P.S.), NIH S10OD020141 (M.P.S.), UL1 TR001085 (M.P.S.) and P30DK116074 (M.P.S.) and by the Stanford Data Science Initiative (M.P.S.). The funders had no role in study design, data collection and analysis, decision to publish or preparation of the manuscript. The content is solely the responsibility of the authors and does not necessarily represent the official views of the National Institutes of Health.

## Author contributions

Author DJP conceptualized the study. Authors DJP, MAA, AKR, HI, TM, and XS contributed to the design of the analyses. Authors DJP, MAA, AKR, and HI conducted the analyses in the iPOP cohort. Author NB conducted the replication analyses in the hPOP cohort. Authors TM, SMSR, KC, DH, SA, and MPS contributed to the collection and preparation of the iPOP data. Authors DJP, MAA, AKR, and HI drafted the manuscript with input from all other authors. All authors contributed to the interpretation of the findings.

## Data availability

The data for the iPOP cohort can be found on the Human Microbiome 2 project site (https://portal.hmpdacc.org/) and the Stanford iPOP site (http://med.stanford.edu/ipop.html). The data for the hPOP cohort can be found at the Stanford hPOP site (https://hpopdb.stanford.edu/).

## List of Supplementary Tables

Supplementary Table 1. Sample summary.

Supplementary Table 2. Annotation information for omic variables.

Supplementary Table 3. Clinical test descriptions.

Supplementary Table 4. Variance per PC per ome.

Supplementary Table 5. All FDR-significant interomic correlations.

Supplementary Table 6. Variable-level network metrics.

Supplementary Table 7. Count of skewed variables by ome.

Supplementary Table 8. Bimodality metrics for bimodal variables not associated with demographic, clinical, or other factors.

Supplementary Table 9. Genotype effects on molecule levels without controlling for principal components.

Supplementary Table 10. Genotype effects on molecule levels controlling for 5 principal components.

Supplementary Table 11. Interindividual CV by lipid class.

Supplementary Table 12. Interindividual CV by metabolite superpathway.

Supplementary Table 13. Interindividual CV by metabolite subpathway.

Supplementary Table 14. Significantly enriched functional categories among intraindividual CVs from GSEA.

Supplementary Table 15. Intraindividual CV by lipid class.

Supplementary Table 16. Intraindividual CV by metabolite superpathway.

Supplementary Table 17. Interindividual CV by metabolite subpathway.

Supplementary Table 18. Longitudinal models of molecules regressed on demographics.

Supplementary Table 19. Standard enrichment analysis results for molecules significantly associated with demographics.

Supplementary Table 20. GSEA enriched annotations among molecules associated with demographics.

Supplementary Table 21. Summary of the all blood omes data set.

Supplementary Table 22. Summary of the all microbes data set.

Supplementary Table 23. Average performance of all demographic prediction models across 10 seeds.

Supplementary Table 24. Top features and hyperparameters for the best demographic prediction models.

Supplementary Table 25. Hyperparameters for the best demographic prediction models.

Supplementary Table 26. Best performing models for each demographic in the replication data set, hPOP.

Supplementary Table 27. Performance of all machine-learning models for each clinical test.

Supplementary Table 28. Best performing models for each clinical test.

Supplementary Table 29. Coefficients and feature importance of blood omic features for predicting clinical tests.

Supplementary Table 30. Coefficients and feature importance of microbial features for predicting clinical tests.

Supplementary Table 31. Best performing models for each clinical test in the replication data set, hPOP.

## List of Supplementary Figures

Supplementary Figure 1. Visit count distribution.

Supplementary Figure 2. Time between visits distribution.

Supplementary Figure 3. Visit span distribution.

Supplementary Figure 4. Age distribution.

Supplementary Figure 5. BMI distribution.

Supplementary Figure 6. Visit count by ome.

Supplementary Figure 7. Variable count by ome.

Supplementary Figure 8. PCA scree plot in iPOP.

Supplementary Figure 9. PCA avalanche plot in the replication cohort, hPOP.

Supplementary Figure 10. Correlation heat map for the stool microbiome.

Supplementary Figure 11. Correlation heat map for stool orthologs.

Supplementary Figure 12. Correlation heat map for the nasal microbiome.

Supplementary Figure 13. Correlation heat map for nasal orthologs.

Supplementary Figure 14. Variable skew after transformation.

Supplementary Figure 15. Distribution of SERPINA5 values before and after controlling for its pQTL.

Supplementary Figure 16. Distribution of interindividual CVs by ome.

Supplementary Figure 17. Distribution of intraindividual CVs by ome.

Supplementary Figure 18. Comparison of interindividual and intraindividual CVs.

Supplementary Figure 19. Longitudinal trajectory of IFNB.

Supplementary Figure 20. Longitudinal trajectory of pregnanediol-3-glucuronide.

Supplementary Figure 21. Longitudinal trajectory of IGHD.

Supplementary Figure 22. Overview of the machine-learning models for predicting demographics and clinical tests.

Supplementary Figure 23. Sensitivity analysis of machine-learning models predicting sex.

Supplementary Figure 24. Sensitivity analysis of machine-learning models predicting BMI.

Supplementary Figure 25. Sensitivity analysis of machine-learning models predicting age and PhenoAge.

Supplementary Figure 26. Sensitivity analysis of machine-learning models predicting age and PhenoAge with overrepresented participant excluded.

Supplementary Figure 27. Top predictors of clinical test models classified as platform validation.

Supplementary Figure 28. Top predictors of clinical test models classified as having potential clinical utility.

Supplementary Figure 29. Comparison of blood omes and microbial features in predicting clinical tests when using data sets of the same size.

Supplementary Figure 30. Comparison of machine-learning model types for predicting clinical tests.

## References

1. Collins, F. S. & Varmus, H. A New Initiative on Precision Medicine. New England Journal of Medicine 372, 793–795 (2015).

2. National Research Council (US) Committee on A Framework for Developing a New Taxonomy of Disease. Toward Precision Medicine: Building a Knowledge Network for Biomedical Research and a New Taxonomy of Disease. (National Academies Press (US), Washington (DC), 2011).

3. Ashley, E. A. Towards precision medicine. Nat Rev Genet 17, 507–522 (2016).

4. Bycroft, C. et al. The UK Biobank resource with deep phenotyping and genomic data. Nature 562, 203–209 (2018).

5. Ferkingstad, E. et al. Large-scale integration of the plasma proteome with genetics and disease. Nat Genet 53, 1712–1721 (2021).

6. Kurki, M. I. et al. FinnGen provides genetic insights from a well-phenotyped isolated population. Nature 613, 508–518 (2023).

7. Nagai, A. et al. Overview of the BioBank Japan Project: Study design and profile. J Epidemiol 27, S2–S8 (2017).

8. Wong, E. et al. The Singapore National Precision Medicine Strategy. Nat Genet 55, 178–186 (2023).

9. Gaziano, J. M. et al. Million Veteran Program: A mega-biobank to study genetic influences on health and disease. J Clin Epidemiol 70, 214–223 (2016).

10. Taliun, D. et al. Sequencing of 53,831 diverse genomes from the NHLBI TOPMed Program. Nature 590, 290–299 (2021).

11. Sun, B. B. et al. Plasma proteomic associations with genetics and health in the UK Biobank. Nature 622, 329–338 (2023).

12. Panyard, D. J., Yu, B. & Snyder, M. P. The metabolomics of human aging: Advances, challenges, and opportunities. Sci Adv 8, eadd6155 (2022).

13. Zhang, S. et al. A metabolomic profile of biological aging in 250,341 individuals from the UK Biobank. Nat Commun 15, 8081 (2024).

14. Beyene, H. B. et al. High-coverage plasma lipidomics reveals novel sex-specific lipidomic fingerprints of age and BMI: Evidence from two large population cohort studies. PLOS Biology 18, e3000870 (2020).

15. Mäkinen, V.-P. & Ala-Korpela, M. Influence of age and sex on longitudinal metabolic profiles and body weight trajectories in the UK Biobank. Int J Epidemiol 53, dyae055 (2024).

16. Fryar, C. D., Carroll, M. D. & Afful, J. Prevalence of overweight, obesity, and severe obesity among adults aged 20 and over: United States, 1960–1962 through 2017–2018. NCHS Health E-Stats (2020).

17. Barapour, N. et al. A comparison of deep multiomics profiles across ethnicity, geography, and age. Cell 189, 3004–3024.e35 (2026).

18. Cheng, J. et al. Identification of 2-Piperidone as a Biomarker of CYP2E1 Activity Through Metabolomic Phenotyping. Toxicol Sci 135, 37–47 (2013).

19. Shen, X. et al. Sex-specific associations between pregnancy zone protein and acute coronary syndrome: a case-control study. Biol Sex Differ 17, 135 (2026).

20. Casabiell, X. et al. Leptin, reproduction and sex steroids. Pituitary 4, 93–99 (2001).

21. Nakhla, A. M. et al. 5 alpha-Androstan-3 alpha,17 beta-diol is a hormone: stimulation of cAMP accumulation in human and dog prostate. J Clin Endocrinol Metab 80, 2259–2262 (1995).

22. Choi, R. H., Tatum, S. M., Symons, J. D., Summers, S. A. & Holland, W. L. Ceramides and other sphingolipids as drivers of cardiovascular disease. Nat Rev Cardiol 18, 701–711 (2021).

23. Sniderman, A. D. et al. Apolipoprotein B Particles and Cardiovascular Disease: A Narrative Review. JAMA Cardiol 4, 1287–1295 (2019).

24. Sehayek, D. et al. ApoB, LDL-C, and non-HDL-C as markers of cardiovascular risk. Journal of Clinical Lipidology 19, 844–859 (2025).

25. Gallego-Paüls, M. et al. Variability of multi-omics profiles in a population-based child cohort. BMC Med 19, 166 (2021).

26. Zhong, W. et al. Whole-genome sequence association analysis of blood proteins in a longitudinal wellness cohort. Genome Med 12, 53 (2020).

27. Romanov, N. et al. Disentangling Genetic and Environmental Effects on the Proteotypes of Individuals. Cell 177, 1308–1318.e10 (2019).

28. Garcia-Bernardo, J. & Dunlop, M. J. Phenotypic Diversity Using Bimodal and Unimodal Expression of Stress Response Proteins. Biophysical Journal 110, 2278–2287 (2016).

29. Dobrzyński, M., Fey, D., Nguyen, L. K. & Kholodenko, B. N. Bimodal Protein Distributions in Heterogeneous Oscillating Systems. in Computational Methods in Systems Biology (eds Gilbert, D. & Heiner, M.) 17–28 (Springer, Berlin, Heidelberg, 2012). doi:10.1007/978-3-642-33636-2_3.

30. Stearns-Kurosawa, D. J., Burgin, C., Parker, D., Comp, P. & Kurosawa, S. Bimodal distribution of soluble endothelial protein C receptor levels in healthy populations. Journal of Thrombosis and Haemostasis 1, 855–856 (2003).

31. Haehner, B. D. et al. Bimodal distribution of renal cytochrome P450 3A activity in humans. Molecular Pharmacology 50, 52–59 (1996).

32. Ling, M.-Y., Chiu, L.-J., Hsieh, C.-C. & Shu, C.-C. Dimerization induces bimodality in protein number distributions. Biosystems 223, 104812 (2023).

33. Bechtel, Y. C., Bonaiti-Pellie, C., Poisson, N., Magnette, J. & Bechtel, P. R. A population and family study N-acetyltransferase using caffeine urinary metabolites. Clinical Pharmacology & Therapeutics 54, 134–141 (1993).

34. Westenberg, H. G. M. & Verhoeven, W. M. A. CSF monoamine metabolites in patients and controls: support for a bimodal distribution in major affective disorders. Acta Psychiatrica Scandinavica 78, 541–549 (1988).

35. Bentsen, H. et al. Bimodal Distribution of Polyunsaturated Fatty Acids in Schizophrenia Suggests Two Endophenotypes of the Disorder. Biological Psychiatry 70, 97–105 (2011).

36. Mocking, R. J. T. et al. Bimodal Distribution of Fatty Acids in Recurrent Major Depressive Disorder. Biological Psychiatry 71, e3–e5 (2012).

37. Boyd, E. M. & Connell, W. F. FURTHER STUDIES OF PLASMA LIPIDS IN TOXIC GOITRE: EVIDENCE SUGGESTING A BIMODAL DISTRIBUTION. QJM 8, 41–45 (1939).

38. Metwally, A. A. et al. Insulin resistance prediction from wearables and routine blood biomarkers. Nature 1–11 (2026) doi:10.1038/s41586-026-10179-2.

39. Schüssler-Fiorenza Rose, S. M., et al. A longitudinal big data approach for precision health. Nat Med 25, 792–804 (2019).

40. Tippetts, T. S., Holland, W. L. & Summers, S. A. Cholesterol—The Devil You Know; Ceramide—The Devil You Don’t. Trends Pharmacol Sci 42, 1082–1095 (2021).

41. Laaksonen, R. et al. Plasma ceramides predict cardiovascular death in patients with stable coronary artery disease and acute coronary syndromes beyond LDL-cholesterol. Eur Heart J 37, 1967–1976 (2016).

42. Panyard, D. J. et al. Large-scale proteome and metabolome analysis of CSF implicates altered glucose and carbon metabolism and succinylcarnitine in Alzheimer’s disease. Alzheimers Dement 19, 5447–5470 (2023).

43. Gordis, L. Epidemiology. (Elsevier Saunders, Philadelphia, PA, 2014).

44. Moaddel, R. et al. Proteomics in aging research: A roadmap to clinical, translational research. Aging Cell 20, e13325 (2021).

45. Sen, P., Shah, P. P., Nativio, R. & Berger, S. L. Epigenetic Mechanisms of Longevity and Aging. Cell 166, 822–839 (2016).

46. Levine, M. E. et al. An epigenetic biomarker of aging for lifespan and healthspan. Aging (Albany NY) 10, 573–591 (2018).

47. Argentieri, M. A. et al. Proteomic aging clock predicts mortality and risk of common age-related diseases in diverse populations. Nat Med 30, 2450–2460 (2024).

48. Avsec, Ž., et al. Advancing regulatory variant effect prediction with AlphaGenome. Nature 649, 1206–1218 (2026).

49. Reel, P. S., Reel, S., Pearson, E., Trucco, E. & Jefferson, E. Using machine learning approaches for multi-omics data analysis: A review. Biotechnology Advances 49, 107739 (2021).

50. Hu, J. & Szymczak, S. A review on longitudinal data analysis with random forest. Brief Bioinform 24, bbad002 (2023).

51. Catling, F. J. R. & Wolff, A. H. Temporal convolutional networks allow early prediction of events in critical care. J Am Med Inform Assoc 27, 355–365 (2020).

52. Centers for Disease Control and Prevention. National Diabetes Statistics Report. https://gis.cdc.gov/grasp/diabetes/diabetesatlas-statsreport.html (2026).

53. Katz, D. H. et al. Proteomic profiling platforms head to head: Leveraging genetics and clinical traits to compare aptamer- and antibody-based methods. Science Advances 8, eabm5164 (2022).

54. Raffield, L. M. et al. Comparison of Proteomic Assessment Methods in Multiple Cohort Studies. Proteomics 20, e1900278 (2020).

55. Kirsher, D. Y. et al. Current landscape of plasma proteomics from technical innovations to biological insights and biomarker discovery. Commun Chem 8, 279 (2025).

56. Bergström, S. et al. Longitudinal protein profiling of blood during childhood into early adulthood. Nat Commun 17, 3700 (2026).

57. Zhou, W. et al. Longitudinal multi-omics of host–microbe dynamics in prediabetes. Nature 569, 663–671 (2019).

58. Ahadi, S. et al. Personal aging markers and ageotypes revealed by deep longitudinal profiling. Nat Med 26, 83–90 (2020).

59. Hornburg, D. et al. Dynamic lipidome alterations associated with human health, disease and ageing. Nat Metab 5, 1578–1594 (2023).

60. Shen, X. et al. Nonlinear dynamics of multi-omics profiles during human aging. Nat Aging 4, 1619–1634 (2024).

61. Piening, B. D. et al. Integrative Personal Omics Profiles during Periods of Weight Gain and Loss. Cell Systems 6, 157–170.e8 (2018).

62. Contrepois, K., Jiang, L. & Snyder, M. Optimized Analytical Procedures for the Untargeted Metabolomic Profiling of Human Urine and Plasma by Combining Hydrophilic Interaction (HILIC) and Reverse-Phase Liquid Chromatography (RPLC)–Mass Spectrometry. Mol Cell Proteomics 14, 1684–1695 (2015).

63. Johnson, W. E., Li, C. & Rabinovic, A. Adjusting batch effects in microarray expression data using empirical Bayes methods. Biostatistics 8, 118–127 (2007).

64. Rosenberg-Hasson, Y. HIMC Protocol: Luminex Multiplex Analysis.

65. Zhang, S. et al. Dissecting the genetic complexity of myalgic encephalomyelitis/chronic fatigue syndrome via deep learning-powered genome analysis. 2025.04.15.25325899 Preprint at 10.1101/2025.04.15.25325899 (2025).

66. Liu, Z. et al. A new aging measure captures morbidity and mortality risk across diverse subpopulations from NHANES IV: A cohort study. PLOS Medicine 15, e1002718 (2018).

67. Liu, Z., et al. Correction: A new aging measure captures morbidity and mortality risk across diverse subpopulations from NHANES IV: A cohort study. PLOS Medicine 16, e1002760 (2019).

68. Shutta, K. H., De Vito, R., Scholtens, D. M. & Balasubramanian, R. Gaussian graphical models with applications to omics analyses. Stat Med 41, 5150–5187 (2022).

69. Blondel, V. D., Guillaume, J.-L., Lambiotte, R. & Lefebvre, E. Fast unfolding of communities in large networks. J. Stat. Mech. 2008, P10008 (2008).

70. Wang, J., Wen, S., Symmans, W. F., Pusztai, L. & Coombes, K. R. The Bimodality Index: A Criterion for Discovering and Ranking Bimodal Signatures from Cancer Gene Expression Profiling Data. Cancer Inform 7, 199–216 (2009).

71. Hartigan, J. A. & Hartigan, P. M. The Dip Test of Unimodality. The Annals of Statistics 13, 70–84 (1985).

72. Pfister, R., Schwarz, K. A., Janczyk, M., Dale, R. & Freeman, J. Good things peak in pairs: a note on the bimodality coefficient. Front. Psychol. 4, (2013).

73. Cerezo, M. et al. The NHGRI-EBI GWAS Catalog: standards for reusability, sustainability and diversity. Nucleic Acids Res 53, D998–D1005 (2025).

74. Kuhn, M. & Wickham, H. Tidymodels: a collection of packages for modeling and machine learning using tidyverse principles. (2020).

75. Friedman, J., Hastie, T. & Tibshirani, R. Regularization Paths for Generalized Linear Models via Coordinate Descent. J Stat Softw 33, 1–22 (2010).

76. Wright, M. N. & Ziegler, A. ranger: A Fast Implementation of Random Forests for High Dimensional Data in C++ and R. Journal of Statistical Software 77, 1–17 (2017).

77. Chen, T., et al. xgboost: Extreme Gradient Boosting. (2026).

78. Greenwell, B., M. & Boehmke, B., C. Variable Importance Plots—An Introduction to the vip Package. The R Journal 12, 343 (2020).

