## Supplementary Note for "Decoding personal and population variability in the human multiome"

*Omic bimodality*

For proteins[1–5], metabolites[6,7], and lipids[8,9], bimodal distributions have been observed with potentially important biological or medical implications, even as early as 1939[10]. For instance, various fatty acids and monoamines have been observed to be bimodal in certain psychiatric conditions, potentially indicating different disease endophenotypes that would be of utmost relevance to precision medicine[7–9]. In other cases, bimodal protein distributions were connected to different environmental or cellular conditions[3,5].

*Predictors of aging*

In the case of ageing, our findings highlight metabolites of branched chain amino acid (BCAA) synthesis and RNA metabolism in addition to the cytokine VCAM1. Our results imply circulating BCAAs (Valine, Isoleucine, and leucine) and their downstream metabolites (2-Hydroxyisocaproic acid) decrease with aging. The role of BCAA metabolism in ageing and ageing-related diseases is still not clearly understood with the literature presenting seemingly contradictory findings. On the one hand, BCAA supplementation leading to elevated BCAA and metabolite levels has been associated with insulin resistance, impaired memory, and neurodegeneration in both mice and humans. However, another growing body of evidence is beginning to demonstrate BCAA supplementation in combination with exercise could stimulate mitochondrial function and protein synthesis. Larger studies with more statistical power must be carried out to validate these results.

Another metabolite, 5-methyluridine is most known for its role as a protective RNA modification that preserves the stability of tRNAs and other ncRNAs. However, a new study has just been released that identified associations of increased levels of this modified riboside in MS. This is currently under investigation but our findings here imply that the role of 5-methyluridine might be applied to aging more generally as opposed to specific diseases.

Most interestingly, VCAM1, a cytokine, was the most influential negative predictor of age in our models. However, a recent study by Yousef and colleagues (Yousef et al. 2019) reported an opposite direction of effect with age in mice, and VCAM1 was associated with cerebrovascular inflammation. There could be a number of explanations for these contradictory findings including the way our cohort was assembled. As previously mentioned, the individuals in our cohort come from relatively affluent areas with healthy diets, potentially experiencing less inflammatory insults and thus introducing a survival bias. That is, we could have “superagers” in our cohort that do not express VCAM1 at similar levels to the general population. This result should be further investigated in separate cohorts.

**References**


1. Dobrzyński, M., Fey, D., Nguyen, L. K. & Kholodenko, B. N. Bimodal Protein Distributions in Heterogeneous Oscillating Systems. in *Computational Methods in Systems Biology* (eds Gilbert, D. & Heiner, M.) 17–28 (Springer, Berlin, Heidelberg, 2012). doi:10.1007/978-3-642-33636-2_3.
2. Stearns-Kurosawa, D. J., Burgin, C., Parker, D., Comp, P. & Kurosawa, S. Bimodal distribution of soluble endothelial protein C receptor levels in healthy populations. *Journal of Thrombosis and Haemostasis* **1**, 855–856 (2003).
3. Garcia-Bernardo, J. & Dunlop, M. J. Phenotypic Diversity Using Bimodal and Unimodal Expression of Stress Response Proteins. *Biophysical Journal* **110**, 2278–2287 (2016).
4. Haehner, B. D. *et al.* Bimodal distribution of renal cytochrome P450 3A activity in humans. *Molecular Pharmacology* **50**, 52–59 (1996).
5. Ling, M.-Y., Chiu, L.-J., Hsieh, C.-C. & Shu, C.-C. Dimerization induces bimodality in protein number distributions. *Biosystems* **223**, 104812 (2023).
6. Bechtel, Y. C., Bonaiti-Pellie, C., Poisson, N., Magnette, J. & Bechtel, P. R. A population and family study N-acetyltransferase using caffeine urinary metabolites. *Clinical Pharmacology & Therapeutics* **54**, 134–141 (1993).
7. Westenberg, H. G. M. & Verhoeven, W. M. A. CSF monoamine metabolites in patients and controls: support for a bimodal distribution in major affective disorders. *Acta Psychiatrica Scandinavica* **78**, 541–549 (1988).
8. Bentsen, H. *et al.* Bimodal Distribution of Polyunsaturated Fatty Acids in Schizophrenia Suggests Two Endophenotypes of the Disorder. *Biological Psychiatry* **70**, 97–105 (2011).
9. Mocking, R. J. T. *et al.* Bimodal Distribution of Fatty Acids in Recurrent Major Depressive Disorder. *Biological Psychiatry* **71**, e3–e5 (2012).

10. Boyd, E. M. & Connell, W. F. FURTHER STUDIES OF PLASMA LIPIDS IN TOXIC GOITRE: EVIDENCE SUGGESTING A BIMODAL DISTRIBUTION. *QJM* **8**, 41–45 (1939).
