## Supplementary Figures for "Decoding personal and population variability in the human multiome"

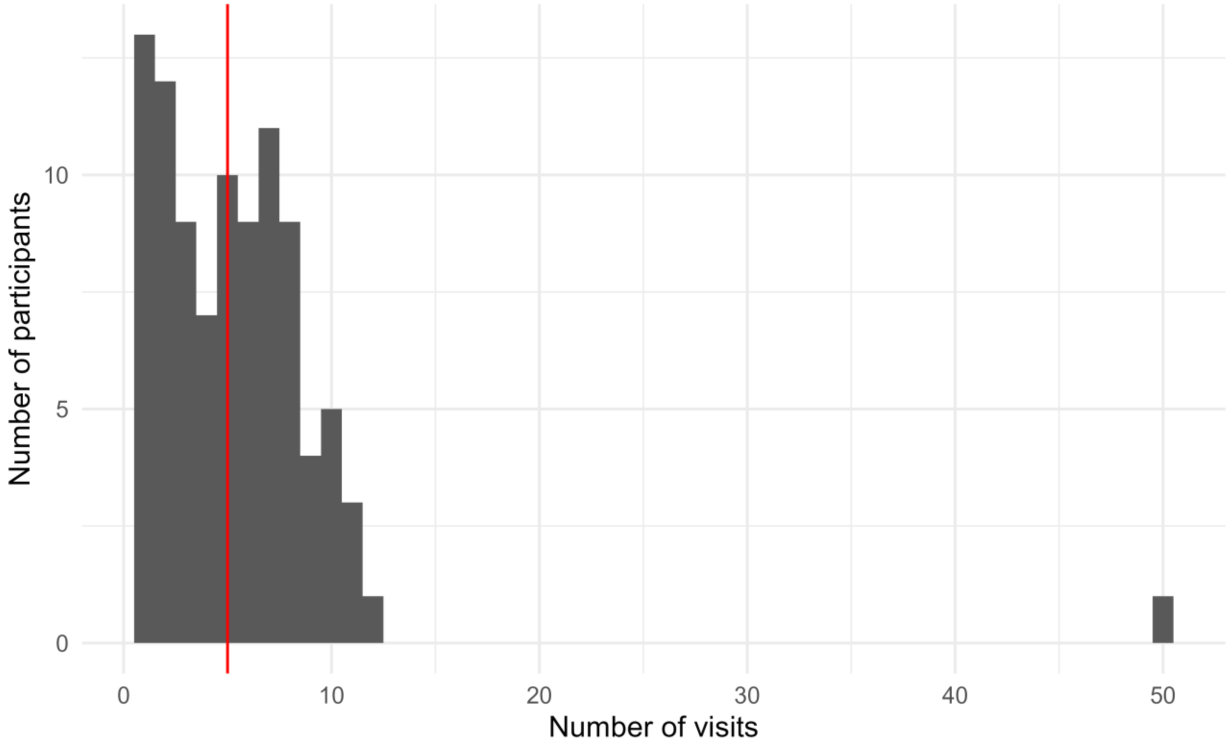

**Supplementary Figure 1. Visit count distribution.** The number of visits per participant is shown for the iPOP study cohort. The median number of visits is indicated by the red line.

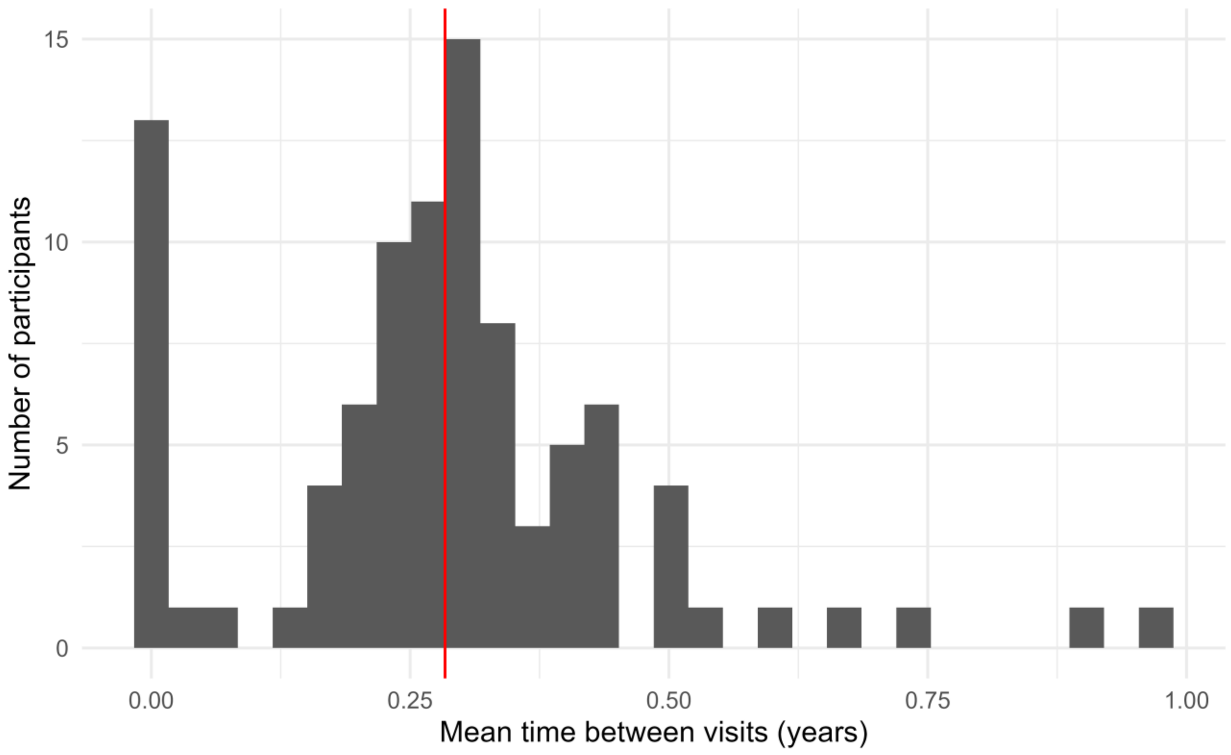

**Supplementary Figure 2. Time between visits distribution.** The distribution of the mean time between visits per participant is shown for the iPOP study cohort. Participants with only a single visit are shown with a mean time between visits of 0. The mean of these values across the study cohort is shown in red.

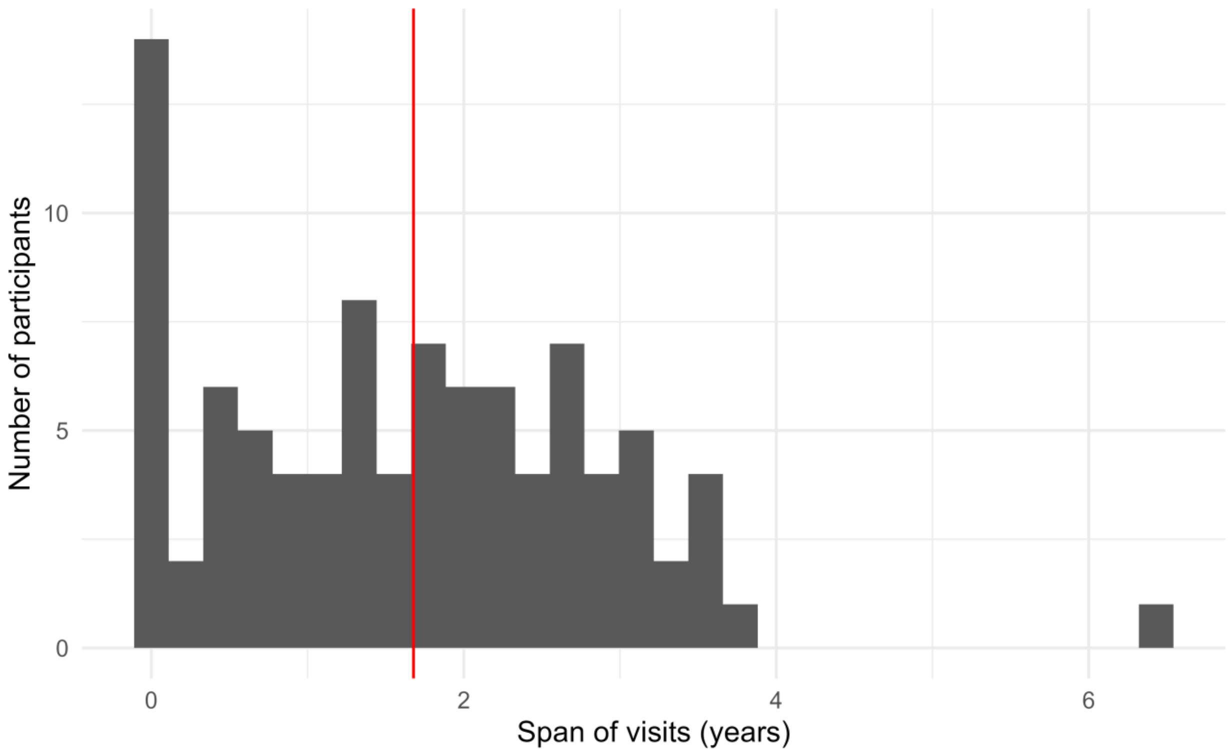

**Supplementary Figure 3. Visit span distribution.** The distribution of the total length of time that a participant was in the iPOP study is shown. The red line indicates the median span of time in the study.

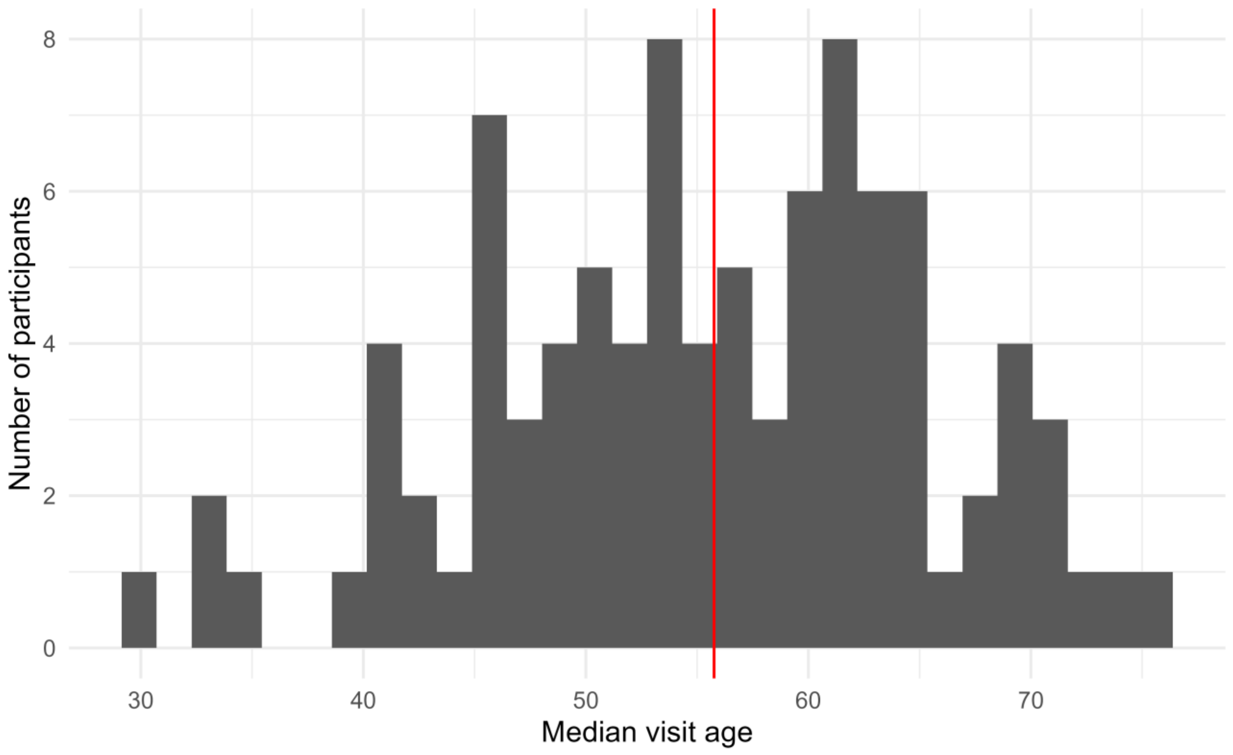

**Supplementary Figure 4. Age distribution.** The distribution of median visit age is shown for the iPOP study cohort. The red line indicates the median of these values.

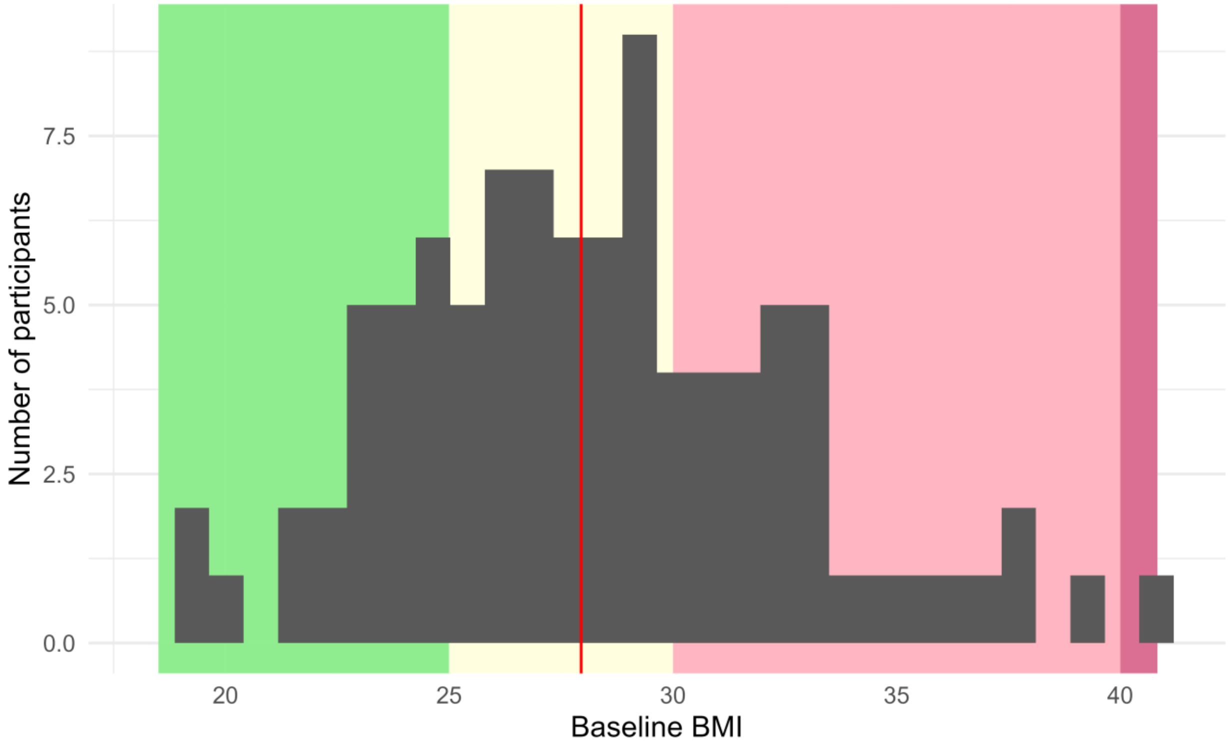

**Supplementary Figure 5. BMI distribution.** The distribution of baseline BMI is shown for the iPOP study cohort. The red line indicates the median BMI. The Centers of Disease Control adult BMI category thresholds are shown in color underneath the histogram: healthy weight (green), overweight (yellow), obesity (Class 1 or Class 2, pink), and severe obesity (Class 3, red).

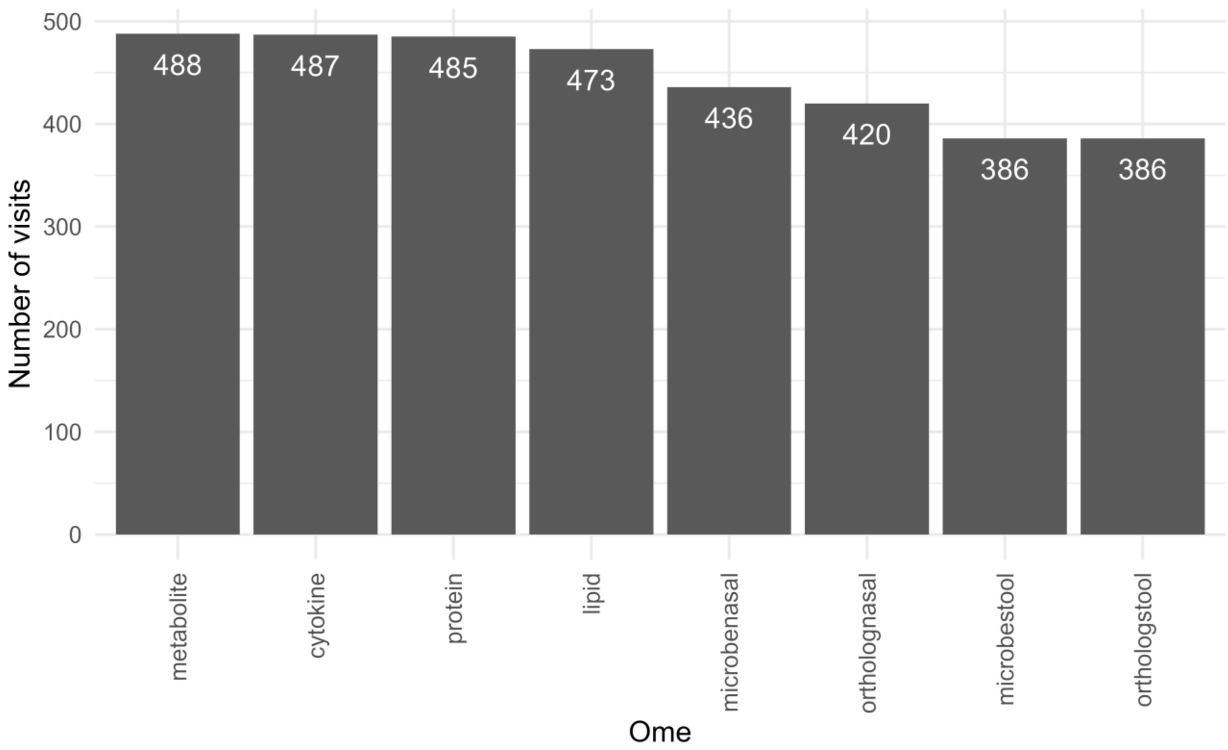

**Supplementary Figure 6. Visit count by ome.** The number of visits available per ome in the iPOP study cohort is shown.

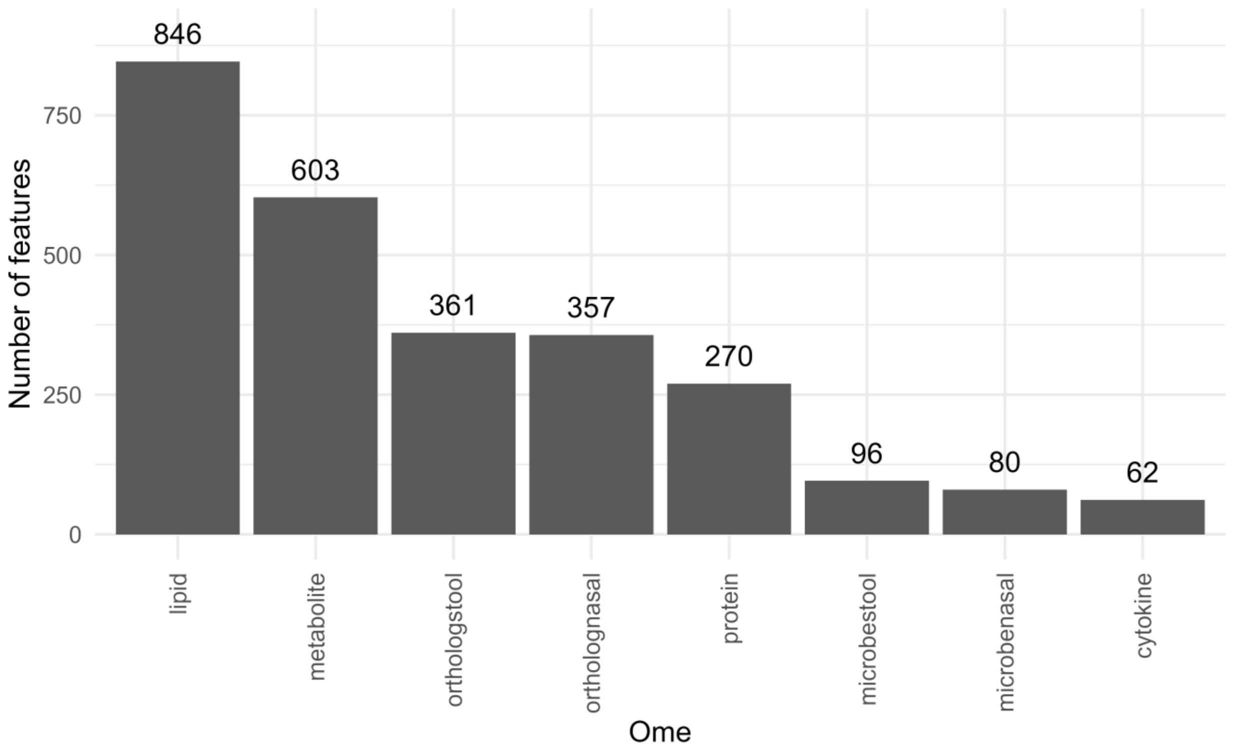

**Supplementary Figure 7. Variable count by ome.** The number of variables available for each ome from the iPOP study cohort is shown.

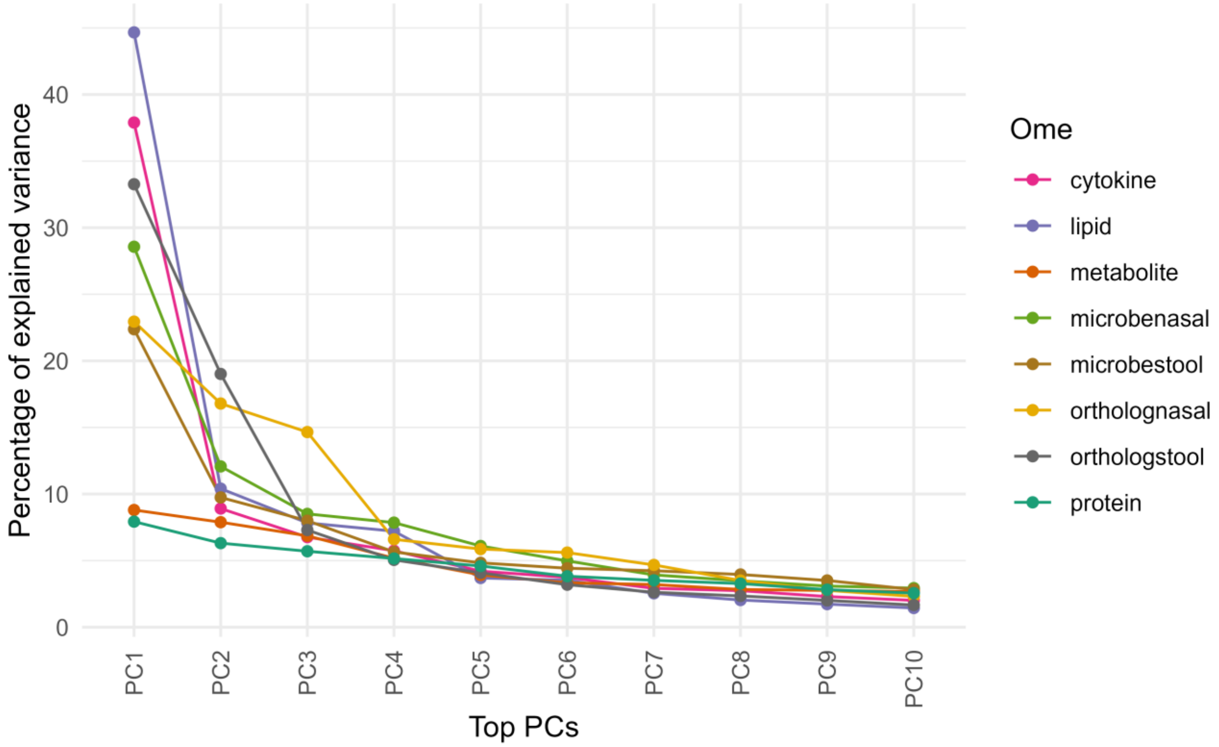

**Supplementary Figure 8. PCA scree plot in iPOP.** For each principal component (PC) within each ome in iPOP, the amount of variation captured by the PC is shown. Only the top 10 PCs are plotted.

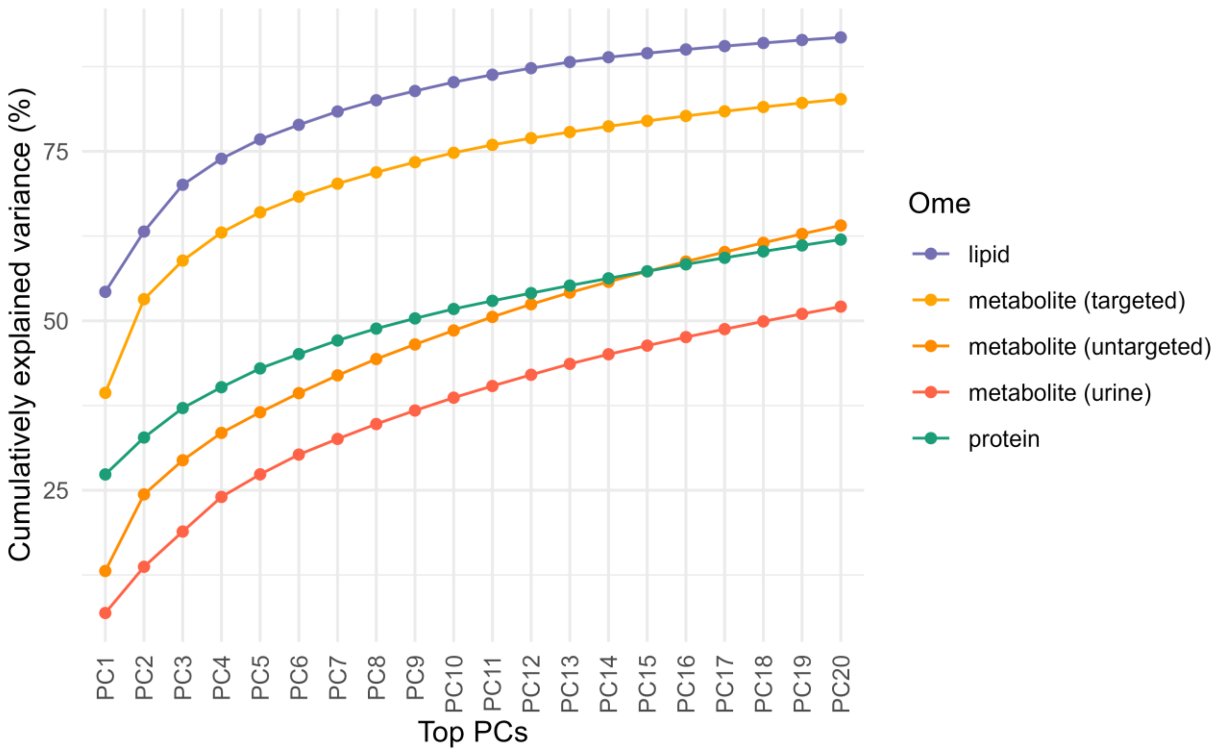

**Supplementary Figure 9. PCA avalanche plot in the replication cohort, hPOP.** For each principal component (PC) and ome in hPOP, the cumulative amount of variation captured by the PC is shown. Only the top 20 PCs are plotted.

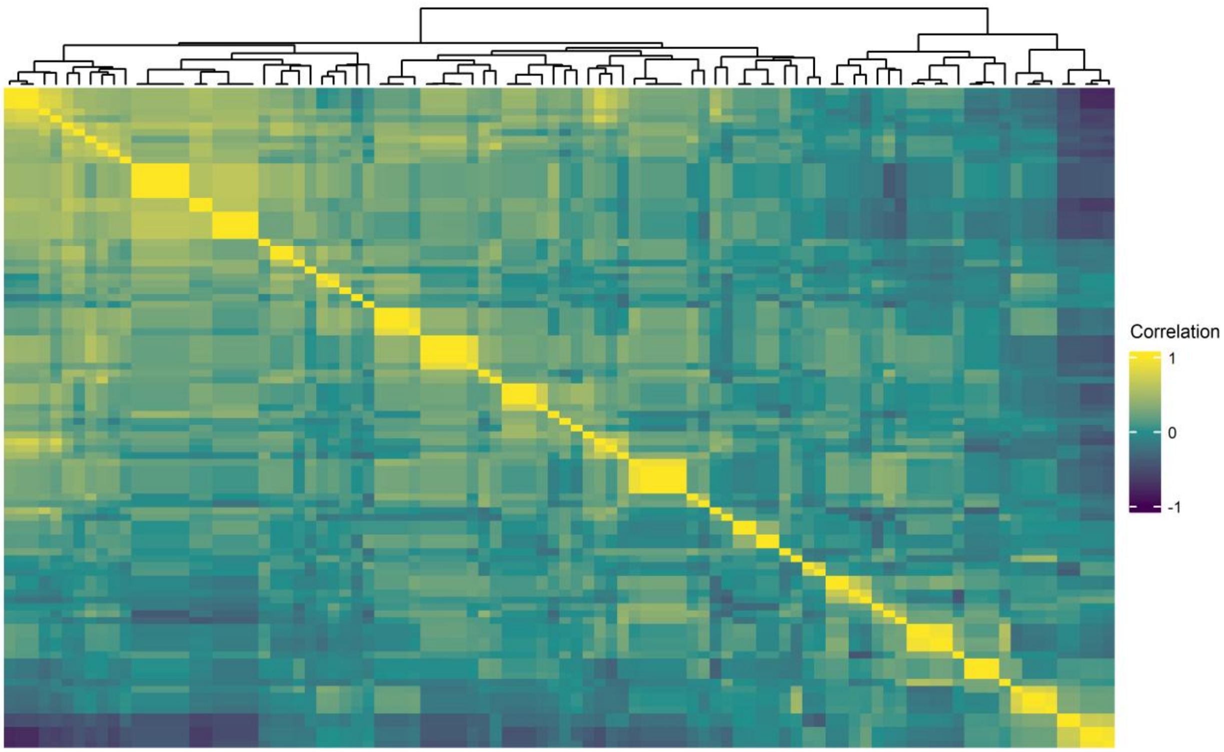

**Supplementary Figure 10. Correlation heat map for the stool microbiome.** The correlation between all pairs of stool microbiome variables is shown.

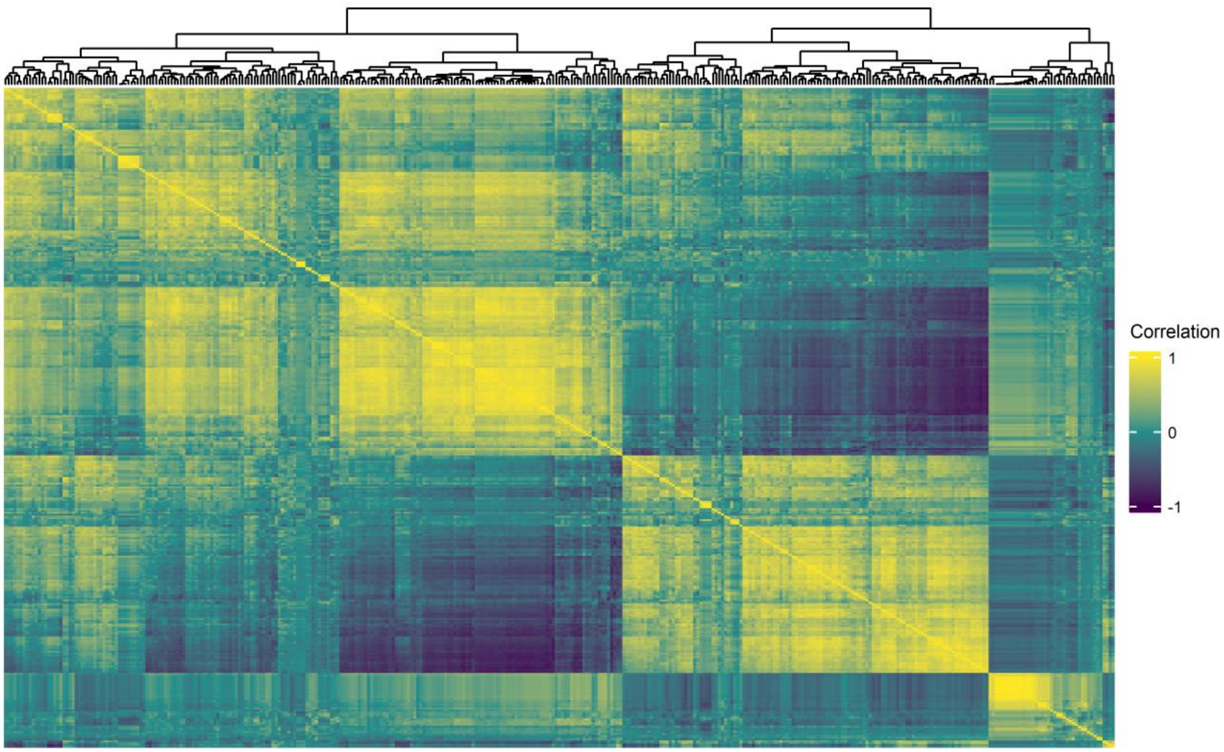

**Supplementary Figure 11. Correlation heat map for stool orthologs.** The correlation between all pairs of stool microbiome ortholog variables is shown.

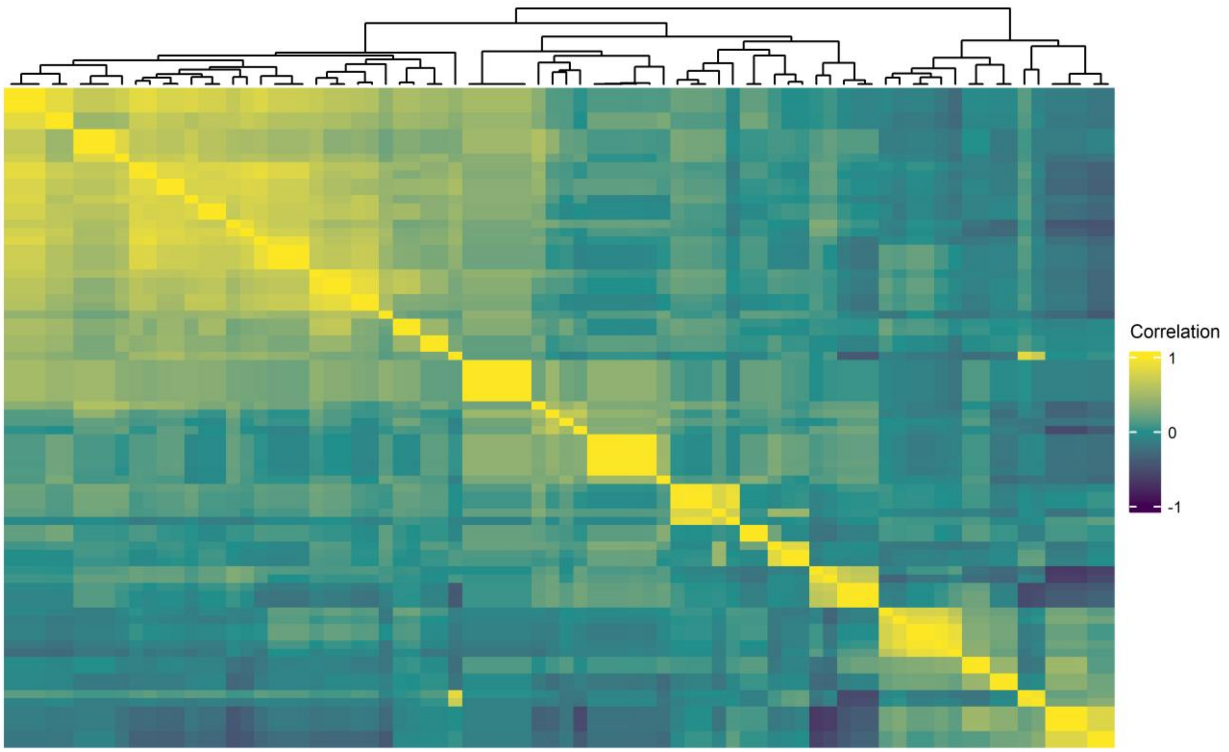

**Supplementary Figure 12. Correlation heat map for the nasal microbiome.** The correlation between all pairs of nasal microbiome variables is shown.

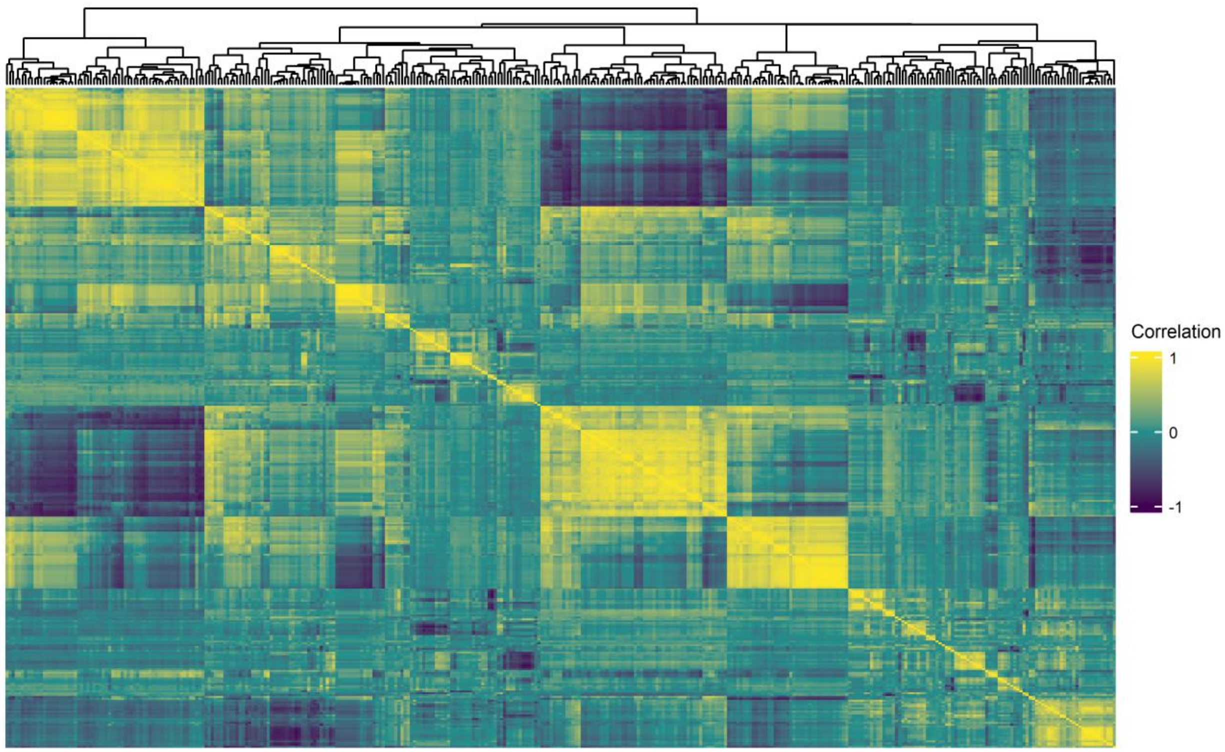

**Supplementary Figure 13. Correlation heat map for nasal orthologs.** The correlation between all pairs of nasal microbiome ortholog variables is shown.

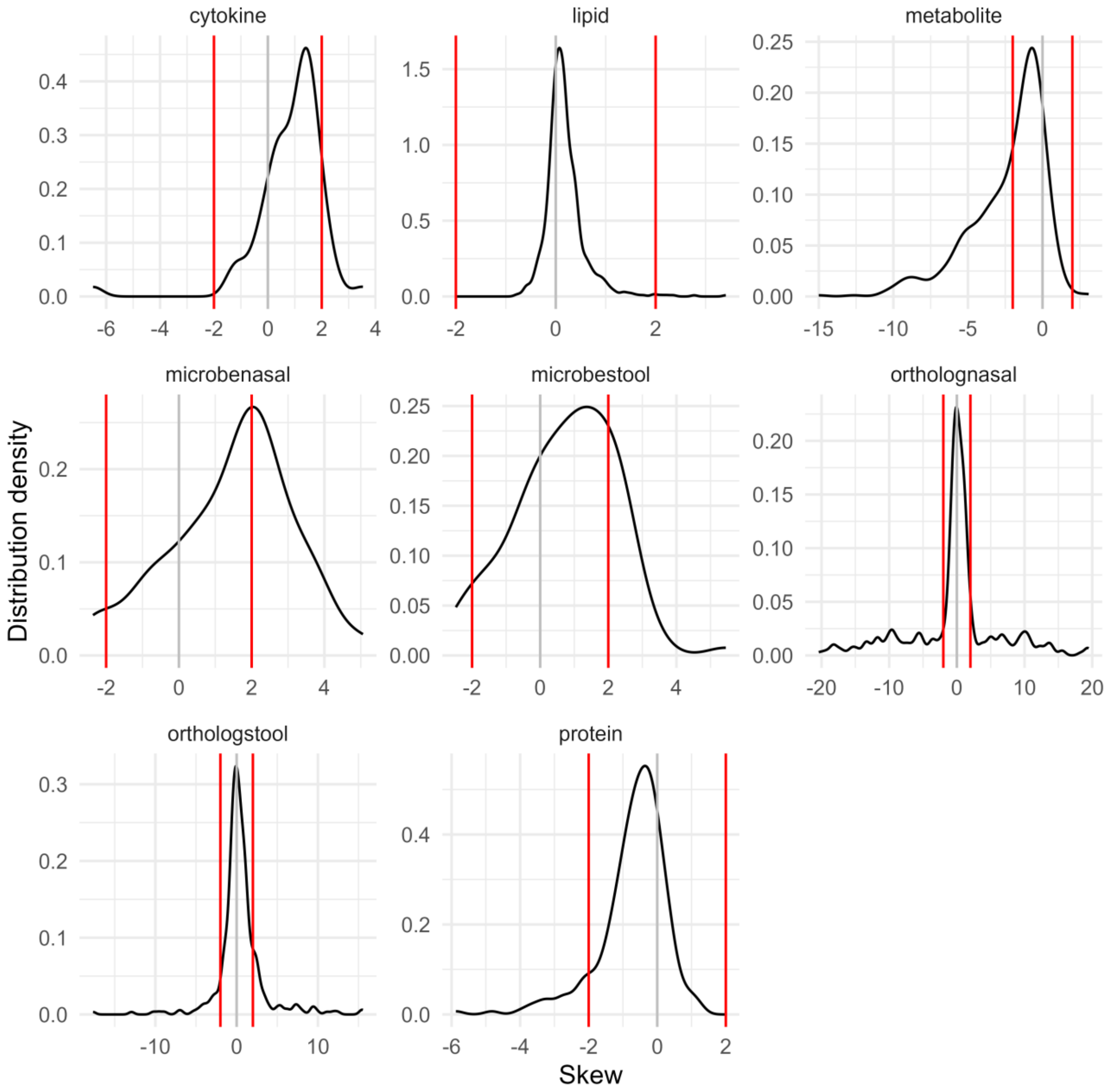

**Supplementary Figure 14. Variable skew after transformation.** The distribution of skew values of the transformed molecular variables are shown for each ome. The black line indicates the distribution density plot, the gray line indicates the value of 0 skew, and the two red lines indicate the thresholds to be considered skewed (-2, 2).

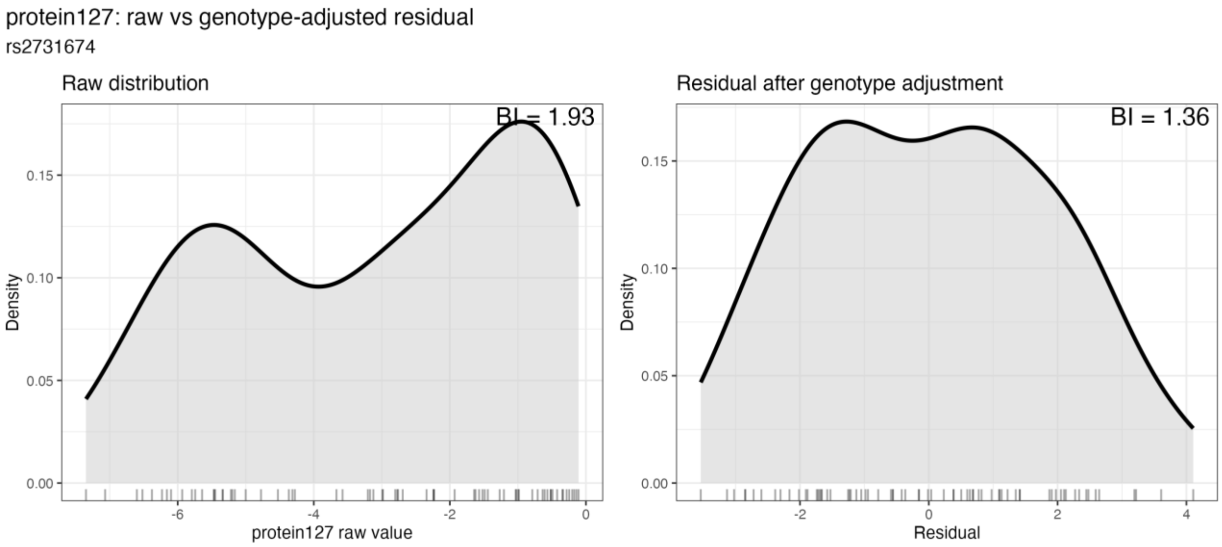

**Supplementary Figure 15. Distribution of SERPINA5 values before and after controlling for its pQTL.** Density plots show the distribution of SERPINA5 protein levels before adjustment (left) and the residual distribution after linear regression adjustment for rs2731674 allele dosage (right). Bimodality index (BI) values are shown in each panel.

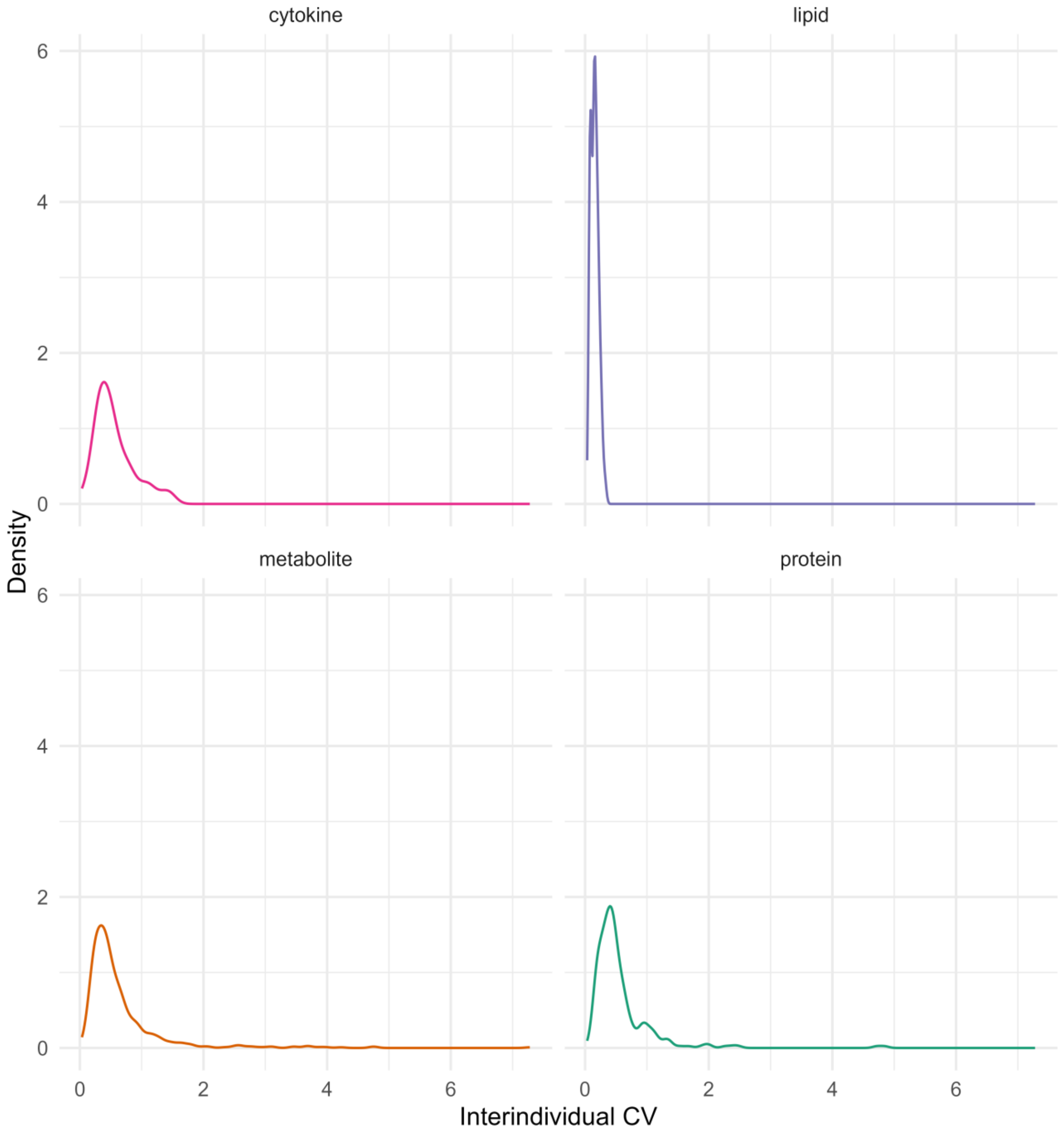

**Supplementary Figure 16. Distribution of interindividual CVs by ome.** The distribution of the interindividual coefficients of variation for molecules from the four blood-based omes are shown. For each molecule measured, the CV was calculated on the untransformed values of the molecule using the median value per participant for that molecule.

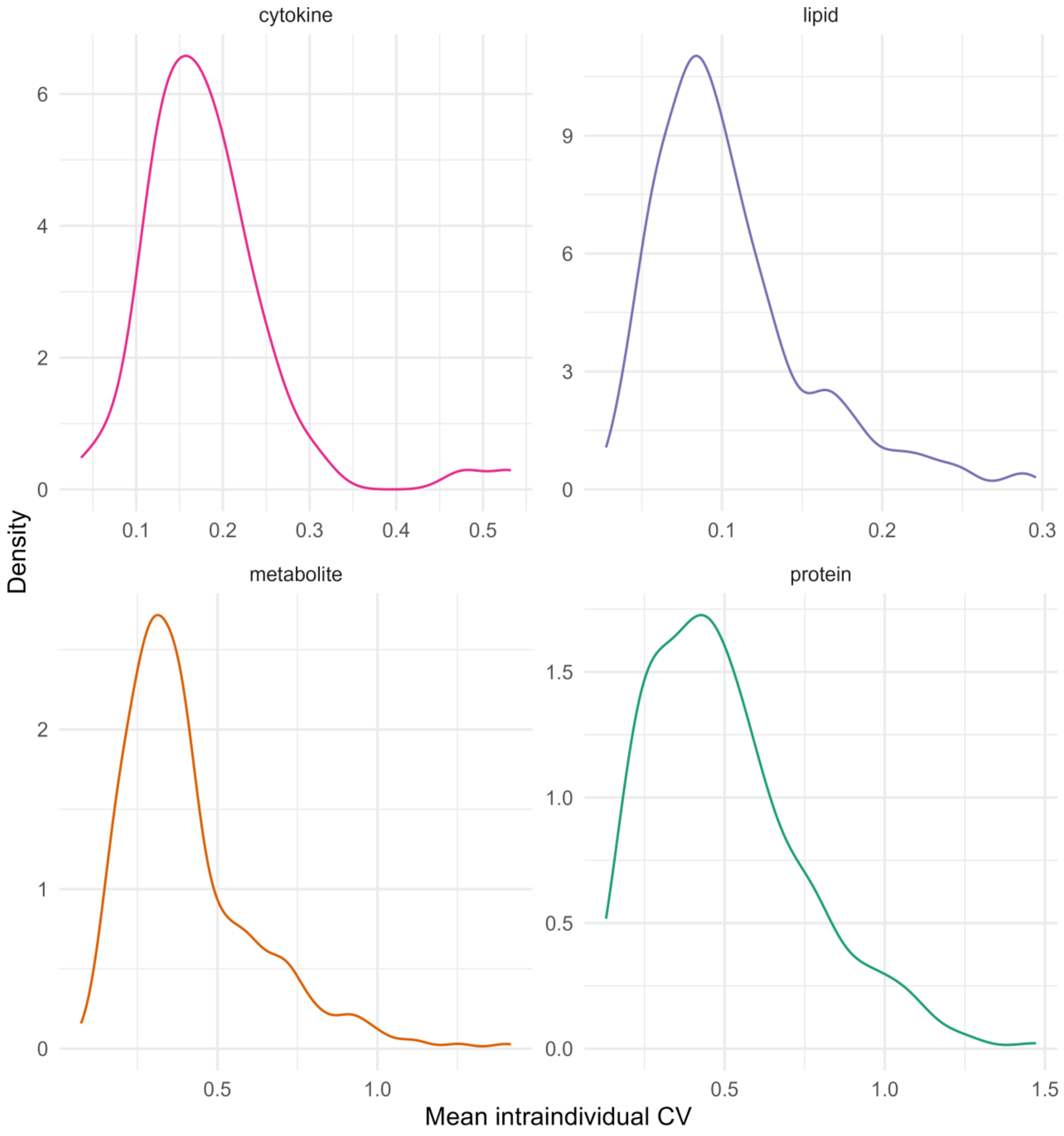

**Supplementary Figure 17. Distribution of intraindividual CVs by ome.** The distribution of the intraindividual coefficients of variation for molecules from the four blood-based omes are shown. For each molecule measured, the CV was calculated on the untransformed values of that molecule from all samples per participant. The mean intraindividual CV for each person was then calculated across all participants.

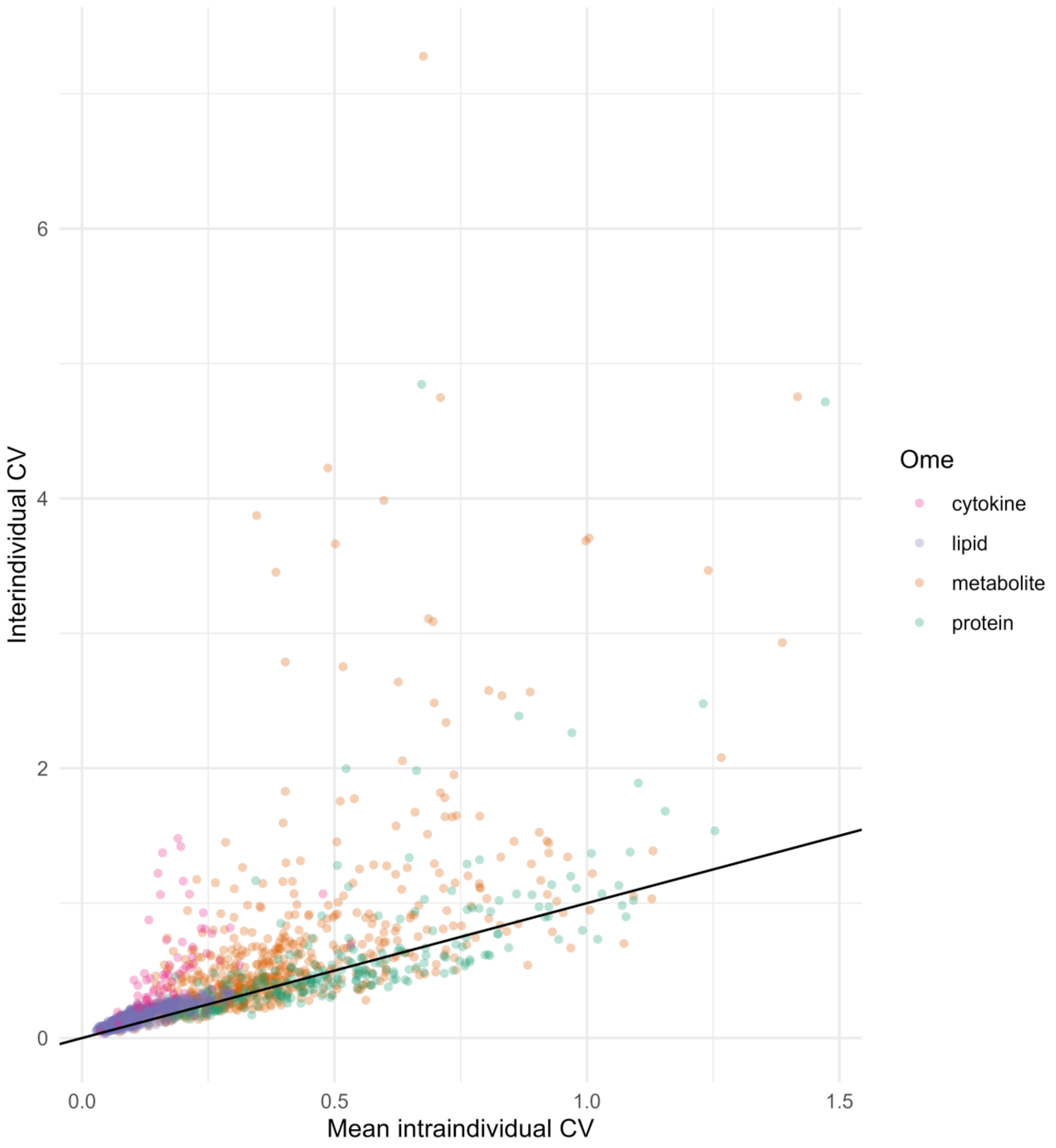

**Supplementary Figure 18. Comparison of interindividual and intraindividual CVs.** For each molecule from the four blood-based omes, both the mean intraindividual CV and the interindividual CV for that molecule is plotted. The black line shows where the intraindividual and interindividual CV values are identical.

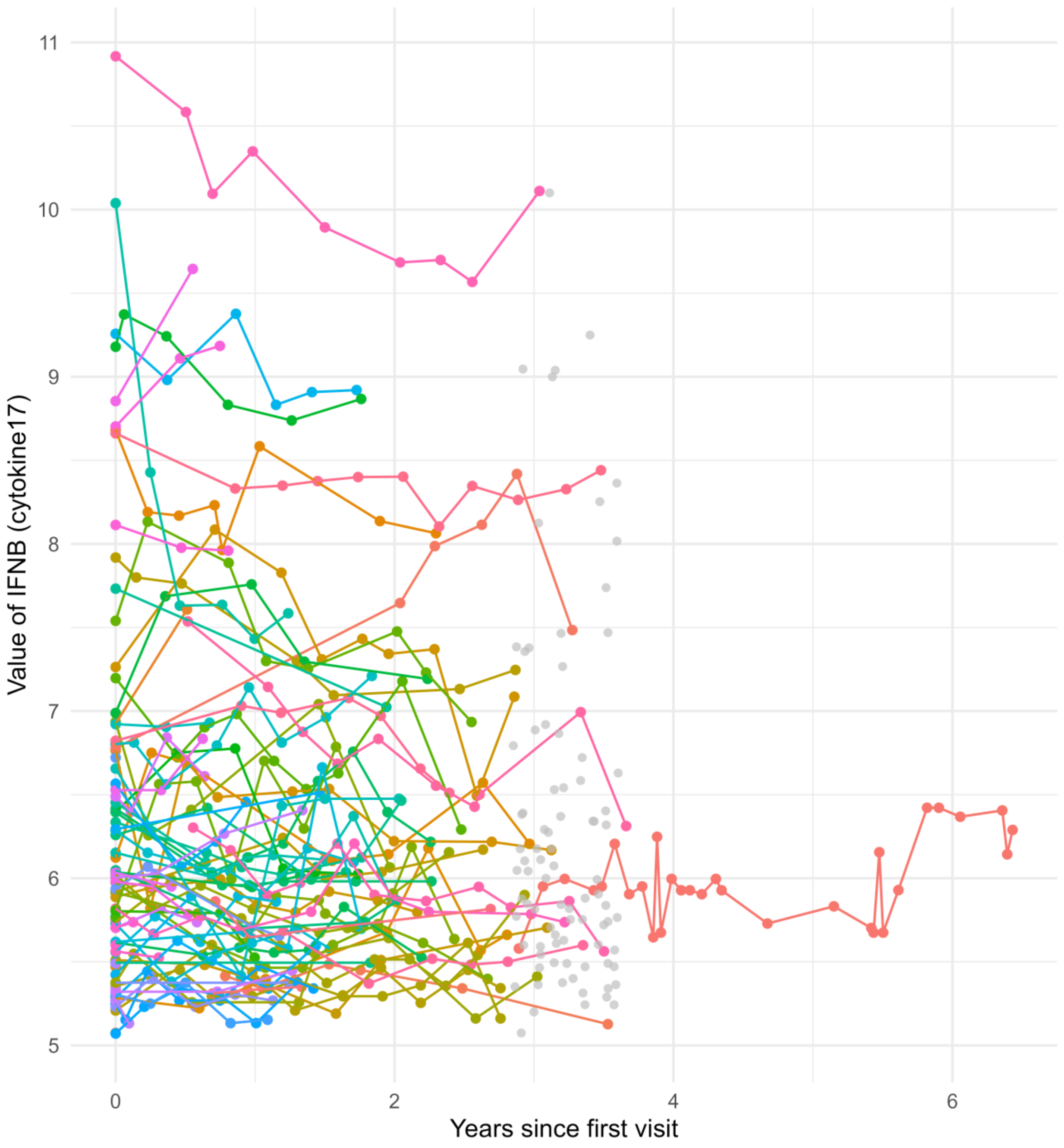

**Supplementary Figure 19. Longitudinal trajectory of IFNB.** The individual trajectories of interferon-beta (IFNB) are shown in colored lines and points. The gray points in the center of the figure show the median values for each individual across time points. This molecule's interindividual CV was greater than the mean intraindividual CV.

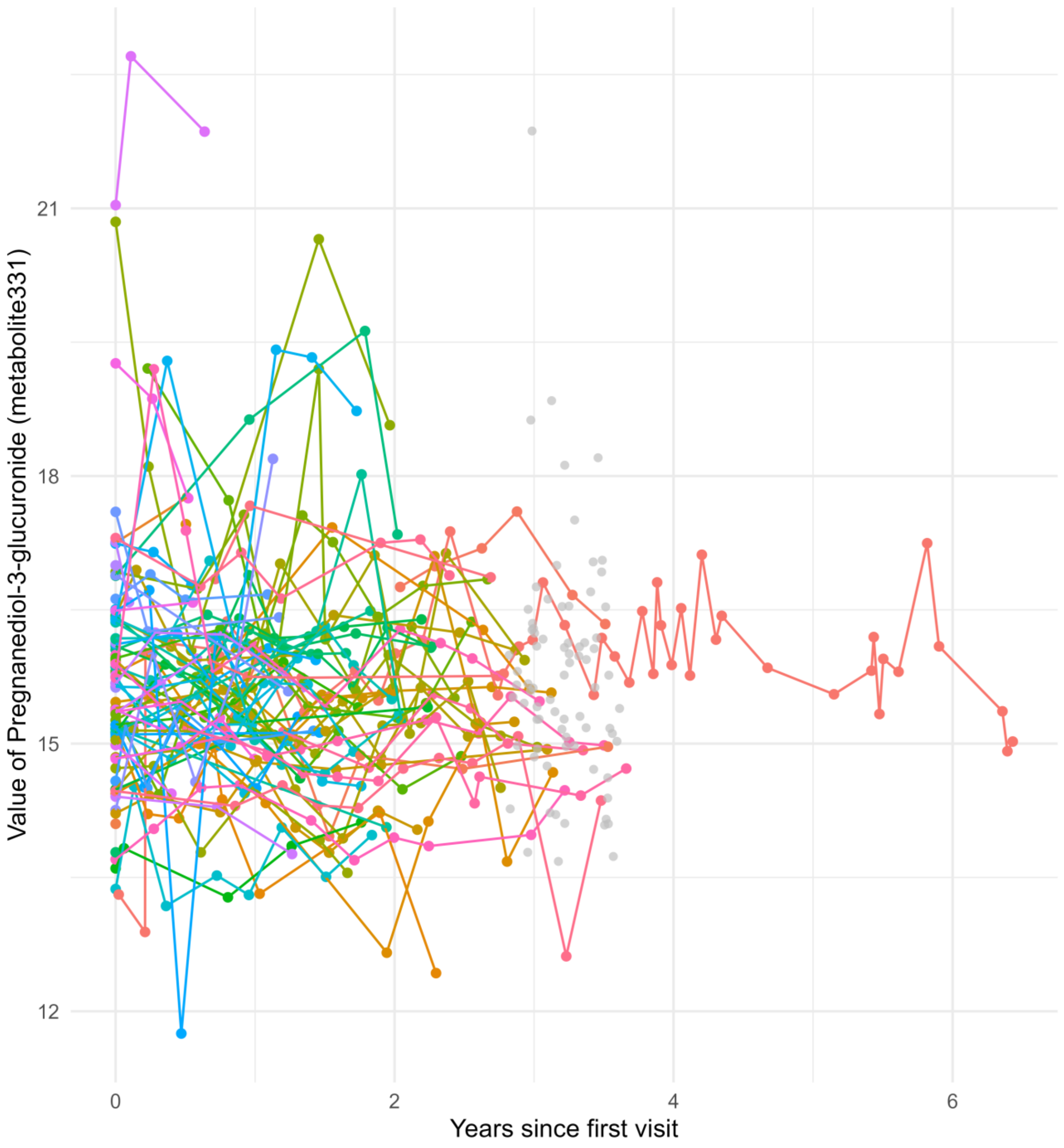

**Supplementary Figure 20. Longitudinal trajectory of pregnanediol-3-glucuronide.** The individual trajectories of pregnanediol-3-glucuronide are shown in colored lines and points. The gray points in the center of the figure show the median values for each individual across time points. This molecule's interindividual CV was greater than the mean intraindividual CV.

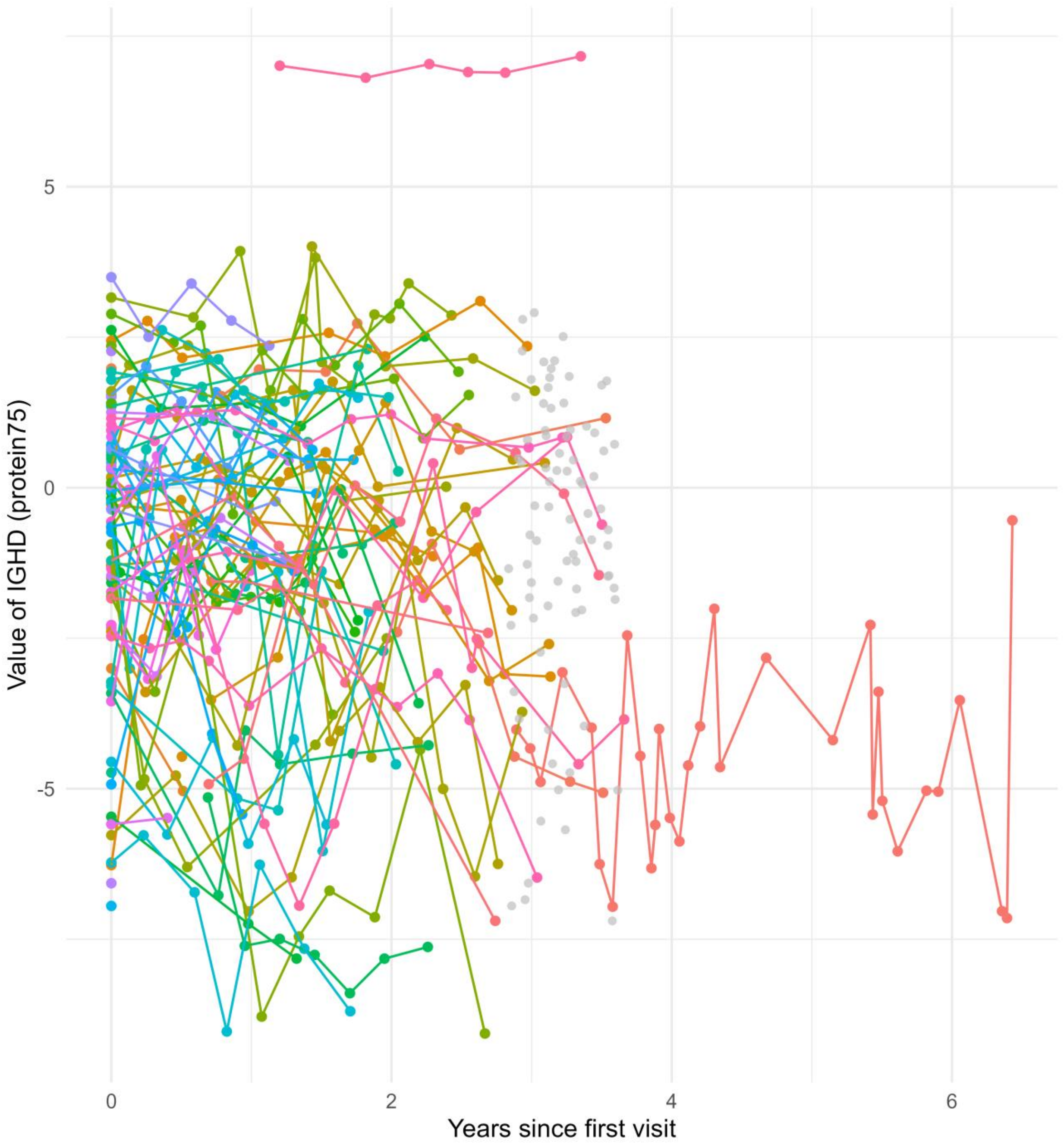

**Supplementary Figure 21. Longitudinal trajectory of IGHD.** The individual trajectories of IGHD are shown in colored lines and points. The gray points in the center of the figure show the median values for each individual across time points. This molecule's interindividual CV was greater than the mean intraindividual CV.

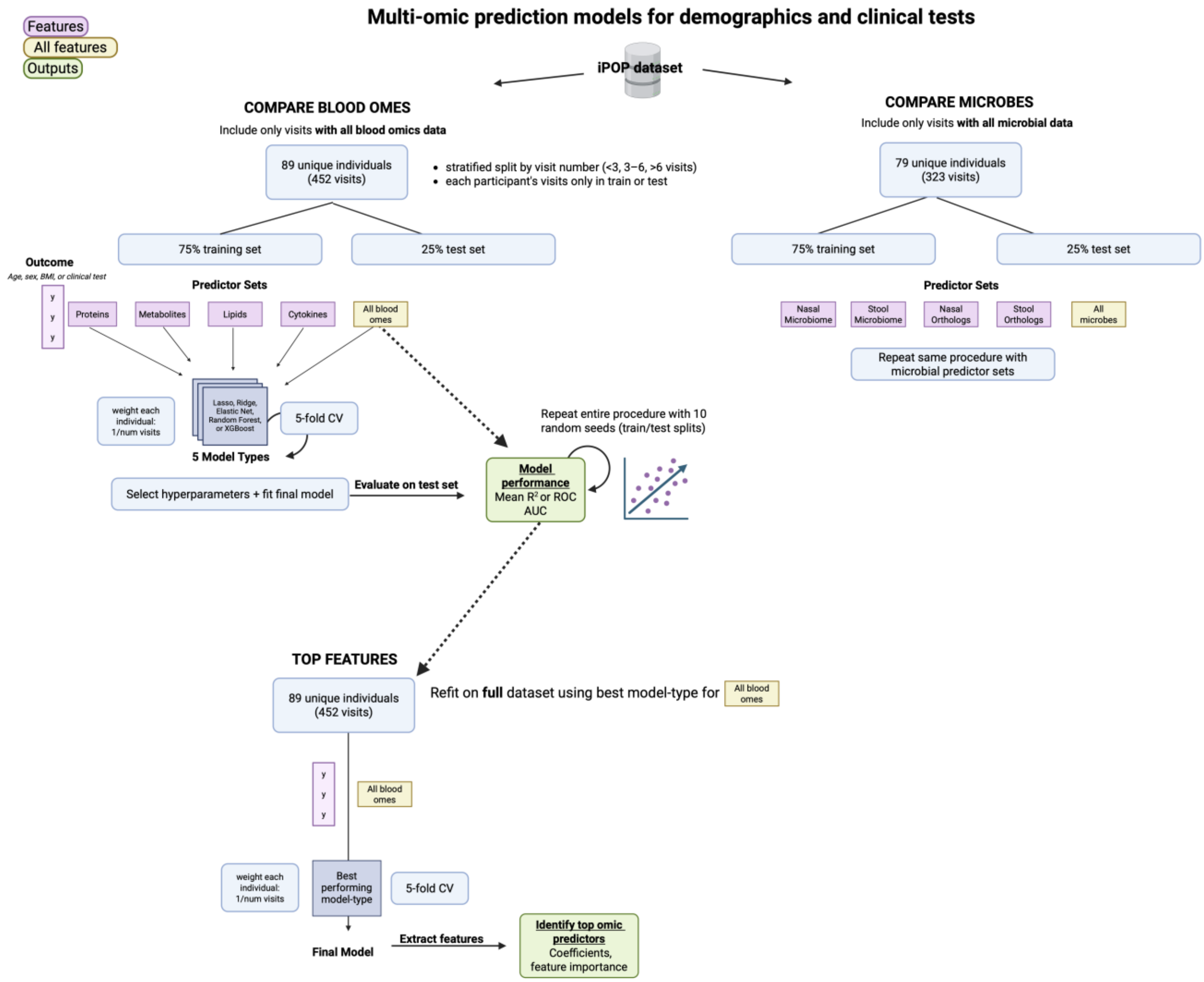

**Supplementary Figure 22. Overview of the machine-learning models for predicting demographics and clinical tests.** We separated blood omic data and microbial data to maximize the input size of the training/test data sets (n=452 and n=323, respectively). In general, microbial data was less routinely collected. We trained five different model types (LASSO regression, ridge regression, elastic net, random forest, and XGBoost) using five-fold cross validation to select hyperparameters. Predictors included each blood ome and microbe data set individually, or “all blood omes” or “all microbes” combined. Outcome variables included age, “phenoage,” sex, BMI, or a clinical test. We extracted top features from a final “all blood omes” or “all microbes” model refit on the entire data set (e.g., n=452 for blood omes, or n=323 for microbes).

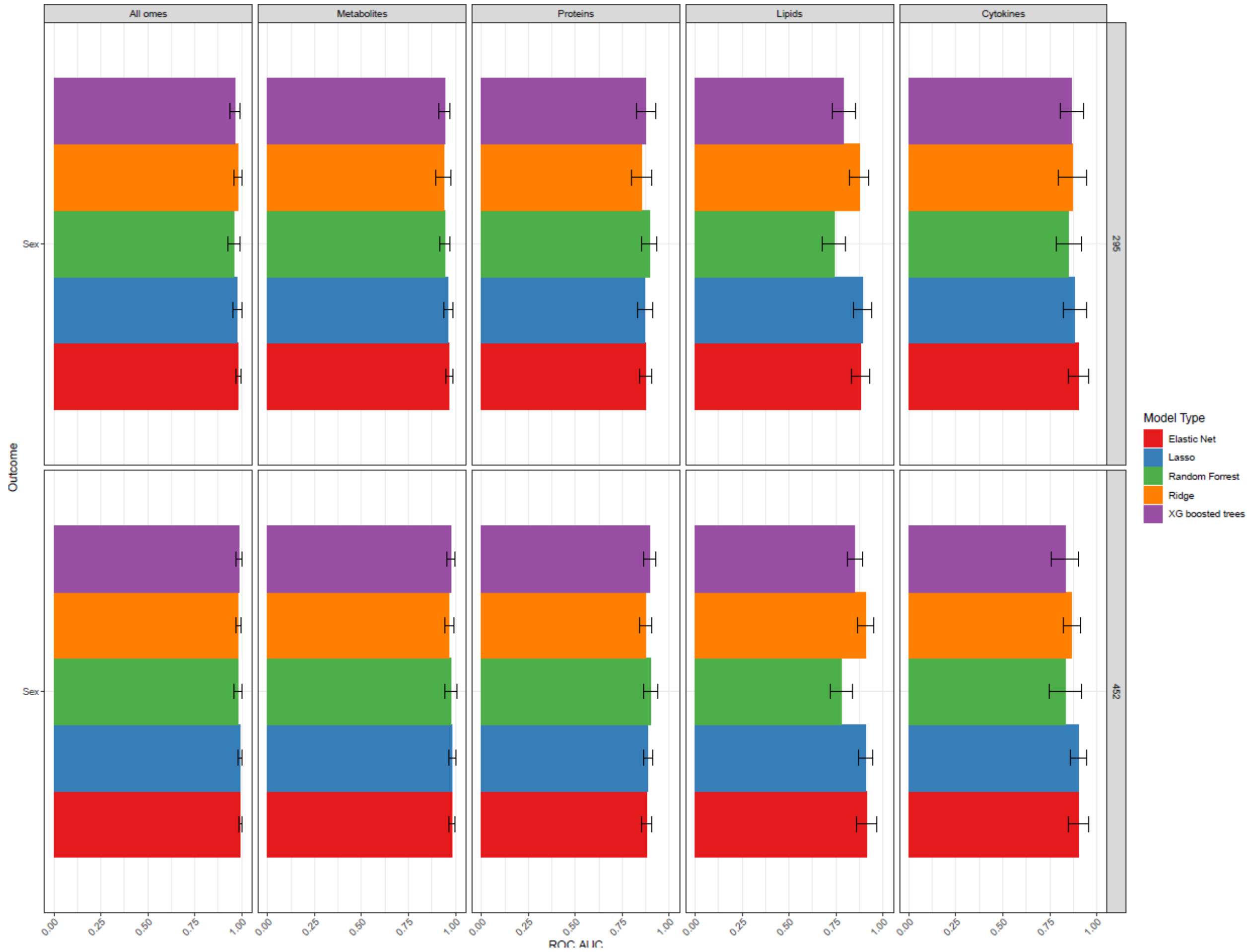

**Supplementary Figure 23. Sensitivity analysis of machine-learning models predicting sex.** The ROC AUC values for each model type and each blood ome predictor set is shown, trained on either the "complete" data set with 295 samples ("295") or the "all blood omes" data set ("452"). The performance of the models was similar though slightly worse in the "complete" data set.

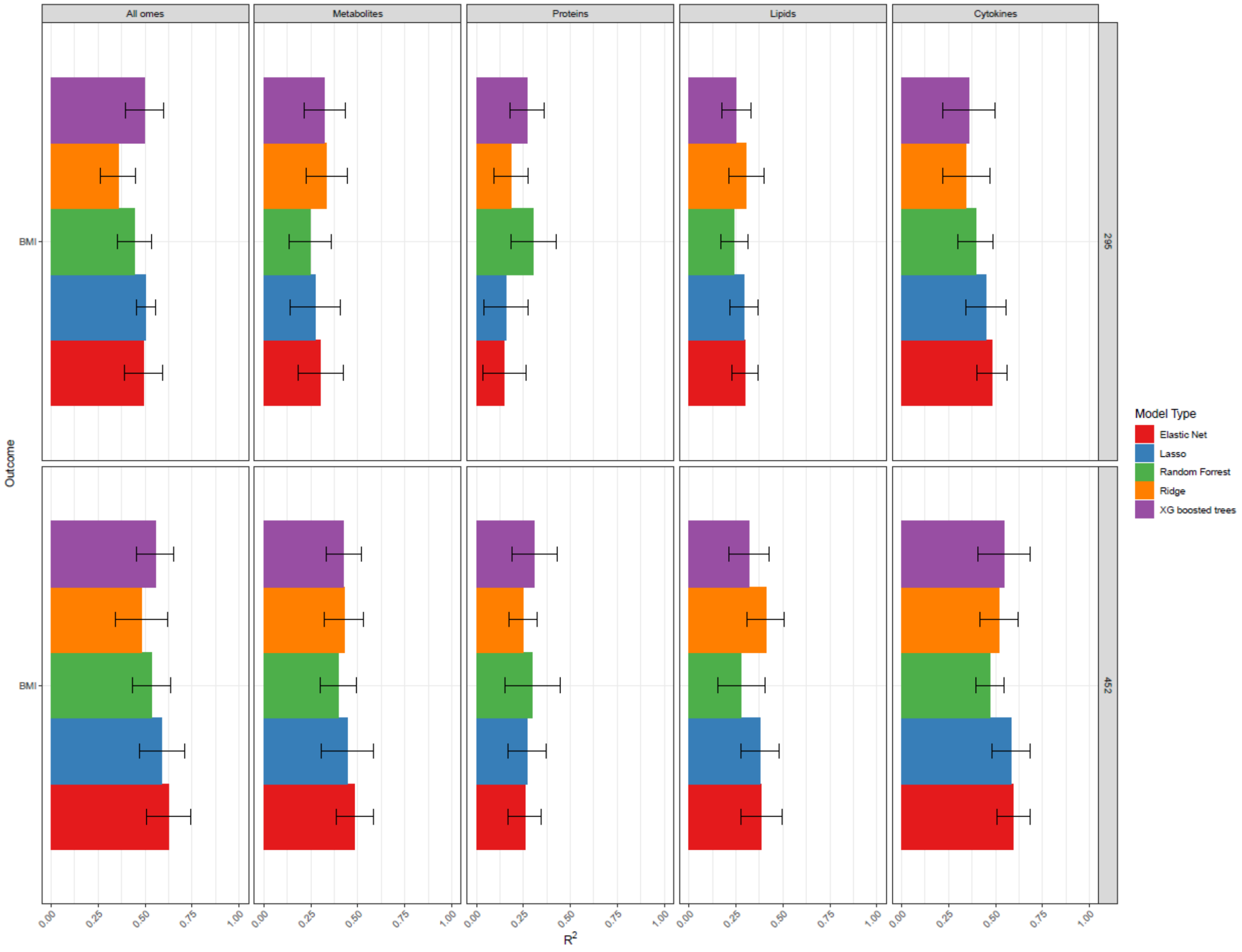

**Supplementary Figure 24. Sensitivity analysis of machine-learning models predicting BMI.** The $R^2$ values for each model type and each blood ome predictor set is shown, trained on either the "complete" data set with 295 samples ("295") or the "all blood omes" data set ("452"). The performance of the models was similar though slightly worse in the "complete" data set.

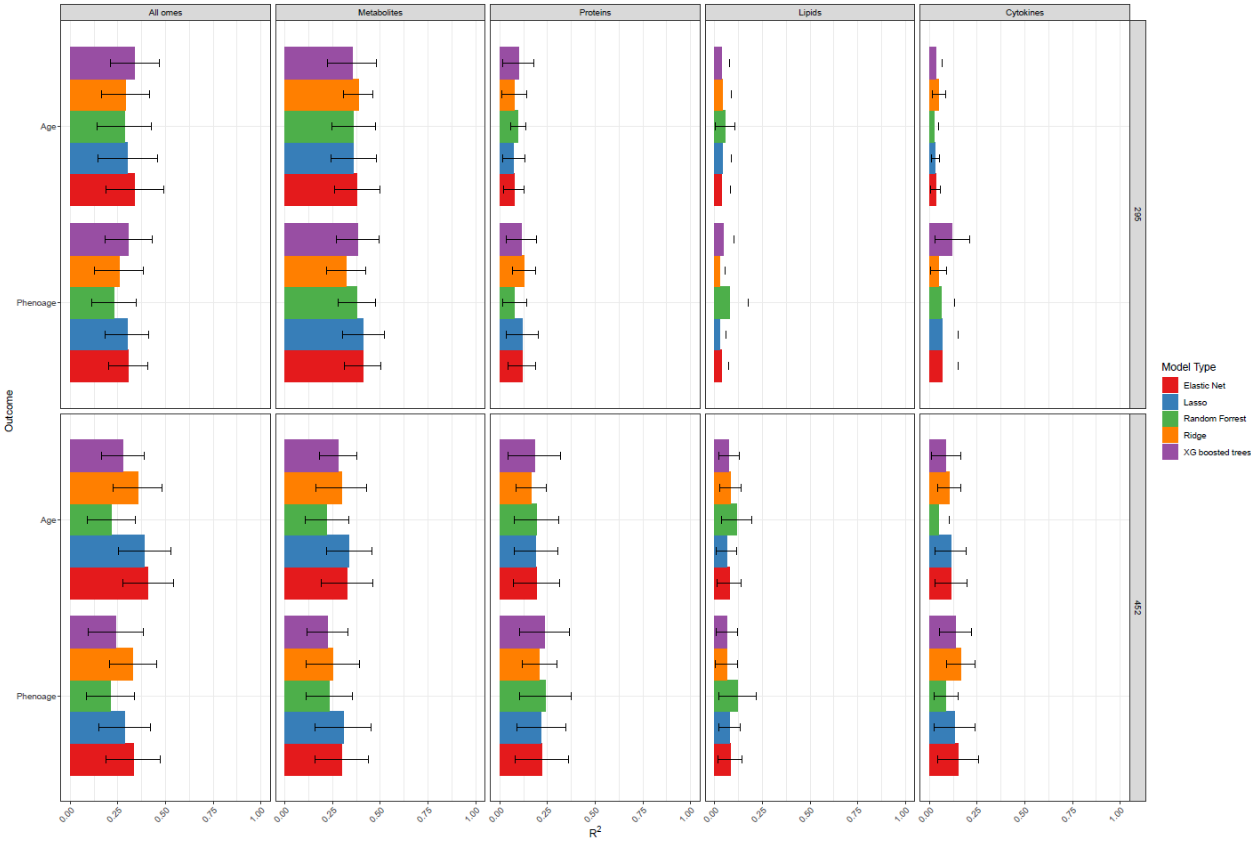

**Supplementary Figure 25. Sensitivity analysis of machine-learning models predicting age and PhenoAge.** The $R^2$ values for each model type and each blood ome predictor set is shown, trained on either the "complete" data set with 295 samples ("295") or the "all blood omes" data set ("452"). Generally, the performance of the models was similar though slightly worse in the "complete" data set. The main exception was with the metabolomic models, where performance was consistently better in the smaller data set.

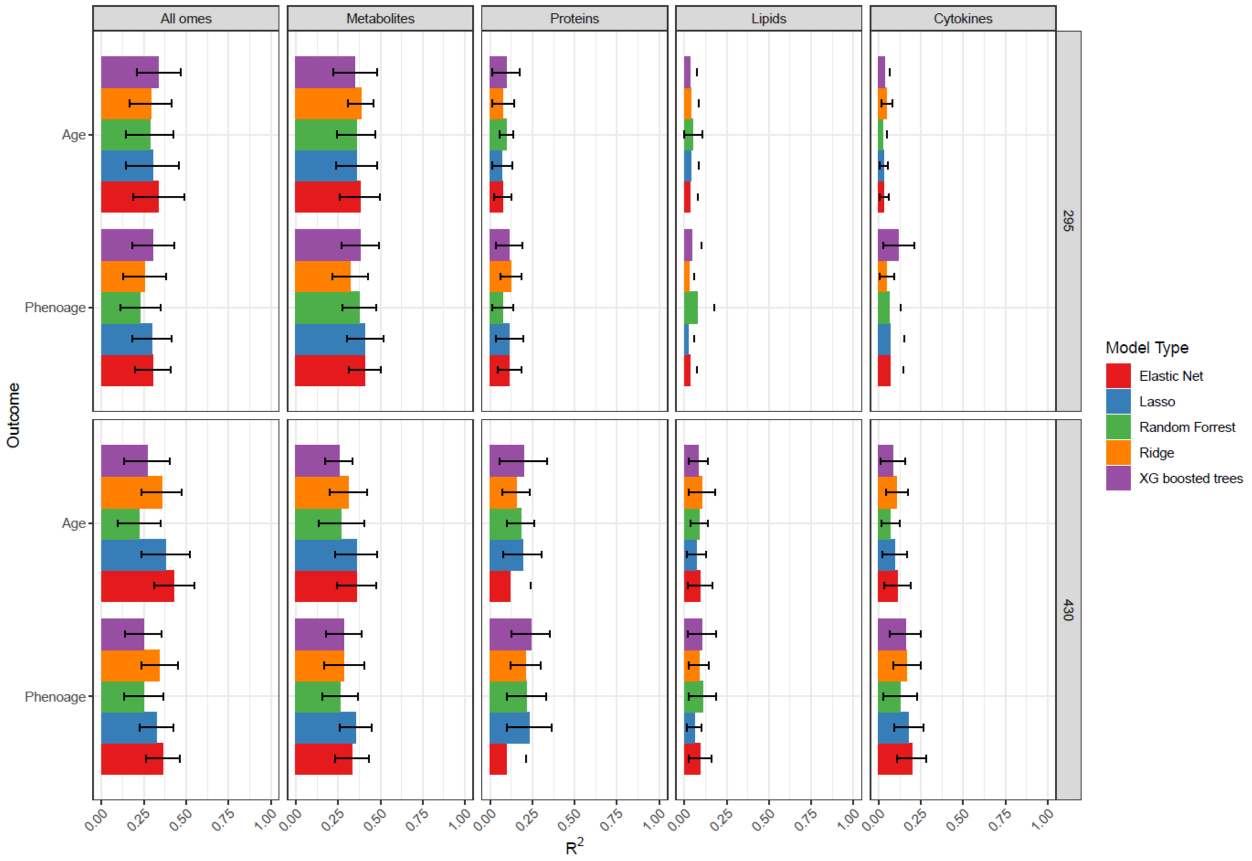

**Supplementary Figure 26. Sensitivity analysis of machine-learning models predicting age and PhenoAge with overrepresented participant excluded.** The $R^2$ values for each model type and each blood ome predictor set is shown, trained on either the “complete” data set with 295 samples (“295”) or the “all blood omes” data set with the overrepresented participant excluded (“430”). The counterintuitive performance decline for the metabolomic models in the larger data set remained even with the overrepresented participant excluded, indicating that overfitting on that participant was unlikely to be driving the performance drop in the larger data set.

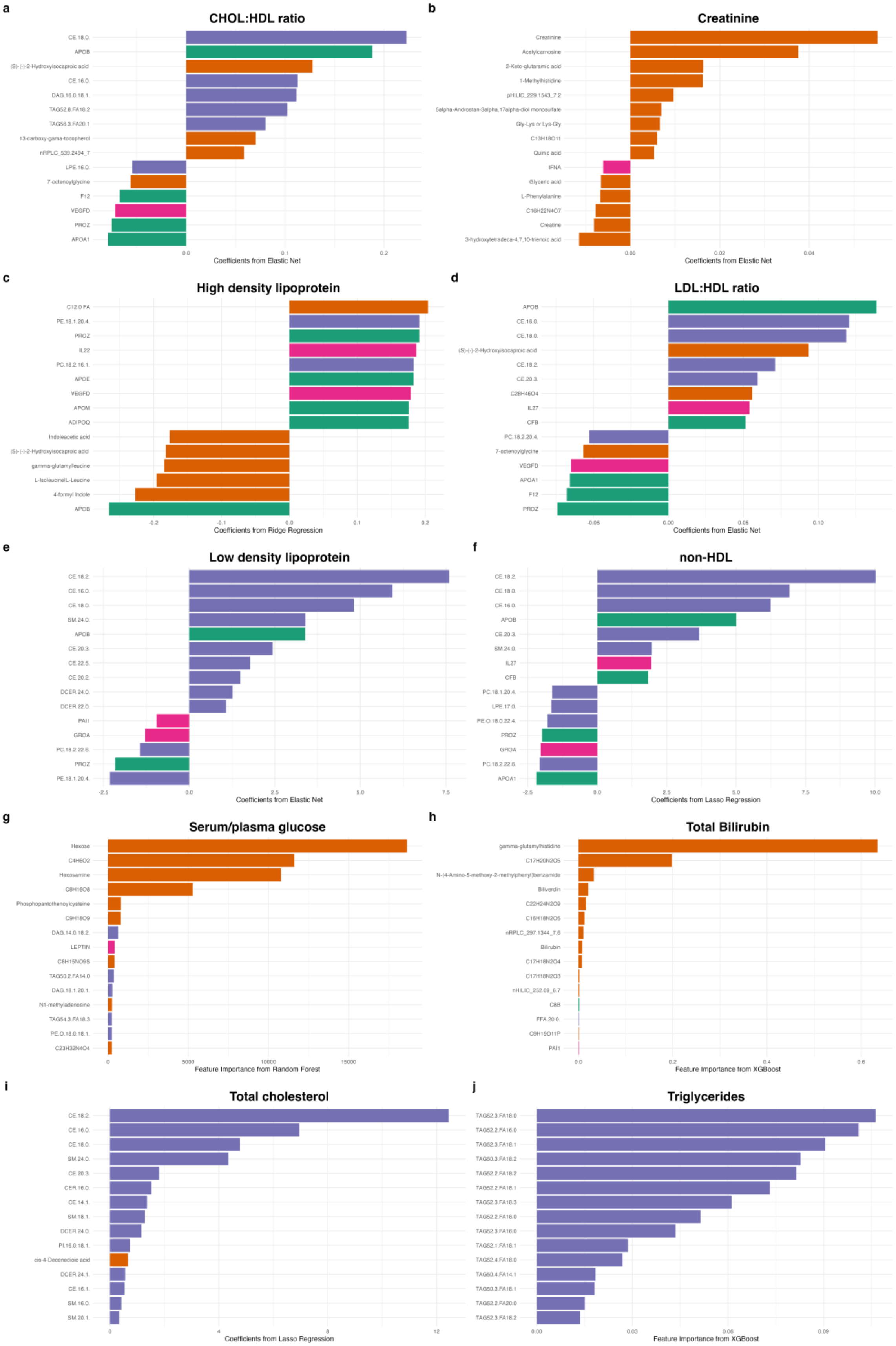

a
CHOL:HDL ratio
CE.18.0.
APOB
(S)-(-)-2-Hydroxyisocaproic acid
CE.16.0.
DAG.16.0.18.1.
TAG52.8.FA18.2
TAG56.3.FA20.1
13-carboxy-gama-tocopherol
nRPLC_539.2494_7
LPE.16.0.
7-octenoylglycine
F12
VEGFD
PROZ
APOA1
0.0
0.1
0.2
Coefficients from Elastic Net
b
Creatinine
Creatinine
Acetylcarnosine
2-Keto-glutaramic acid
1-Methylhistidine
pHILIC_229.1543_7.2
5alpha-Androstan-3alpha,17alpha-diol monosulfate
Gly-Lys or Lys-Gly
C13H18O11
Quinic acid
IFNA
Glyceric acid
L-Phenylalanine
C16H22N4O7
Creatine
3-hydroxytetradeca-4,7,10-trienoic acid
0.00
0.02
0.04
Coefficients from Elastic Net
c
High density lipoprotein
C12.0 FA
PE.18.1.20.4.
PROZ
IL22
PC.18.2.16.1.
APOE
VEGFD
APOM
ADIPOQ
Indoleacetic acid
(S)-(-)-2-Hydroxyisocaproic acid
gamma-glutamylleucine
L-Isoleucine|L-Leucine
4-formyl Indole
APOB
-0.2
-0.1
0.0
0.1
0.2
Coefficients from Ridge Regression
d
LDL:HDL ratio
APOB
CE.16.0.
CE.18.0.
(S)-(-)-2-Hydroxyisocaproic acid
CE.18.2.
CE.20.3.
C28H46O4
IL27
CFB
PC.18.2.20.4.
7-octenoylglycine
VEGFD
APOA1
F12
PROZ
-0.05
0.00
0.05
0.10
Coefficients from Elastic Net
e
Low density lipoprotein
CE.18.2.
CE.16.0.
CE.18.0.
SM.24.0.
APOB
CE.20.3.
CE.22.5.
CE.20.2.
DCER.24.0.
DCER.22.0.
PAI1
GROA
PC.18.2.22.6.
PROZ
PE.18.1.20.4.
-2.5
0.0
2.5
5.0
7.5
Coefficients from Elastic Net
f
non-HDL
CE.18.2.
CE.18.0.
CE.16.0.
APOB
CE.20.3.
SM.24.0.
IL27
CFB
PC.18.1.20.4.
LPE.17.0.
PE.O.18.0.22.4.
PROZ
GROA
PC.18.2.22.6.
APOA1
-2.5
0.0
2.5
5.0
7.5
10.0
Coefficients from Lasso Regression
g
Serum/plasma glucose
Hexose
C4H6O2
Hexosamine
C8H16O8
Phosphopantothenoylcysteine
C9H18O9
DAG.14.0.18.2.
LEPTIN
C8H15NO9S
TAG50.2.FA14.0
DAG.18.1.20.1.
N1-methyladenosine
TAG54.3.FA18.3
PE.O.18.0.18.1.
C23H32N4O4
0
5000
10000
15000
Feature Importance from Random Forest
h
Total Bilirubin
gamma-glutamylhistidine
C17H20N2O5
N-(4-Amino-5-methoxy-2-methylphenyl)benzamide
Biliverdin
C22H24N2O9
C16H18N2O5
nRPLC_297.1344_7.6
Bilirubin
C17H18N2O4
C17H18N2O3
nHILIC_252.09_6.7
C8B
FFA.20.0.
C9H19O11P
PAI1
0.0
0.2
0.4
0.6
Feature Importance from XGBoost
i
Total cholesterol
CE.18.2.
CE.16.0.
CE.18.0.
SM.24.0.
CE.20.3.
CER.16.0.
CE.14.1.
SM.18.1.
DCER.24.0.
PI.16.0.18.1.
cis-4-Decenedioic acid
DCER.24.1.
CE.16.1.
SM.16.0.
SM.20.1.
0
4
8
12
Coefficients from Lasso Regression
j
Triglycerides
TAG52.3.FA18.0
TAG52.2.FA16.0
TAG52.3.FA18.1
TAG50.3.FA18.2
TAG52.2.FA18.2
TAG52.2.FA18.1
TAG52.3.FA18.3
TAG52.2.FA18.0
TAG52.3.FA16.0
TAG52.1.FA18.1
TAG52.4.FA18.0
TAG50.4.FA14.1
TAG50.3.FA18.1
TAG52.2.FA20.0
TAG52.3.FA18.2
0.00
0.03
0.06
0.09
Feature Importance from XGBoost

**Supplementary Figure 27. Top predictors of clinical test models classified as platform validation.** The top omic features are shown for 10 clinical tests that were both highly predicted by one or more omes ($R^2$ > 0.5) and whose models included predictors analogous or highly similar to the measured clinical analyte.

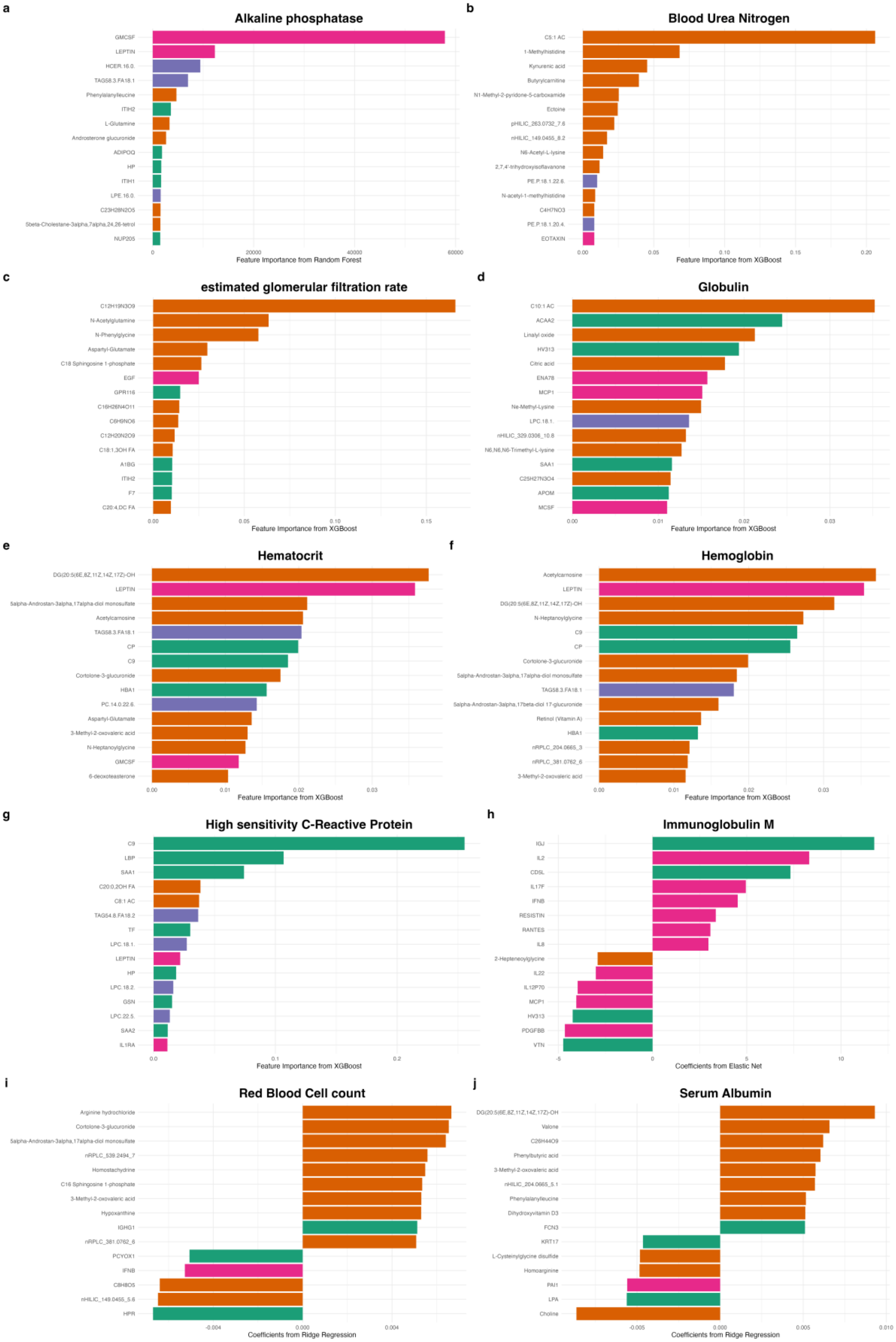
a
Alkaline phosphatase
Feature Importance from Random Forest
b
Blood Urea Nitrogen
Feature Importance from XGBoost
c
estimated glomerular filtration rate
Feature Importance from XGBoost
d
Globulin
Feature Importance from XGBoost
e
Hematocrit
Feature Importance from XGBoost
f
Hemoglobin
Feature Importance from XGBoost
g
High sensitivity C-Reactive Protein
Feature Importance from XGBoost
h
Immunoglobulin M
Coefficients from Elastic Net
i
Red Blood Cell count
Coefficients from Ridge Regression
j
Serum Albumin
Coefficients from Ridge Regression

**Supplementary Figure 28. Top predictors of clinical test models classified as having potential clinical utility.** The top omic features are shown for 10 clinical tests that were modestly predicted by one or more omes ($R^2 > 0.2$) and whose models did not include predictors analogous to the measured clinical analyte. Note: immunoglobulin M (IgM) was predicted by immunoglobin J chain (IGJ), a protein component that links immunoglobulin monomers, and hemoglobin was predicted by hemoglobin subunit alpha 1 (HBA1), a direct structural component of hemoglobin. Although these predictors were closely related to their respective clinical analytes, the models were not classified as "platform validation" because their $R^2$ values were modest and these individual features did not clearly dominate the remaining feature importance or coefficient values.

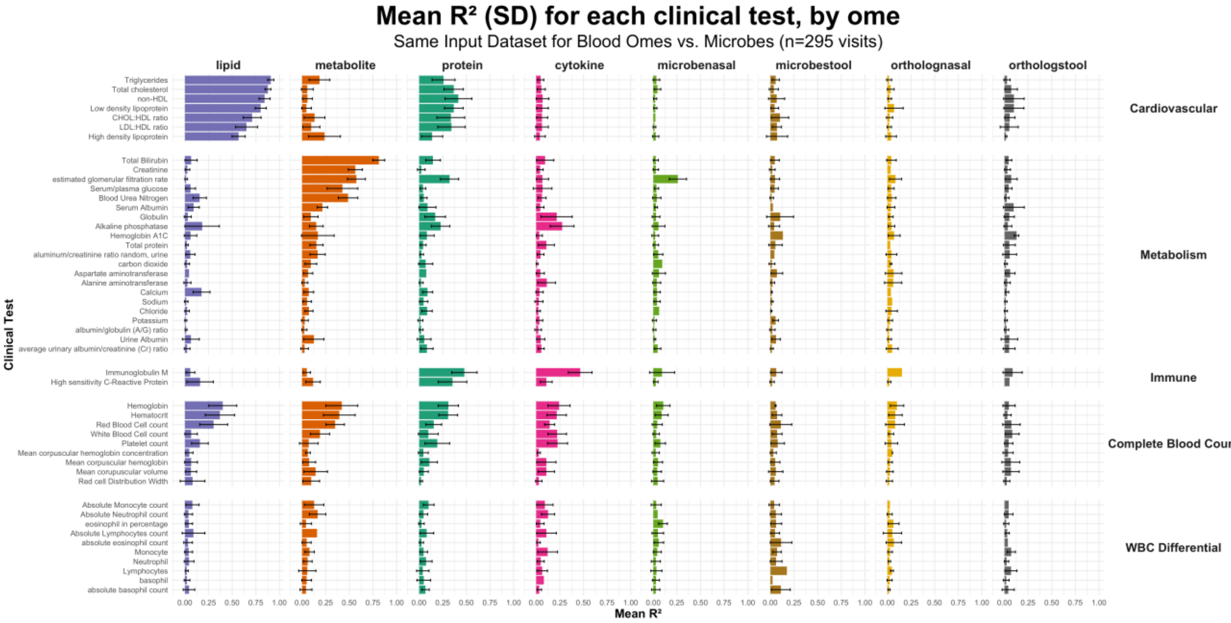

**Supplementary Figure 29. Comparison of blood omes and microbial features in predicting clinical tests when using data sets of the same size.** To evaluate whether the smaller size of the microbiome train/test dataset (n = 323 visits) influenced our findings compared to the blood omes data set (n = 452 visits), we reran all blood omes and microbiome models on a smaller, shared data set of 295 visits where all data were available. In this sensitivity analysis, we again observed that the microbiome underperformed relative to blood omes in predicting clinical tests.

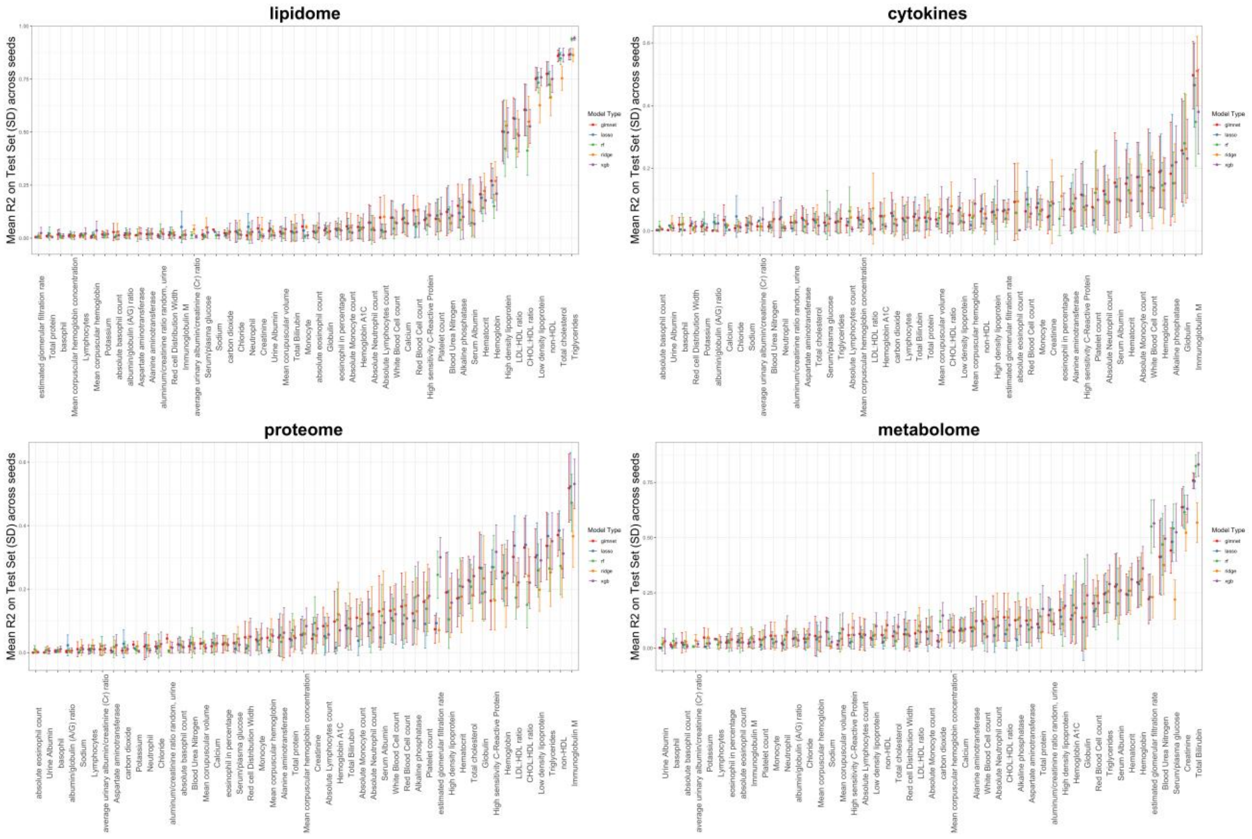

**Supplementary Figure 30. Comparison of machine-learning model types for predicting clinical tests.** The mean R2 and standard deviation for five model types in predicting each clinical test is shown. Model types included elastic net (glmnet), LASSO, random forest, ridge regression, and XGBoost and were implemented with the tidymodels package in R.
